# Linking regulatory variants to target genes by integrating single-cell multiome methods and genomic distance

**DOI:** 10.1101/2024.05.24.24307813

**Authors:** Elizabeth Dorans, Karthik Jagadeesh, Kushal Dey, Alkes L. Price

## Abstract

Methods that analyze single-cell paired RNA-seq and ATAC-seq multiome data have shown great promise in linking regulatory elements to genes. However, existing methods differ in their modeling assumptions and approaches to account for biological and technical noise—leading to low concordance in their linking scores—and do not capture the effects of genomic distance. We propose pgBoost, an integrative modeling framework that trains a non-linear combination of existing linking strategies (including genomic distance) on fine-mapped eQTL data to assign a probabilistic score to each candidate SNP-gene link. We applied pgBoost to single-cell multiome data from 85k cells representing 6 major immune/blood cell types. pgBoost attained higher enrichment for fine-mapped eSNP-eGene pairs (e.g. 21x at distance >10kb) than existing methods (1.2-10x; p-value for difference = 5e-13 vs. distance-based method and < 4e-35 for each other method), with larger improvements at larger distances (e.g. 35x vs. 0.89-6.6x at distance >100kb; p-value for difference < 0.002 vs. each other method). pgBoost also outperformed existing methods in enrichment for CRISPR-validated links (e.g. 4.8x vs. 1.6-4.1x at distance >10kb; p-value for difference = 0.25 vs. distance-based method and < 2e-5 for each other method), with larger improvements at larger distances (e.g. 15x vs. 1.6-2.5x at distance >100kb; p-value for difference < 0.009 for each other method). Similar improvements in enrichment were observed for links derived from Activity-By-Contact (ABC) scores and GWAS data. We further determined that restricting pgBoost to features from a focal cell type improved the identification of SNP-gene links relevant to that cell type. We highlight several examples where pgBoost linked fine-mapped GWAS variants to experimentally validated or biologically plausible target genes that were not implicated by other methods. In conclusion, a non-linear combination of linking strategies, including genomic distance, improves power to identify target genes underlying GWAS associations.

## Introduction

More than 90% of disease-associated genetic variants implicated in genome-wide association studies (GWAS) lie outside of protein-coding regions of the genome, often within regulatory elements hypothesized to act by modulating the expression of nearby genes^1–7^. However, non-coding variants often do not regulate the nearest gene^8–11^; thus, our understanding of the biological mechanisms underlying these associations remains limited^12,13^. Previous experimental and computational methods have leveraged transcriptomic, epigenomic, chromatin accessibility, and 3D contact information to infer tissue-agnostic or tissue-specific links between non-coding elements and their target genes^2,10,14–17^, but methods analyzing data from bulk tissues or cell lines may be underpowered to detect cell-type-specific patterns^3,18–20^. Nascent single-cell multiome technologies co-assaying chromatin accessibility and gene expression (scRNA/ATAC-seq) enable the detection of *in vivo* regulatory relationships by correlating the accessibility of candidate regulatory elements (as measured by ATAC-seq peaks) with gene expression (as measured by RNA-seq) across individual cells^21–31^. While existing single-cell peak-gene linking methods have shown promise in detecting regulatory links in diverse cell types and states^25–31^, we observe low concordance in their linking scores (**Figure 1a**); in addition, these methods underperform a simple genomic distance-based linking score in their enrichment for fine-mapped eSNP-eGene pairs (**Figure 1b**) and enrichment for other evaluation sets of links derived from independent data (see below).

**Figure 1.**
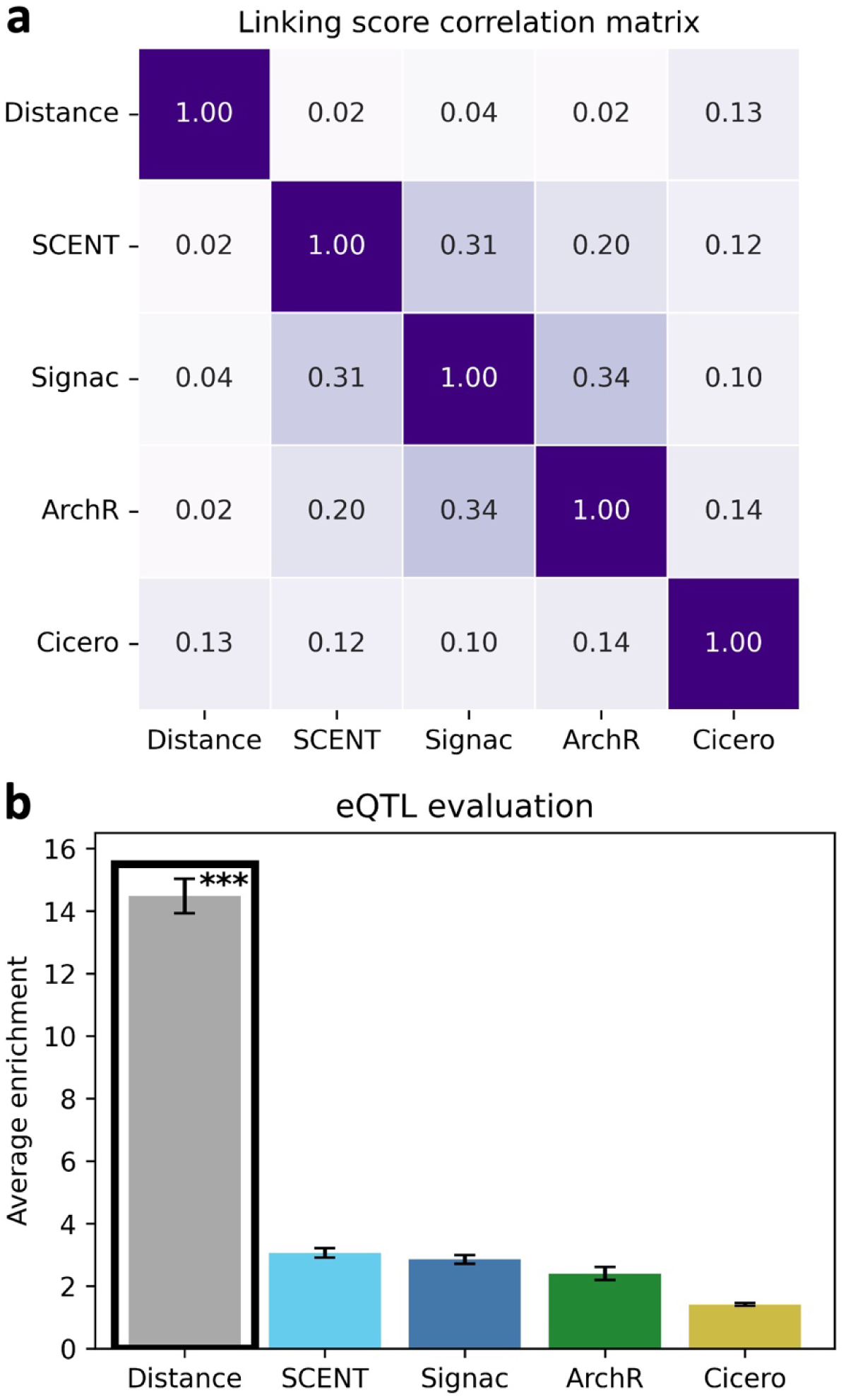
Existing single-cell peak-gene linking methods exhibit low concordance and underperform a distance-based linking score. **a)** Pearson correlation matrix of linking scores predicted by genomic distance from TSS and 4 single-cell peak-gene linking methods (**Table 1** and **Supplementary Table 1**) across 2,387,131 candidate SNP-gene links scored by any method in T cells from the 10X PBMC data set^47^. **b)** Average enrichment across recall values (in [0,0.35]) of links predicted by distance and 4 single-cell peak-gene linking methods (applied to 3 single-cell multiome data sets; **Table 2** and **Supplementary Table 2**) for fine-mapped eSNP-eGene pairs attaining maximum PIP > 0.5 across all GTEx tissues (**Methods**). Confidence intervals denote standard errors. Stars denote p-values for difference (***: p < 0.001) of top method (denoted in black outline) vs. each other method (actual p-values < 5e-119 vs. each method). Numerical results are reported in **Supplementary Table 3**.

**Table 1.** Overview of 4 single-cell peak-gene linking methods. For each method linking ATAC-seq peaks to genes, we indicate the input data (either scRNA/ATAC-seq multiome data or scATAC-seq data), scoring procedure, significance testing approach (if applicable), and candidate link selection criteria. Further details are provided in **Methods** and **Supplementary Table 1**.

| Method | Input data | Scoring procedure | Significance testing | Candidate link criteria |
| --- | --- | --- | --- | --- |
| SCENT<sup>25</sup> | RNA + ATAC | Poisson regression effect size | Bootstrapping across cells | Peak & gene in $\geq 5\%$ of cells |
| Signac<sup>26</sup> | RNA + ATAC | Correlation across single cells | Permutation across peaks | Peak & gene in $\geq 10$ cells |
| ArchR<sup>27</sup> | RNA + ATAC | Correlation across metacells | Analytical p-value | N/A |
| Cicero<sup>28</sup> | ATAC | Peak-promoter co-accessibility across metacells | N/A | Promoter overlaps a peak |

**Table 2.** Overview of pgBoost features. We indicate the cell types profiled, number of cells passing QC, experimental protocol, and number of pgBoost features derived from each single-cell multiome data set (plus 2 distance-based features). 10x Multiome: 10x Genomics Single Cell Multiome ATAC + Gene Expression. Further details are provided in **Supplementary Table 2**.

| Data set | Cell types | # cells | Experimental protocol | # pgBoost features |
| --- | --- | --- | --- | --- |
| 10x PBMC<sup>47</sup> | T, B, myeloid | 10,561 | 10x Multiome | 33 |
| Luecken BMMC<sup>48</sup> | T, B, myeloid, erythroid, dendritic | 57,371 | 10x Multiome | 55 |
| SHARE-seq LCL<sup>22</sup> | lymphoblastoid cell line (LCL) | 17,644 | SHARE-Seq | 11 |
| Genomic distance | N/A | N/A | N/A | 2 |
| *All* | *6 cell types* | *85,576 cells* |  | *101 features* |

Here, we propose an eQTL-informed gradient boosting^32^ approach (pgBoost) that integrates linking scores from existing peak-gene linking methods across cell types and data sets with genomic distance, training on fine-mapped eQTL data to assign a single probabilistic score to each candidate SNP-gene link. We evaluate the performance of pgBoost and existing single-cell peak-gene linking methods by evaluating their enrichment for several sets of SNP-gene links derived from eQTL^33,34^, Activity-By-Contact (ABC)^17,35^, CRISPRi^15,36–42^, and GWAS^43,44^ data. We also investigate whether restricting to single-cell data from a focal cell type can improve power to detect regulatory links relevant to that cell type. We highlight examples demonstrating that our integrative approach yields biological insights that can aid in the interpretation of GWAS variant-disease associations. Accurately mapping the regulatory architectures connecting regulatory variants to their target genes can improve our understanding of the molecular mechanisms driving disease and highlight regulatory links of potential therapeutic importance.

## Results

### Overview of Methods

pgBoost integrates information from 4 constituent peak-gene linking scores derived from single-cell RNA/ATAC-seq multiome data^25–27^ (or scATAC-seq data^28^), as well as genomic distance, to predict regulatory SNP-gene links (**Table 1** and **Figure 2**). The constituent scores assess the co-activity of candidate ATAC peaks and genes^25–27^ (or the co-accessibility of candidate peaks and promoter regions of candidate genes^28^) across single cells^25,26^ (or metacells^27,28^) and are computed separately for each multiome data set and cell type. pgBoost optimizes a gradient boosting classification task^32^, using constituent scores and genomic distance as features and fine-mapped GTEx eSNP-eGene pairs^33,34^ as training data (**Figure 2**). The gradient boosting algorithm is a decision tree-based ensemble approach that learns the non-linear relationships among these features to predict the causal probability of candidate SNP-gene links (we caution that causal probabilities are estimated with respect to the training data and may not reflect biological causality).

**Figure 2.**
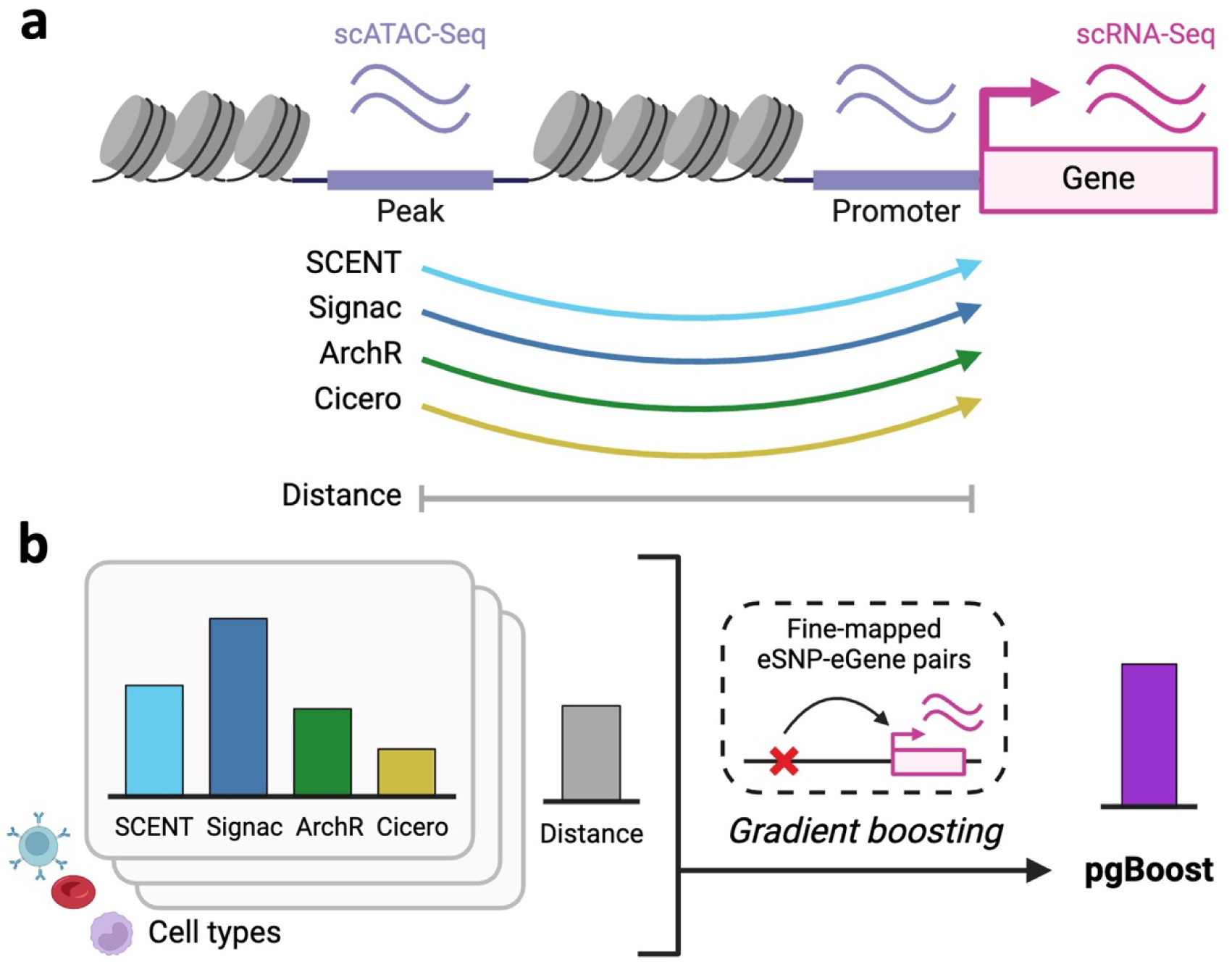
Overview of pgBoost method. **a)** Single-cell peak-gene linking methods assign linking scores measuring the co-activity of ATAC peak-gene pairs. **b)** pgBoost learns a non-linear combination of SNP-gene distance and linking scores across methods, cell types, and data sets to assign probabilistic linking scores using a gradient boosting classification framework and a training set of positive and negative links comprised of fine-mapped eSNP-eGene pairs attaining maximum PIP > 0.5 and maximum PIP < 0.01 across all GTEx tissues, respectively.

In detail, we computed peak-gene linking scores using the existing single-cell methods SCENT^25^, Signac^26^, ArchR^27^, and Cicero^28^, restricting to *cis* peak-gene pairs with peak-gene distance >1kb and <500kb, thus focusing primarily on enhancer-gene links rather than promoter-gene links (**Table 1**, **Supplementary Table 1**, and **Methods**). To integrate linking scores from existing methods across data sets, we intersected data set-specific peaks with 1000 Genomes Project variants with minor allele count ≥ 5^10,45,46^, thus analyzing SNP-gene links instead of peak-gene links. pgBoost features for each candidate SNP-gene link include 11 features for each data set and cell type—4 linking scores produced by the existing methods SCENT, Signac, ArchR, and Cicero in each cell type, 3 corresponding significance levels (Cicero does not output significance level), and 4 binary variables indicating whether the candidate link was scored by each method— and 2 distance-based features: SNP-gene distance and a binary variable indicating whether the candidate SNP links to the gene with closest TSS (**Table 2** and **Methods**). As positive and negative training sets, we used fine-mapped eSNP-eGene pairs assigned maximum PIP > 0.5 and maximum PIP < 0.01 across GTEx tissues^33,34^, respectively. To avoid overfitting, we trained pgBoost using a leave-one-chromosome-out (LOCO) approach, generating predictions on held-out chromosomes. We have publicly released open-source software implementing pgBoost (see Code Availability).

We compared pgBoost to each constituent method (specifically, the top score across data sets and cell types analyzed for each method) and to a distance-based method that ranks candidate SNP-gene links based on their genomic distance (**Methods**). For a given evaluation data set and a given method, we computed *average enrichment across recall values*, defined as the enrichment of predicted SNP-gene links (surviving a score threshold selected to yield a specific value of recall) in the set of evaluation links, averaged across values of recall (ranging from 0 to the maximum recall attained by all constituent scores) (**Methods**). To evaluate performance at more proximal vs. more distal links, we computed average enrichment when restricting candidate and evaluation SNP-gene links to various distance thresholds ranging from >1kb to >100kb. We conservatively restrict the set of candidate SNP-gene links used in all computations to SNP-gene pairs involving a SNP lying in an ATAC-seq peak (not all *cis* SNP-gene pairs as in ref.^25^), leading to much lower enrichment for all methods. As a secondary metric, we also computed the area under the precision-recall curve (AUPRC), stratified by distance threshold (**Methods**).

We analyzed 3 scRNA/ATAC-seq multiome data sets (10X PBMC^47^, Luecken BMMC^48^, SHARE-seq LCL^22^) that each spanned 1-5 immune/blood cell types and 11k-57k cells after QC (total of 9 data set-cell type pairs, spanning 6 cell types and 85K cells) (**Table 2**, **Supplementary Table 2**, **Supplementary Figure 1**, and **Methods**; see Data Availability). This resulted in 101 pgBoost features (combined in a single model), including 2 features based on genomic distance. We computed each of these features for 4,503,599 candidate SNP-gene links spanning 476,186 SNPs (lying in scATAC-seq peaks in any of the cell types in this data) and 17,443 genes. We have publicly released linking scores and percentiles for every method considered in this study (see Data Availability).

### pgBoost improves power to identify SNP-gene links predicted by eQTL and ABC

We constructed pgBoost predictions and assessed whether SNP-gene links predicted by pgBoost recapitulated SNP-gene links predicted by eQTL (via fine-mapped eSNP-eGene pairs^33,34^) and Activity-By-Contact (ABC)^17,35^. We note that although SNP-gene links predicted by eQTL/ABC are only partially accurate, high enrichment with respect to these evaluation sets is still indicative of high power to detect true links. In addition, each evaluation set that we analyze could alternately be considered as a prediction or used as a training data set (see below). In primary analyses, pgBoost uses the eQTL evaluation set for training. We use a LOCO approach such that overfitting, if present, would reduce (and not improve) model performance on the eQTL evaluation set. However, results on this evaluation set should still be viewed as a best-case scenario for pgBoost.

Results for 4,420 SNP-gene links implicated by eQTL (fine-mapped eSNP-eGene pairs with maximum PIP > 0.5 across GTEx tissues^33,34^; see Data Availability) are reported in **Figure 3a**, **Supplementary Figure 2**, and **Supplementary Table 3**. pgBoost substantially outperformed the 4 constituent scores (SCENT, Signac, ArchR, Cicero) at all distance thresholds, including >10kb (21x vs. 1.2-3.1x, all p-values for difference < 4e-35) and >100kb (35x vs. 0.89-6.6x, all p-values for difference < 0.002). pgBoost slightly outperformed the distance-based method when including all links (distance >1kb; 18x vs. 14x, p = 3e-5), but substantially outperformed it at increasingly distal thresholds, including >10kb (21x vs. 10x, p = 5e-13) and >100kb (35x vs. 3.9x, p = 1e-6). Analyses using Area Under the Precision-Recall Curve (AUPRC) produced similar findings (**Supplementary Figure 4a**).

**Figure 3.**
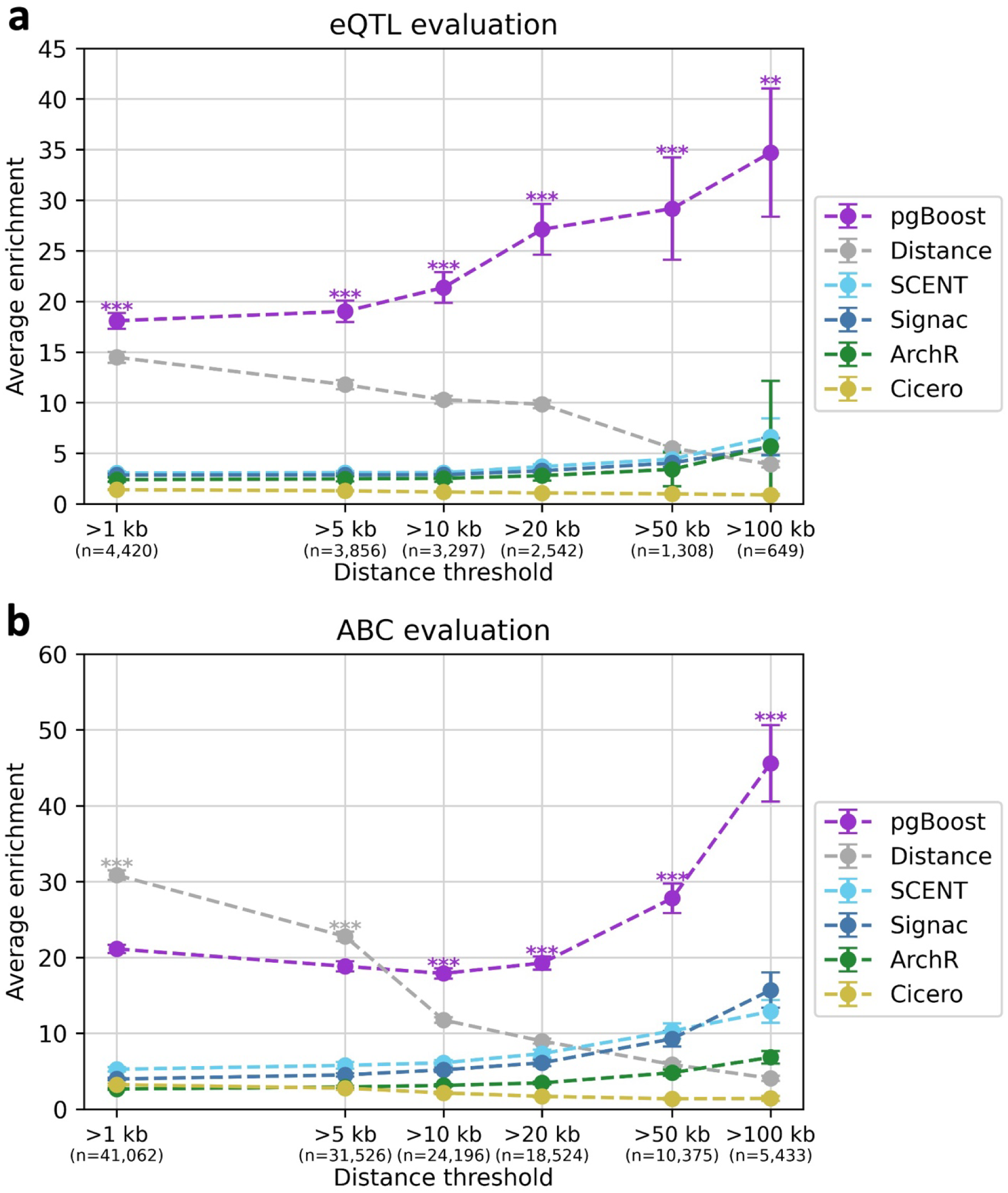
Performance of pgBoost and other methods on eQTL and ABC evaluation data sets. **a)** Average enrichment across recall values (in [0,0.35]) of links predicted by pgBoost, distance, and 4 constituent methods for 4,420 fine-mapped eSNP-eGene pairs attaining maximum PIP > 0.5 across GTEx tissues, at various distance thresholds. **b)** Average enrichment across recall values (in [0,0.25]) of links predicted by pgBoost, distance, and 4 constituent methods for 41,062 SNP-gene pairs attaining maximum ABC score > 0.2 across 344 biosamples, at various distance thresholds. The number of positive evaluation links at each distance threshold is specified in parentheses. Confidence intervals denote standard errors. Stars denote p-values for difference (*: p < 0.05, **: p < 0.01, ***: p < 0.001) of top method vs. each other method. Numerical results are reported in **Supplementary Table 3** and **Supplementary Table 4**.

Results for 41,062 SNP-gene links implicated by ABC (ABC score > 0.2 in at least one of 344 biosamples^17,35^; see Data Availability and **Supplementary Table 7**) are reported in **Figure 3b**, **Supplementary Figure 3**, and **Supplementary Table 4**. Again, pgBoost substantially outperformed the 4 constituent scores (SCENT, Signac, ArchR, Cicero) at all distance thresholds, including >10kb (18x vs. 2.1-6.1x, all p-values for difference < 6e-65) and >100kb (46x vs. 1.4-16x, all p-values for difference < 6e-9). pgBoost underperformed the distance-based method when including all links (distance >1kb; 21x vs. 31x, p = 2e-42), but substantially outperformed it at increasingly distal thresholds, including >10kb (18x vs. 12x, p = 3e-17) and >100kb (46x vs. 4.1x, p = 2e-16); we note that the ABC evaluation set strongly favors links at shorter distances (thus favoring the distance-based method), as it is derived from Hi-C contact frequencies that are largely determined by genomic distance^15,49,50^ (**Supplementary Figure 5**). Analyses using AUPRC produced similar findings (**Supplementary Figure 4b**). For eQTL and ABC evaluations, differences in performance of constituent methods were partially driven by the subset of candidate links scored by each method, particularly for SCENT (**Supplementary Figure 6a**-b, **Supplementary Table 1**, and **Supplementary Table 11**).

We investigated which features contributed most to the classification of SNP-gene links in these evaluation data sets (**Methods**). First, we compared pgBoost to restricted gradient boosting models using a subset of linking scores as features. We determined that pgBoost substantially outperformed models restricted to a single constituent score (**Supplementary Figure 7a**-b) or a single constituent score plus distance (**Supplementary Figure 8a**-b). We further determined that models ablating a single constituent score or distance generally perform slightly worse (and often significantly worse) than pgBoost, particularly when ablating distance (**Supplementary Figure 9a**-b). Second, we compared pgBoost to restricted gradient boosting models using linking scores from a subset of the 3 multiome data sets. We determined that pgBoost significantly outperformed models restricted to a single multiome data set (**Supplementary Figure 10a**) and models ablating a single multiome data set (**Supplementary Figure 10b**). Third, we conducted a SHAP^51^ analysis of feature importance. As expected, we determined that genomic distance attained the highest feature importance, followed by the “closest TSS” binary indicator, but features based on constituent scores were also broadly important (**Supplementary Figure 11**).

We performed 6 additional secondary analyses. First, we assessed how pgBoost scores vary with SNP-gene distance and determined that pgBoost scores generally decrease with distance, even at larger distances (**Supplementary Figure 12**). Second, we compared pgBoost to a simple logistic regression model using identical features and determined that gradient boosting outperforms logistic regression (**Supplementary Figure 13a**-b). Third, we compared pgBoost to an eQTL-based linking strategy (ranking candidate SNP-gene links by maximum PIP across GTEx tissues). We determined that pgBoost significantly outperformed the eQTL-based linking strategy on ABC evaluation data (**Supplementary Figure 14a**). Fourth, we compared pgBoost to an ABC-based linking strategy (ranking candidate SNP-gene links by maximum ABC score across 344 biosamples^17,35^; see **Supplementary Table 7**) and determined that pgBoost significantly outperformed ABC on eQTL evaluation data (**Supplementary Figure 15a**) (but see below). Fifth, we assessed the sensitivity of pgBoost and the 4 constituent scores to variation in cell count and sequencing depth by downsampling to 50% of the input data (**Methods**). We observed that the performance of pgBoost and the 4 constituent scores in eQTL evaluation data generally does not significantly differ between 2-fold downsampled and non-downsampled conditions (**Supplementary Figure 16**). Sixth, we assessed the calibration of the pgBoost probabilistic score. The relationship between the proportion of predicted links that were concordantly predicted by eQTL or ABC vs. the pgBoost score was monotonic but not perfectly linear, supporting the prioritization of top-scoring links rather than probabilistic scores in enrichment computations and downstream analyses (**Supplementary Figure 17**).

As more distal links are generally more difficult to detect and are of high biological interest (in particular, causal variants in disease GWAS often do not link to the closest gene^8–11^), these results indicate substantial potential for pgBoost to identify biologically important SNP-gene links.

### pgBoost improves power to identify gold-standard SNP-gene links defined by CRISPR and GWAS

We assessed whether SNP-gene links predicted by pgBoost recapitulated relatively smaller sets of gold-standard SNP-gene links that were either experimentally validated using CRISPR^15,36–42^ or implicated by fine-mapped non-coding variants near genes with fine-mapped coding variants in disease/complex trait GWAS^43,44^, providing strong evidence of biological causality. We note that the CRISPR SNP-gene links were derived from regulatory element-gene links based on SNPs that reside in a regulatory element.

Results for 571 gold-standard SNP-gene links experimentally validated in 8 CRISPR perturbation studies^15,36–42^ (**Supplementary Table 12**; see Data Availability) are reported in **Figure 4a, Supplementary Figure 18**, and **Supplementary Table 5**. pgBoost substantially outperformed the 4 constituent scores (SCENT, Signac, ArchR, Cicero) at all distance thresholds, including >10kb (4.8x vs. 1.6-2.1x, all p-values for difference < 2e-5) and >100kb (15x vs. 1.6-2.5x, all p-values for difference < 0.009). pgBoost and the distance-based method performed similarly when including all links (distance >1kb; 5.8x vs. 5.8x, p = 0.90) and at distance >10kb (4.8x vs. 4.1x, p = 0.25), but pgBoost performed substantially better at distance >100kb (15x vs. 1.6x, p = 0.008). Analyses using Area Under the Precision-Recall Curve (AUPRC) produced similar findings (**Supplementary Figure 20a**). We note that all CRISPR evaluation data was derived from the K562 erythroleukemia cell line, a cell line that is widely analyzed in CRISPR experiments^52^ (but was not directly included in the pgBoost model); while CRISPR experiments are viewed as a general gold-standard^25,35^, the relevance of these links to individual cell types will be explored below.

**Figure 4.**
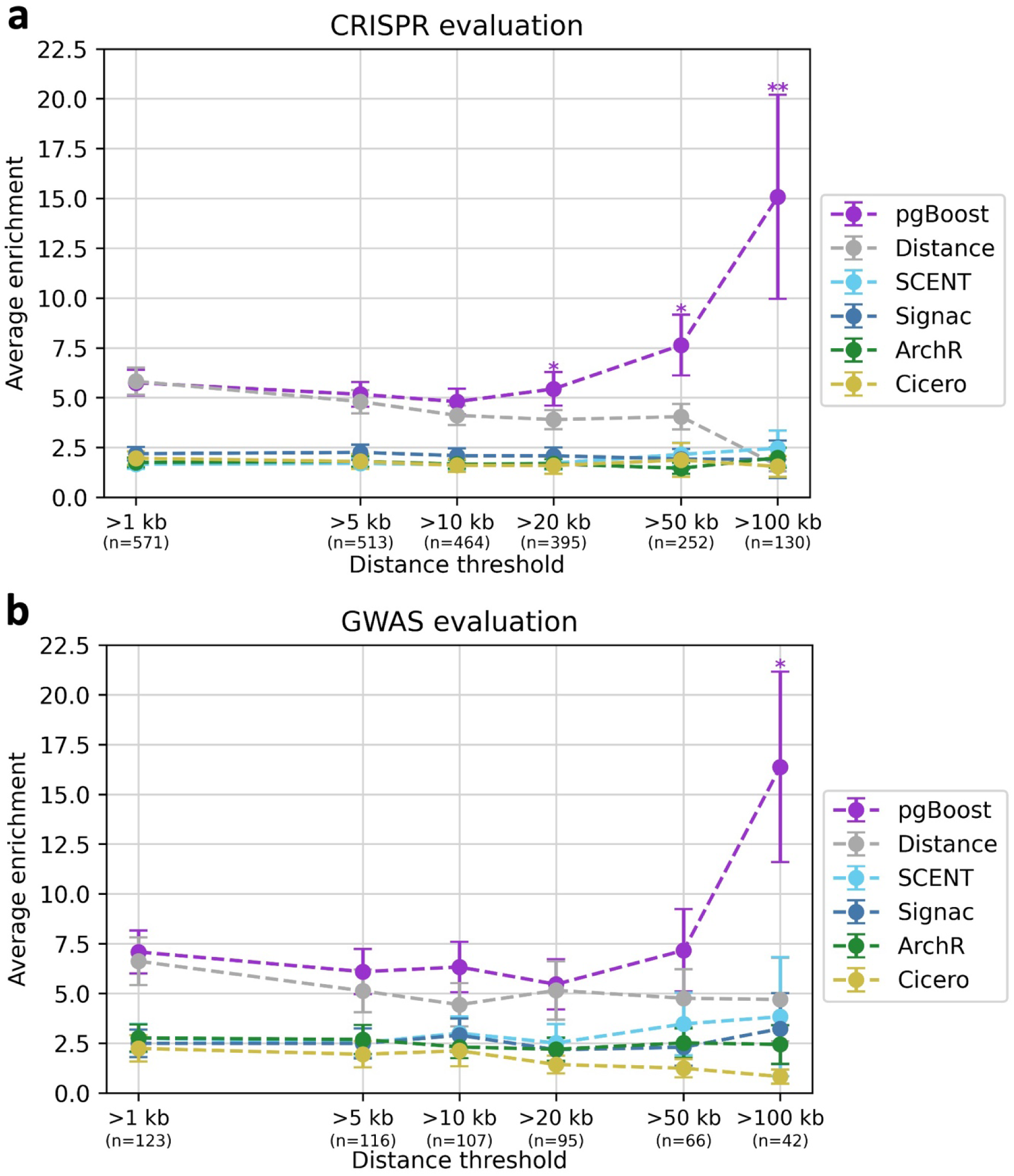
Performance of pgBoost and other methods on gold-standard evaluation data sets. **a)** Average enrichment across recall values (in [0,0.5]) of links predicted by pgBoost, distance, and 4 constituent methods for 571 links validated by CRISPR, at various distance thresholds. **b)** Average enrichment across recall values (in [0,0.25]) of links predicted by pgBoost, distance, and 4 constituent methods for 123 non-coding SNP-gene pairs derived from fine-mapped GWAS variants with a unique fine-mapped coding variant within a 2 Mb window, at various distance thresholds. The number of positive evaluation links at each distance threshold is specified in parentheses. Confidence intervals denote standard errors. Stars denote p-values for difference (*: p < 0.05, **: p < 0.01, ***: p < 0.001) of top method vs. each other method. Numerical results are reported in **Supplementary Table 5** and **Supplementary Table 6**.

Results for 123 gold-standard SNP-gene links implicated by fine-mapped non-coding variants near genes with fine-mapped coding variants^43^ in GWAS of 94 diseases/complex traits from the UK Biobank^44,53^ (PIP > 0.1 for the non-coding SNP, with a coding variant for exactly one candidate gene attaining PIP > 0.5 for the same trait in the 1 Mb window around the non-coding variant, termed “gold-standard” by ref.^10^; see Data Availability) are reported in **Figure 4b, Supplementary Figure 19**, and **Supplementary Table 6**. pgBoost substantially outperformed the 4 constituent scores (SCENT, Signac, ArchR, Cicero) at all distance thresholds, including >10kb (6.3x vs. 2.1-3.0x, all p-values for difference < 0.03) and >100kb (16x vs. 0.83-3.8x, all p-values for difference < 0.01). pgBoost and the distance-based method performed similarly when including all links (distance >1kb; 7.1x vs. 6.6x, p = 0.62) and at distance >10kb (6.3x vs. 4.4x, p = 0.06), but pgBoost performed substantially better at distance >100kb (16x vs. 4.7x, p = 0.02). Analyses using AUPRC produced similar findings (**Supplementary Figure 20b**). For CRISPR and GWAS evaluations, differences in performance of constituent methods were partially driven by the subset of candidate links scored by each method (but less so than for eQTL and ABC evaluations) (**Supplementary Figure 6c**-d, **Supplementary Table 1**, and **Supplementary Table 11**).

We evaluated the performance of pgBoost trained on other training data sets (e.g. CRISPR or ABC) instead of eQTL, using the same set of input features; as noted above, each link set that we analyze can be considered as a prediction, used for model training, or used for evaluation. First, we trained pgBoost using CRISPR links. We determined that training on eQTL significantly outperformed training on CRISPR for eQTL and ABC evaluations (**Supplementary Figure 21a-b**). While other regulatory linking methods have used CRISPR links for model training^15,35^, pgBoost may perform better when trained on eQTL due to the limited amount of CRISPR data, pervasive aneuploidy and structural variants in the K562 cell line^52^, and/or better representation of multiple cell types in GTEx eQTL data. However, training on CRISPR vs. eQTL yielded similar performance on CRISPR and GWAS evaluations, motivating further research on training using CRISPR data (**Supplementary Figure 21c**-d). Second, we trained pgBoost using ABC links (ABC score > 0.2 or > 0.4 in at least one of 344 biosamples^17,35^). Training using eQTL and ABC links attained the highest enrichments for eQTL and ABC evaluations, respectively, but training on eQTL links significantly outperformed training on ABC links for CRISPR evaluation, with a concordant but non-significant result for GWAS evaluation (**Supplementary Figure 22**). Third, we trained pgBoost using fine-mapped eSNP-eGene pairs in whole blood (the GTEx tissue closest to the single-cell data) instead of all GTEx tissues. Training on eSNP-eGene pairs from all GTEx tissues significantly outperformed training on blood eSNP-eGene pairs for eQTL, ABC, CRISPR, and GWAS evaluations (**Supplementary Figure 23**). Fourth, we trained pgBoost using fine-mapped eSNP-eGene pairs at other PIP thresholds, instead of PIP > 0.5. We determined that more stringent PIP thresholds yielded smaller training data sets and significantly lower pgBoost enrichments for eQTL, ABC, CRISPR, and GWAS evaluations (**Supplementary Figure 24**). We did not train pgBoost using the set of gold-standard SNP-gene links identified by GWAS, as this set contained only 123 links.

We performed secondary analyses computing CRISPR evaluation and GWAS evaluation enrichments attained by gradient boosting models restricted to subsets of features (**Supplementary Figure 7c**-d, **Supplementary Figure 8c**-d, and **Supplementary Figure 9c**-d) and a logistic regression model (**Supplementary Figure 10c**-d), and reached similar conclusions (when well-powered to detect a difference) as in eQTL and ABC evaluations (see above). We also compared the performance of pgBoost to eQTL and ABC-based linking strategies (see above) on CRISPR and GWAS evaluation data. pgBoost performed similarly to eQTL on CRISPR evaluation data (significantly outperforming for longer-range links), with a concordant but non-significant result for GWAS evaluation (**Supplementary Figure 14b**-c). pgBoost performed similarly to ABC on CRISPR and GWAS evaluations (**Supplementary Figure 15b**-c).

These results indicate substantial power for pgBoost to detect experimentally validated links and links underlying disease/complex trait GWAS associations. In particular, pgBoost strongly outperforms the 4 constituent scores and distance-based method in capturing longer-range links.

### Identifying and evaluating SNP-gene links in a focal cell type

pgBoost leverages features from an input set of data set-cell type pairs, which may often span multiple cell types (**Figure 2**, **Table 2**). However, in settings where the goal is to predict SNP-gene links in a focal cell type, it may be beneficial to restrict model features to that cell type. We considered two different types of evaluation involving a focal cell type: *cell-type-specific* evaluation, involving evaluation sets of SNP-gene links that are present in the focal cell type and absent from other cell types, and *cell-type-level* evaluation, involving evaluation sets of SNP-gene links that are present in the focal cell type (irrespective of presence/absence in other cell types). We generated independent pgBoost predictions for each of the 4 major cell types of the Luecken BMMC^48^ multiome data set (spanning > 8,000 cells: T cell, B cell, myeloid, erythroid; **Table 2** and **Supplementary Table 2**) by restricting model features to each cell type in turn (as well as the 2 distance-based features; **Methods**) during model training and prediction. We restricted most analyses to the Luecken BMMC^48^ data to minimize the impact of data set-specific experimental factors. We continued to use fine-mapped eSNP-eGene pairs^33,34^ (maximum PIP > 0.5 vs. maximum PIP < 0.01 across GTEx tissues) as training data. We compared these pgBoost predictions (as well as a pgBoost prediction using model features from all 4 major cell types) using both cell-type-specific and cell-type-level evaluation data. Given the key role of distance-based features (**Figure 3**, **Figure 4**) and previous work showing that distal/enhancer-driven architectures are more cell-type-specific than proximal/promoter-driven architectures^3,18–20,54^, we primarily focused these analyses on relatively distal candidate SNP-gene links with distance >10kb; we consider other distance thresholds in secondary analyses.

We first analyzed cell-type-specific ABC evaluation data, defined by SNP-gene links strongly implicated in biosample(s) related to the focal cell type (ABC score > 0.2) and not strongly implicated in other biosamples (ABC score < 0.1; **Supplementary Table 7** and **Methods**). Across all 4 cell-type-specific ABC evaluation sets, the pgBoost model restricted to features from the focal cell type significantly outperformed other pgBoost models, including models restricted to other cell types and the model spanning all 4 cell types (**Figure 5a**, **Supplementary Figure 25**, and **Supplementary Table 8**); results were similar at other distance thresholds (>1kb to >100kb, instead of >10kb) (**Supplementary Figure 26**).

**Figure 5.**
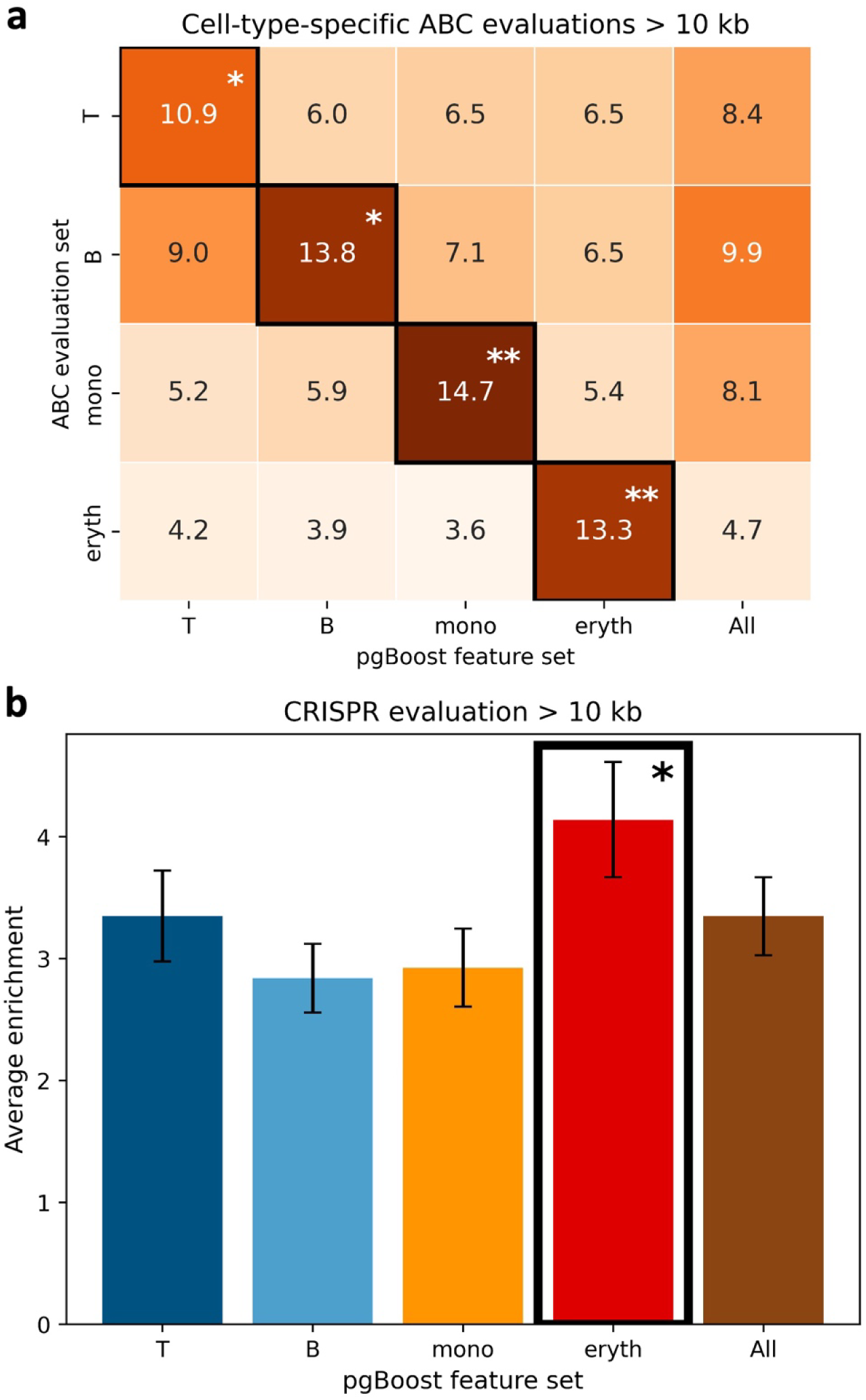
Performance of pgBoost restricted to features from a focal or non-focal cell type in evaluation data sets relevant to the focal cell type. **a)** Average enrichment across recall values (in [0,1]) of links predicted by pgBoost restricted to features from a focal cell type for training and prediction (columns; All denotes all features) for cell-type-specific ABC evaluation links (rows), at distance >10kb (number of evaluation links in each cell type: n_T_ = 1,732, n_B_ = 1,150, n_mono_ = 412, n_eryth_ = 115). **b)** Average enrichment across recall values (in [0,1]) of links predicted by pgBoost restricted to features from a focal cell type (columns; All denotes all features) for cell-type-level CRISPR evaluation links in K562, at distance >10kb (number of evaluation links: n = 461). Confidence intervals denote standard errors. Stars denote p-values for difference (*: p < 0.05, **: p < 0.01, ***: p < 0.001) of top method (defined separately for each row in **a)**; denoted in black outline) vs. each other method. Numerical results are reported in **Supplementary Table 8** and **Supplementary Table 9**.

We next analyzed cell-type-level CRISPR evaluation data, defined by SNP-gene links experimentally validated in the K562 cell line, an erythroleukemia cell line widely used to study erythroid differentiation and red blood cell biology^55–57^. The pgBoost model restricted to features from erythroid cells significantly outperformed other pgBoost models, including models restricted to other cell types and the model spanning all 4 cell types (**Figure 5b**, **Supplementary Figure 27**, and **Supplementary Table 9**); results were similar at other distance thresholds (> 1kb to >100kb, instead of >10kb) (**Supplementary Figure 28**). We did not consider cell-type-specific evaluation, because all CRISPR evaluation data was derived from K562 (a widely analyzed cell line in CRISPR experiments^52^).

We also analyzed cell-type-level GWAS variant evaluation data (evaluating SNPs linked to genes, instead of evaluating SNP-gene links), defined by fine-mapped variants (PIP > 0.2; **Methods**) for 7 red blood cell or platelet-related blood cell traits (for which erythroid cells are most relevant) and 7 autoimmune diseases and granulocyte-related blood cell traits (for which T cells, B cells and monocytes are most relevant^58^). (We did not consider SNP-gene links implicated by fine-mapped non-coding and coding variants in GWAS^43,44^ (**Figure 4b**) restricted to these sets of traits, as this would result in prohibitively small evaluation sets.) For the first set of traits, pgBoost models restricted to erythroid cells significantly outperformed other pgBoost models (**Supplementary Figure 29a**). For the second set of traits, pgBoost models restricted to monocytes significantly outperformed other cell-type-level pgBoost models and non-significantly outperformed a pgBoost model spanning all 4 cell types (**Supplementary Figure 29b**). We also analyzed cell-type-level GTEx evaluation data, defined by fine-mapped eSNP-eGene pairs identified in EBV-transformed lymphocytes or LCL (PIP > 0.2); we expanded the set of pgBoost models to include a model restricted to LCL features from the SHARE-seq LCL^22^ data set (**Table 2**). The pgBoost model restricted to LCL and the model spanning all 5 cell types non-significantly outperformed other models (**Supplementary Figure 30**).

We performed 4 additional secondary analyses. First, instead of restricting features to cell type X for training and prediction and evaluating in cell type Y in cell-type-specific ABC evaluation data (**Figure 5a**), we restricted features to cell type X for training, used features in cell type Y for prediction, and evaluated in cell type Y in cell-type-specific ABC evaluation data. We determined that the cell type used for training does not significantly impact performance (**Supplementary Figure 31**), suggesting that the cell-type-specific patterns of enrichment in **Figure 5a** were driven by the use of the focal cell type for prediction rather than the use of the focal cell type for training. Second, we generated independent pgBoost predictions for each of the 3 major cell types shared by the Luecken BMMC^48^ and 10X PBMC^47^ multiome data sets (T cell, B cell, myeloid; **Table 2** and **Supplementary Table 2**) by restricting model features to linking scores from both data sets for each cell type (as well as the 2 distance-based features). Results in cell-type-specific ABC evaluation data were qualitatively similar to **Figure 5a** but less statistically significant (**Supplementary Figure 32**). Third, we analyzed cell-type-specific evaluation data defined by SNP-gene links identified in biosample(s) related to the focal cell type (ABC score > 0.2) but below a more stringent threshold in non-focal cell types (ABC score < 0.05), and observed qualitatively similar patterns (**Supplementary Figure 33**). Fourth, we analyzed cell-type-level ABC evaluation data, defined by SNP-gene links identified in biosample(s) related to the focal cell type (ABC score > 0.2), irrespective of other biosamples. Models restricted to the focal cell type generally significantly outperformed models restricted to other cell types but attained similar performance as the model spanning all 4 cell types (**Supplementary Figure 34**).

We conclude that training pgBoost by restricting to features from a focal cell type generally maximizes power to detect SNP-gene links corresponding to that cell type in cell-type-specific and cell-type-level evaluation data—despite the inclusion of fine-mapped eSNP-eGene pairs from all GTEx tissues in our training data (see **Supplementary Figure 23**). However, training pgBoost using all cell types attained similar performance as training pgBoost in the focal cell type in some cell-type-level evaluation experiments (**Supplementary Figure 30** and **Supplementary Figure 34**).

### pgBoost links variants to genes mediating GWAS associations

We have shown that SNP-gene links identified by pgBoost may not be identified by other methods (**Figure 3** and **Figure 4**). Below, we dissect four loci in which pgBoost links fine-mapped GWAS variants to experimentally validated or biologically plausible target genes that are not identified by other single-cell methods; for simplicity, we defined SNP-gene links identified by a given method based on the top 5% of linking scores across all candidate links at distance >1kb (**Supplementary Table 11**; see **Discussion**). The four examples were derived from fine-mapping results for 94 UK Biobank diseases/complex traits^44,53^: 9,732 candidate SNP-gene links, spanning 852 variants with PIP > 0.5 for at least one trait and 5,577 genes; 625 SNP-gene links in the top 5% for pgBoost, reflecting enrichment for pgBoost links that link fine-mapped GWAS variants (**Supplementary Table 11**).

First, pgBoost linked rs1427407 (fine-mapped for red blood cell count with PIP = 0.99) to *BCL11A* (40kb distance to TSS; closest TSS) (**Figure 6a**). rs1427407 lies in an experimentally validated intronic enhancer of *BCL11A*. *BCL11A* is known to play a critical role in fetal-to-adult hemoglobin switching in erythroid cells^59,60^, and the disruption of this enhancer has been shown to result in decreased *BCL11A* expression and elevated fetal hemoglobin^61^. This link also scored in the top 5% of SCENT links (although it failed to pass the established SCENT threshold of FDR < 0.1^25^), but scored outside the top 5% of links predicted by other single-cell methods and the distance-based method. We also examined cell-type-level pgBoost predictions and observed that this link is scored highly by pgBoost_eryth_ (99.6% percentile) but not pgBoost_T/B/mono_ (89.2-92.6% percentile), consistent with experimental evidence that activity of the focal enhancer is necessary for *BCL11A* expression in erythroid cells but not in other cell types^62^.

**Figure 6.**
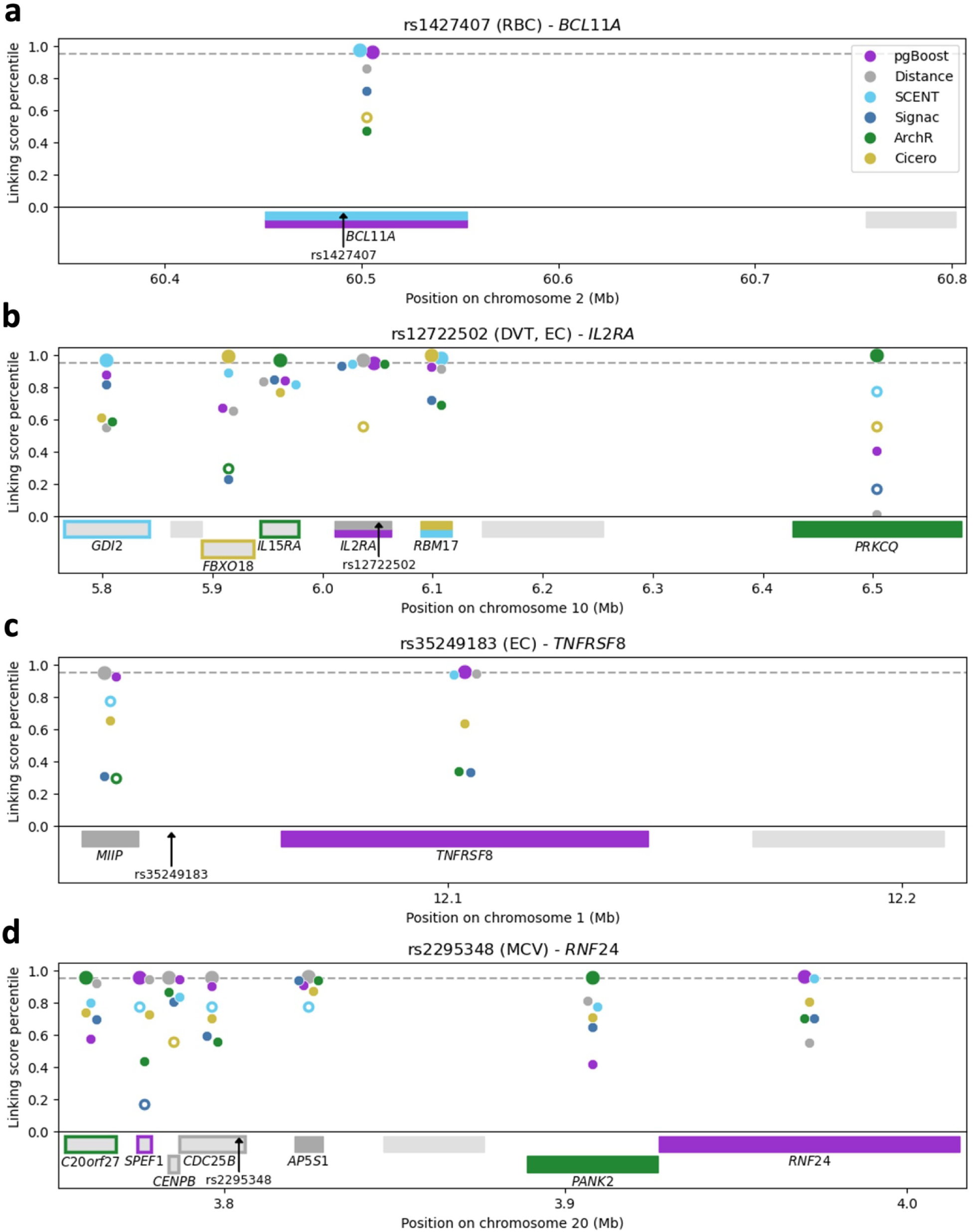
Examples of SNP-gene links identified by pgBoost. We report links prioritized by pgBoost, distance, and 4 constituent methods for 4 fine-mapped GWAS variants: **(a)** rs1427407, **(b)** rs11864973, **(c)** rs35249183, and **(d)** rs2295348. Figure panel headings for each SNP report the corresponding GWAS trait(s)^44,53^ (in parentheses) and the SNP-gene link attaining the highest pgBoost score. Linking scores for each method are reported as dots in the middle of each gene (jittered for easier visualization). Large dots denote links scored in the top 5% by a given method. Hollow dots denote links that were not scored by a given method (with their percentile defined as the percentage of links not scored by the method). Genes scored in the top 5% by any method are reported at the bottom of each panel (with gene name); nearby genes are also displayed (without gene name). Colored boxes denote the gene attaining the highest linking score percentile > 95% by one or more method(s); colored borders denote other genes scored in the top 5% by a given method. Numerical results are reported in **Supplementary Table 13.** RBC: red blood cell count, DVT: deep vein thrombosis, EC: eosinophil count, MCV: mean corpuscular volume.

Second, pgBoost linked rs12722502 (fine-mapped for both deep vein thrombosis with PIP = 0.93 and eosinophil count with PIP = 0.51) to *IL2RA* (11kb distance to TSS; closest TSS) (**Figure 6b**). rs12722502 lies in an intronic enhancer of *IL2RA* experimentally validated by CRISPR activation^63^. *IL2RA* encodes a key component of the interleukin 2 signaling pathway critical for T cell proliferation and homeostasis^64,65^ and has been implicated in rheumatoid arthritis^66^, multiple sclerosis^67^, systemic sclerosis^68^, systemic lupus^69^, and type 1 diabetes^70^. This link also scored in the top 5% of links predicted by the distance-based method, whereas existing single-cell methods assigned top priority to other genes (*RBM17, PRKCQ*) and additionally scored other genes in the top 5% of links (*GDI2*, *FBXO18*, *IL15RA*).

Third, pgBoost linked rs35249183 (fine-mapped for eosinophil count with PIP = 1.00) to *TNFRSF8* (also known as *CD30*) (24kb distance to TSS; second closest TSS) (**Figure 6c**). rs35249183 lies in an ENCODE cCRE with a distal enhancer signature^71^. This is considered a gold-standard SNP-gene link, based on an independent *TNFRSF8* GWAS coding variant that was fine-mapped for eosinophil count^43,44^. *TNFRSF8* has been widely studied for its role in NF-*κ*B transcription and lymphocyte growth and transformation, in particular for its involvement in lymphoblastic malignancies^72–75^, and is thus a biologically plausible target gene. This link scored outside of the top 5% of links predicted by other single-cell methods as well as the distance-based method, which implicated a different gene (*MIIP*).

Fourth, pgBoost linked rs2295348 (fine-mapped for mean corpuscular volume with PIP = 0.58) to *RNF24* (210kb distance to TSS; 11th closest TSS) (**Figure 6d**). rs2295348 lies in an ENCODE cCRE with a distal enhancer signature^71^, within an intron of *CDC25B*. *RNF24* encodes an integral membrane protein of the Golgi apparatus^76^. While the candidate link between rs2295348 and *RNF24* has not been experimentally tested (to our knowledge), knockout of *RNF24* resulted in decreased hemoglobin content and erythrocyte cell number in mice^77,78^, suggesting that *RNF24* is a biologically plausible target gene. The disruption of *CDC25B*, which encodes a phosphatase driving cell division, results in no apparent phenotypes besides female sterility^79,80^, and compensatory mechanisms among *CDC25A/CDC25B/CDC25C* require triple knockout to elicit phenotypic effects^81,82^. rs2295348-*RNF24* scored outside of the top 5% of links predicted by other single-cell methods and the distance-based method, which assigned top priority or top 5% of linking scores to other genes: *C20orf27*, *CENPB*, *AP5S1*, *PANK2*, and *CDC25B* (for which knockout mouse models identified no relevant phenotypic effects^77–80^). We note that one other link, rs2295348-*SPEF1* (distance 27kb), also scored in the top 5% of pgBoost links; the knockout of *SPEF1* results in no relevant phenotypic effects in mice^81,82^, demonstrating that thresholding at the top 5% of links is not expected to attain perfect specificity and may implicate false-positive links (see **Discussion**). We further note that rs2295348-*RNF24* also scored in the top 0.2% of pgBoost links at distance >100kb.

These examples demonstrate that pgBoost can combine weak or conflicting evidence from constituent methods to link regulatory variants to experimentally validated or biologically plausible target genes.

## Discussion

We developed a gradient boosting^32^ framework (pgBoost) integrating single-cell peak-gene linking scores^25–28^ with genomic distance by training on fine-mapped eSNP-eGene pairs^33,34^ to predict the causal probability of candidate SNP-gene links. We applied pgBoost and existing peak-gene linking methods to 3 scRNA/ATAC-seq multiome data sets^22,47,48^ spanning 6 cell types and 85K cells. We demonstrated that pgBoost significantly outperforms constituent single-cell methods and genomic distance in enrichment for SNP-gene links derived from eQTL^33,34^, Activity-By-Contact (ABC)^17,35^, CRISPRi^15,36–42^, and GWAS^43,44^ data. In particular, pgBoost substantially outperformed existing methods in evaluations of longer-range links, which are of high biological importance (for example, non-coding variants in GWAS often do not regulate the closest gene^8–11^) and are more difficult to capture using distance-based approaches (e.g. linking SNPs to genes within genomic windows^83–85^). We have not conclusively established whether longer-range links reflect *cis*-regulatory or *trans*-regulatory biology, but we hypothesize that they are predominantly *cis*-regulatory, as pgBoost scores generally decrease with distance, even at larger distances (**Supplementary Figure 12**). We further demonstrated that restricting pgBoost to features from a focal cell type improves power to detect regulatory links relevant to that cell type, supporting the future application of this framework to a wider variety of biological contexts. Finally, we applied pgBoost scores to fine-mapping results for 94 UK Biobank diseases/traits^44,53^ to nominate target genes mediating GWAS associations, highlighting several examples where pgBoost links a fine-mapped variant to an experimentally validated or biologically plausible target gene.

Our work has several implications for downstream analyses. First, we have publicly released pgBoost scores as a resource to aid in the functional interpretation of GWAS variant-disease associations or epigenomic assays. While we highlight links scoring in the top 5% by pgBoost score, we note that this is a stringent cutoff at larger distances (for example, only 0.3% of links >100kb are included in the top 5% of all links); thus, this threshold could plausibly be relaxed for longer-range links. Second, pgBoost scores nominate SNP-gene pairs with high causal probability for functional follow-up (e.g. CRISPRi^15,36–42^ or base editing^86^) to validate predicted links and dissect underlying biological mechanisms. Third, we have publicly released open-source software enabling users to retrain pgBoost while incorporating features from additional scRNA/ATAC-seq multiome data sets^87–89^ (or additional single-cell methods), which is likely to improve pgBoost predictions (**Supplementary Figure 10**); in particular, as well-powered non-immune data sets become available, incorporating these data sets can broaden the scope of pgBoost applications to non-immune-related diseases and functions. We note that we have performed our analyses at the level of SNP-gene links, but users can also generate pgBoost predictions for peak-gene links by computing constituent peak-gene linking scores using peaks that are merged across data sets and defining positive and negative training sets of peak-gene links based on the maximum PIP (of fine-mapped eSNP-eGene pairs) across SNPs within a focal peak. Fourth, the eQTL, ABC, CRISPR and GWAS evaluation sets of SNP-gene links that we have used here may prove useful in future studies.

Our work has several limitations and directions for future research. First, because pgBoost uses as features linking scores computed from ATAC peaks, predictions are largely limited to SNPs lying in regions of accessible chromatin. However, chromatin accessibility is a known predictor of regulatory activity, making ATAC peaks a well-supported prior for regulatory regions^14,16,17^. In addition, it would be straightforward to apply pgBoost using a genomic tiling approach to define candidate regulatory regions instead of ATAC peaks (although we anticipate limited benefit from this approach). Second, we focused our analyses on immune/blood cell types (for which there currently exist multiple high-quality multiome data sets^22,47,48^), but it will be important to apply pgBoost to other tissues/cell types as more multiome data becomes available. Third, pgBoost may fail to identify links specific to rare cell types or disease contexts. Encouragingly, pgBoost identifies links specific to major immune/blood cell types when including fine-mapped eSNP-eGene pairs from all GTEx tissues in model training. However, it will be important to investigate pgBoost performance in rare contexts as more multiome data from such contexts becomes available. Fourth, pgBoost does not exhaustively include all single-cell peak-gene linking methods^25–31^. However, our analyses suggest that there will be diminishing returns from including additional single-cell peak-gene linking methods (**Supplementary Figure 9**); indeed, although our primary recommendation is to include 4 peak-gene linking methods^25–28^, a convenient and plausible alternative is to ablate ArchR^27^, which requires separate processing of raw fragment/BAM files. Fifth, pgBoost does not include as features linking scores derived from bulk data sets (e.g. ABC, evaluated in **Supplementary Figure 15**) or enhancer-associated biochemical and epigenomic annotations^14,90–94;^ including these features in future extensions of pgBoost may further improve performance. Sixth, we have not conclusively established whether it is better to use eQTL data or CRISPR data for training (considering comparable performance on GWAS-derived evaluation data). It will be of interest to further explore training on CRISPR-validated links as more CRISPR data sets become available. Seventh, a high pgBoost score (for example, in the top 5% of all links) does not imply biological causality, underscoring the importance of experimental validation^15,36–42,61,63^. Despite these limitations, we have shown that pgBoost improves power to link regulatory variants to genes and can aid the functional interpretation of GWAS variant-disease associations.

## Code Availability

The code to implement pgBoost has been made publicly available at https://github.com/elizabet hdorans/pgBoost.

## Data Availability

All single-cell multiome data sets analyzed are publicly available. The 10x PBMC data set is available at https://www.10xgenomics.com/datasets. The Luecken BMMC and SHARE-Seq LCL data sets are available at Gene Expression Omnibus (accession codes <u>GSE194122</u> and GSE140203, respectively).

Linking scores and percentiles for pgBoost and constituent methods have been made publicly available at 10.5281/zenodo.11211926.

Fine-mapped eQTL data from ref.^34^ are available at https://www.finucanelab.org/data.

ABC scores are available on the ENCODE portal (https://www.encodeproject.org/).

Biosample IDs and file accessions are listed in **Supplementary Table 7**.

The CRISPR data set is available at https://github.com/EngreitzLab/CRISPR_comparison/blob/ main/resources/crispr_data/EPCrisprBenchmark_ensemble_data_GRCh38.tsv.gz.

GWAS-derived SNP-gene links from ref.^43^ are available at https://github.com/Deylab999/GWAS_benchmark_IGVF/blob/main/UKBiobank.ABCGene.anyabc.tsv.

GWAS fine-mapping results from ref.^44^ are available at https://www.finucanelab.org/data.

## Acknowledgements

We thank Jesse Engreitz, Robin Andersson, Andreas Gschwind, and Weilin Qiu for valuable feedback on the manuscript and Soumya Raychaudhuri, Shamil Sunyaev, Vijay Sankaran, Saori Sakaue, Carles Boix, Ben Strober, Jordan Rossen, and Martin Zhang for helpful discussions. This research was funded by NIH grants U01 HG012009, R56 HG013083, R01 MH101244, R37 MH107649, R01 HG006399, and R01 MH115676.

## Methods

### Existing single-cell peak-gene linking methods

We applied 4 existing single-cell peak-gene linking methods (SCENT^25^, Signac^26^, ArchR^27^, Cicero^28^; **Table 1**) separately to each cell type in each data set (**Table 2** and **Supplementary Table 2**), generating 9 total sets of predictions from each method (1 for each data set-cell type pair). We selected candidate links as peak-gene pairs with an ATAC-seq peak centered ±500kb from the gene’s TSS (as in refs.^25,26,28^) and not overlapping any promoter region (defined as ± 1kb from the gene’s TSS as in ref.^95^). Some methods implement additional requirements for candidate peak-gene pairs to test for association (for example, SCENT only tests peak-gene pairs from which the peak and gene are both active in ≥ 5% of cells; **Supplementary Table 1**). To integrate linking scores across data sets and facilitate downstream variant-level analyses (e.g. interpreting GWAS variant disease associations), we intersected peaks with 1000 Genomes Project variants with minor allele count ≥ 5^10,45,46^ to define SNP-gene linking scores from peak-gene linking scores.

Brief descriptions and implementation details of each linking method are provided below: SCENT^25^ uses Poisson regression to test the association of each candidate peak-gene pair (while controlling for batch and cell-level covariates) and quantifies the significance of associations by bootstrapping across cells. We ran SCENT on gene expression count matrices and binarized ATAC peak matrices using the SCENT_algorithm function, specifying number of unique molecular identifiers (UMIs) and percentage of mitochondrial reads as cell-level covariates. We then computed FDR (Benjamini Hochberg correction) from bootstrapped p-values as in ref.^25^

Signac^26^ computes the correlation between binarized peak accessibility and normalized gene expression across single cells for each candidate peak-gene pair and quantifies significance by permuting across off-chromosome peaks matched to the focal peak for peak accessibility, peak length, and GC content. After normalizing and scaling gene expression data using the Seurat SCTransform function^96^, we ran Signac on the normalized gene expression and binarized ATAC peak matrices using the LinkPeaks function with a p-value cutoff set to 1 and a score cutoff set to 0.

ArchR^27^ defines metacells (low-level cell aggregates) using *k*-nearest neighbors, aggregates RNA and ATAC peak counts across single cells in each metacell, and computes the correlation between normalized aggregated RNA and ATAC peak counts across metacells. ArchR outputs an FDR computed from the analytical correlation p-value. We ran ArchR using the addPeak2GeneLinks function and accessed peak-gene links using the getPeak2GeneLinks function with FDR cutoff set to 1.

Cicero^28^ (which uses as input scATAC-seq data only) computes partial correlations across metacells between pairs of ATAC peaks, quantifying the association between two peaks after accounting for accessibility of all other peaks ± 500kb. Cicero uses the Graphical LASSO^97^ with a distance-dependent penalty term to prioritize local *cis*-regulatory interactions. Our results show that this penalty term does not effectively capture the impact of genomic distance (**Figure 3** and **Figure 4**). These peak-peak associations can be interpreted as peak-gene associations when one peak overlaps a gene promoter (we define the promoter region as ± 1kb from the gene’s TSS as in ref.^95^). We ran Cicero on binarized ATAC peak matrices using the run_cicero function.

### pgBoost framework

pgBoost is a gradient boosting classification model (implemented using XGBoost software^32^) which takes as input (features) linking scores from existing methods and genomic distance and outputs (predictions) probabilistic scores for each pair.

The gradient boosting algorithm iteratively builds regression trees to capture relationships between features *x_i_* and observed “truth” values *y_i_*, minimizing the following objective function at each iteration *t*:

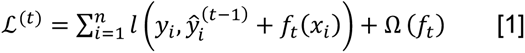

where *l*(*y_i_*, *ŷ_i_*)i s a loss function measuring the difference between “truth” (*y_i_*) and prediction (*y_i_*) for instance *i*, *f_t_*(*x_i_*) is a regression tree fit to the errors from the previous iteration, and Ω(*f_t_*) is a regularization term which prevents the model from overfitting to the training data. The model predicts the causal probability of candidate links by summing over tree outputs: 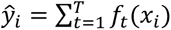.

Across candidate SNP-gene links, pgBoost considers as features for each cell subset (one cell type in one data set): SCENT regression effect size and bootstrapped FDR, Signac correlation and permutation p-value, ArchR correlation and FDR, Cicero peak-promoter co-accessibility, four binary indicators (1 or 0) based on whether a candidate link is scored by each of these methods, SNP-TSS distance, and a binary indicator for “closest TSS.” A feature matrix of 4,503,999 candidate SNP-gene links x 101 features was thus defined for candidate SNP-gene links profiled in any of the 3 multiome data sets (**Table 2** and **Supplementary Table 2**). If a candidate link was not scored by a given method in a given cell subset, a 0 was entered as the corresponding score feature (SCENT effect size, Signac correlation, ArchR correlation, Cicero co-accessibility), and a 1 was entered as the corresponding p-value/FDR feature, if applicable (SCENT FDR, Signac p-value, ArchR FDR).

We defined a training set comprised of fine-mapped eSNP-eGene pairs attaining a posterior inclusion probability (PIP) > 0.5 in any GTEx tissue as positives and PIP < 0.01 in all GTEx tissues as negatives^33,34^. We intersected this set of SNP-gene pairs with candidate SNP-gene links profiled in any of the 3 multiome data sets (**Table 2** and **Supplementary Table 2**) and removed any “duplicate” SNP-gene pairs receiving the same scores across data set-cell type pairs (intuitively, limiting to 1 entry per uniquely-defined regulatory element). We also “gene-matched” negative and positive SNP-gene pairs such that all genes in the set of negative SNP-gene pairs are also included in the set of positive SNP-gene pairs. In total, pgBoost was trained on 4,420 positive SNP-gene pairs and 65,114 negative SNP-gene pairs.

We specified the following model parameters (passed to the xgboost^32^ function): 1000 iterations, maximum tree depth (10, 15, 25), learning rate (0.05), gamma (10), minimum child weight (6, 8, 10), and subsample (0.6, 0.8, 1). Gamma and learning rate were selected in accordance with previous studies^98–100^ to minimize overfitting. Minimum child weight, subsample, and number of iterations were also specified in accordance with refs.^100,101^

### Single-cell data sets

We downloaded 3 publicly available scRNA/ATAC-seq multiome data sets (10X PBMC^47^, Luecken BMMC^48^, and SHARE-seq^22^; **Table 2** and **Supplementary Table 2**) (see Data Availability). We obtained publicly available QCed gene expression and ATAC peak matrices for the Luecken BMMC data set^48^ and processed the 10x PBMC^47^ and SHARE-seq^22^ data sets using both Seurat 4.3.0.1^102^ and ArchR 1.0.2^27^ (ArchR requires raw fragment/BAM files as input and thus separate processing). We first selected cells from each data set passing QC filters on total ATAC and RNA reads, nucleosome signal, and TSS enrichment (**Supplementary Table 10**) in accordance with ref.^25^ We next identified “peaks” of open chromatin in each data set using macs2^103^ and excluded non-standard chromosomes and blacklist regions. To ensure uniform cell and peak sets were used by all methods, we retained cells that passed QC filters (**Supplementary Table 10**) in both Seurat and ArchR and imported the set of ATAC peaks into ArchR using the addPeakSet function.

These steps yielded matrices of gene expression and ATAC peak counts in individual cells for each data set. We also generated binarized ATAC peak matrices, in which an entry of 1 for a given peak-cell pair indicates that ≥ 1 ATAC fragments overlap the peak and an entry of 0 indicates that no ATAC fragments overlap the peak. Finally, we annotated cells with cell types, using cell type labels provided by the authors for the Luecken BMMC^48^ and SHARE-seq^22^ data sets and performing reference-query mapping with a PBMC reference object in Seurat to define cell type labels in the 10X PBMC^47^ data set. We restricted all analyses to the set of genes analyzed in ref.^35^

### Defining single-cell and distance-based SNP-gene linking scores

Running existing peak-gene linking methods separately in each of the 9 data set-cell type pairs (**Table 2** and Supplementary **Table 2**) typically yielded multiple scores per method for a single candidate SNP-gene link. To facilitate downstream analyses, we assigned a single score per method to each candidate SNP-gene link in accordance with ref.^25^ For SCENT, we assigned each candidate SNP-gene link the minimum FDR observed across cell subsets. For Signac, we assigned each candidate SNP-gene link the maximum |score| associated with p-value < 0.05 (default parameter in Signac software described by ref.^26^) observed across cell subsets. For ArchR, we assigned each candidate SNP-gene link the maximum |correlation| associated with FDR < 0.1 (in accordance with ref.^25^) across cell subsets. For Cicero, we assigned each candidate SNP-gene link the maximum |co-accessibility| observed across cell subsets.

To compare the performance of pgBoost and existing methods to a distance-based strategy, we defined a distance-based linking score by ranking all candidate SNP-gene links by SNP-TSS distance (such that more proximal SNP-gene links are assigned higher linking scores).

### Evaluation data sets

We defined 4 main evaluation sets derived from eQTL^33,34^, Activity-By-Contact (ABC)^17,35^, CRISPR^15,36–42^, and GWAS^43,44^ (see Data Availability). We intersected all evaluation sets with candidate SNP-gene links profiled in at least one single-cell multiome data set analyzed (involving a SNP lying in an ATAC-seq peak and a gene expressed in the focal data set; **Table 2**).

Brief descriptions of each evaluation set are provided below:

The eQTL evaluation set is comprised of 4,420 fine-mapped eSNP-eGene pairs attaining a posterior inclusion probability (PIP) > 0.5 in any GTEx tissue^33,34^. Fine-mapping was conducted by ref.^34^ (by applying the SuSiE^104^ fine-mapping method to 49 GTEx tissues^33^).

The Activity-By-Contact evaluation set is comprised of 41,062 SNP-gene links attaining ABC score > 0.2 in any of 344 biosamples passing QC^17,35^. Activity-By-Contact scores^17^ were computed and made publicly available by ref.^35^ (344 biosamples were obtained by removing duplicate assays corresponding to same cell type from the 352 biosamples released by ref.^35^). A full list of biosamples analyzed is reported in **Supplementary Table 7**.

The CRISPR evaluation set is comprised of 571 SNP-gene links experimentally validated by CRISPR interference experiments^15,36–42^. Validated links are defined as those marked “TRUE” in the “Regulated” column of the combined CRISPR evaluation data set (see Data Availability). We derived 571 CRISPR-validated SNP-gene links from these 472 element-gene links based on SNPs that reside in a regulatory element. CRISPR results were assembled and made publicly available by ref.^35^ A full list of CRISPR studies included in this evaluation set is reported in **Supplementary Table 12**.

The GWAS-derived evaluation set is comprised of 123 SNP-gene pairs defined by non-coding variants fine-mapped to a focal trait (PIP > 0.1) with a coding variant for exactly one candidate gene ± 1 Mb from the non-coding variant attaining PIP > 0.5 for the same trait^43^. This evaluation set was defined by ref.^43^ (based on fine-mapping results from ref.^44^ on 94 diseases and complex traits from the UK Biobank^53^).

### Average enrichment metric

We used an enrichment-based metric, *average enrichment across recall values*, to assess the ability of each linking method to prioritize (assign higher linking scores to) “true” links over “false” links (as defined by an evaluation set of SNP-gene links).

For a given evaluation set and given method, we ranked candidate SNP-gene links by linking score. At each unique linking score *c* assigned to any candidate SNP-gene link, we computed enrichment and recall as follows:

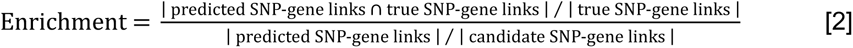

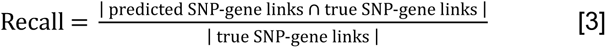

where “predicted SNP-gene links” denotes the binarized set of SNP-gene links attaining linking score ≥ *c*, “true SNP-gene links” denotes the binarized set of “true” SNP-gene links defined by the evaluation set (intersected with the set of “candidate SNP-gene links”), and “candidate SNP-gene links” denotes SNP-gene pairs profiled in at least one single-cell multiome data set (involving a SNP lying in an ATAC-seq peak and a gene expressed in the focal data set; **Table 2**). When computing enrichment for evaluation sets derived from CRISPR^15,36–42^ and GWAS^43,44^ we further restricted the set of “candidate SNP-gene links” to SNP-gene pairs tested by a CRISPR experiment^15,36–42^ (for CRISPR evaluation) or SNP-gene pairs involving a SNP with a “true” GWAS-derived link (for GWAS evaluation). We did not further restrict the set of “candidate SNP-gene links” for evaluation sets derived from eQTL^33,34^ and ABC^17,35^, as these methods can detect SNP-gene associations genome-wide. Intuitively, equation 2 quantifies the extent of overlap between predicted SNP-gene links and evaluation SNP-gene links relative to the overlap expected by chance.

For a given evaluation set and set of methods, we used equation 2 and equation 3 to construct enrichment-recall curves and observed *r* ≤ 1, the maximum recall achieved by all methods (if SCENT, Signac, ArchR, and Cicero attain maximum recall values of *r*_1_, *r*_2_, *r*_3_, and *r*_4_, respectively, we define *r* as the minimum of *r*_1_, *r*_2_, *r*_3_, and *r*_4_). For each method, we then measured the average enrichment at each unique value of recall in the range [0, *r*], thus defining *average enrichment across recall values* (or *average enrichment*, shortened) across values of recall shared by all methods. Intuitively, this metric can be interpreted as area under the enrichment-recall curve. We obtained standard errors on average enrichment for each method, as well as standard errors and p-values on the difference in average enrichment between two methods, by bootstrapping genes using 1,000 bootstrap iterations.

### AUPRC metric

As a secondary evaluation metric, we computed *area under the precision-recall curve* (AUPRC) or, equivalently, *average precision across recall values*. At each unique linking score *c* assigned to any candidate SNP-gene link, we computed precision as follows:

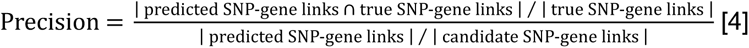

where “predicted SNP-gene links” denotes the binarized set of SNP-gene links attaining linking score ≥ *c*, “true SNP-gene links” denotes the binarized set of “true” SNP-gene links as defined by the evaluation set, and the set of “candidate SNP-gene links” is defined depending on the evaluation set (see *Average enrichment metric*).

For a given evaluation data set and set of methods, we used equation 4 and equation 3 to construct precision-recall curves for each method, observed *r* ≤ 1 (maximum recall achieved by all methods), and measured the average precision at each unique value of recall [0, *r*] for each method. We obtained standard errors on AUPRC for each method by bootstrapping genes using 1,000 iterations.

### Analyses of feature importance

To determined which features contribute most to pgBoost performance, we conducted analyses ablating sets of features from the pgBoost framework. We first trained pgBoost using features from one constituent score or one constituent score plus distance, resulting in models spanning either 2 distance-based features; 27 features derived from SCENT^25^, Signac^26^, or ArchR^27^; or 18 features derived from Cicero^28^ (29 or 20 features, respectively, using one constituent score plus distance-based features). We similarly trained gradient boosting models with features from a single constituent score ablated, resulting in models spanning 99 features (with 2 distance-based features ablated), 74 features (with SCENT^25^, Signac^26^, or ArchR^27^ features ablated), or 83 features (with Cicero^28^ features ablated). We then trained pgBoost using linking scores computed in one data set (**Table 2**), resulting in models spanning 35, 57,13 or features (linking scores computed in the 10X PBMC^47^, Luecken BMMC^48^, and SHARE-seq LCL^22^ data sets, respectively, plus distance-based features). We similarly trained gradient boosting models with features from one data set ablated, resulting in models spanning 68, 46, or 90 features (ablating linking scores computed in the 10X PBMC^47^, Luecken BMMC^48^, and SHARE-seq LCL^22^ data sets, respectively).

We also applied Shapley Additive Explanation (SHAP), a tool for interpreting complex nonlinear models^51,105^. SHAP uses positive/negative training labels and the trained model to quantify the importance of individual features driving pgBoost predictions while accounting for nonlinear interactions. We computed SHAP scores using SHAPforxgboost software^106^.

### Downsampling analyses varying cell count and sequencing depth

We performed downsampling analyses assessing the sensitivity of pgBoost and constituent method to variation in cell count, RNA count, ATAC count, and RNA + ATAC count simultaneously. We conducted all downsampling analyses in T cells from the NeurIPS data set (**Table 2** and **Supplementary Table 2**). We generated matrices reflecting 4 downsampling conditions (50% of cells; 50% of RNA counts and 100% of ATAC peak counts; 100% of RNA counts and 50% of ATAC peak counts; 50% of RNA counts and 50 % of ATAC peak counts), ran the 4 constituent methods and pgBoost in each of these downsampled conditions, and computed average enrichment across recall values (see *Average enrichment metric*) of each method in downsampled and non-downsampled conditions. (We note that Cicero only uses ATAC-seq data as input and thus is unaffected by downsampling of ATAC peak counts.)

In each downsampled condition, pgBoost is trained on the 2 distance-based features and constituent linking scores computed on downsampled data. Because ArchR software^27^ does not support modification of the ATAC peak count matrix, we did not consider ArchR in experiments downsampling ATAC or RNA counts. To reflect this limitation, we trained 4 pgBoost models ablating ArchR features: 3 models using linking scores computed in downsampled conditions (downsampled RNA, downsampled ATAC, downsampled RNA + ATAC) and 1 model using linking scores computed in non-downsampled conditions for comparison (**Supplementary Figure 16b-d**).

### Training pgBoost models in a focal cell type

To investigate whether restricting model features to a focal cell type improves power to detect SNP-gene links relevant to that cell type, we trained individual pgBoost models and generated predictions for T cells, B cells, myeloid cells, and erythroid cells in the Luecken BMMC^48^ multiome data set (**Table 2** and **Supplementary Table 2**). We restricted most analyses to the Luecken BMMC^48^ data set to avoid data set-level confounding (and selected this data set because 4 major cell types are represented by > 8,000 cells). We trained pgBoost models in a focal cell type by restricting model features to the 11 features derived from the focal cell type (linking scores and indicators, see *pgBoost framework*) plus 2 distance-based features, totaling 13 total features. We used fine-mapped eSNP-eGene pairs (maximum PIP > 0.5 vs. maximum PIP < 0.01 across GTEx tissues) as training data (intersecting these SNP-gene pairs with the set of candidate SNP-gene links profiled by the Luecken BMMC^48^ data set).

### ABC evaluation data in a focal cell type

We defined evaluation SNP-gene links relevant to each of the 4 focal cell types (T cells, B cells, myeloid cells, and erythroid cells; see *Training pgBoost models in a focal cell type*) from ABC data^17,35^. We first assigned 344 biosamples profiled by ABC data^17,35^ to either 1 focal cell type (if a biosample is closely biologically related to that cell type) or 0 focal cell types (if a biosample is not related to any of the 4 focal cell types *or* is equally related to more than 1 focal cell type), yielding 53 biosamples related to T cells, 14 biosamples related to B cells, 6 biosamples related to myeloid cells, and 1 biosample related to erythroid cells (a mapping of these 74 ABC biosamples to focal cell types is provided in **Supplementary Table 7**). We then defined 4 *cell-type-level* sets of SNP-gene links strongly implicated in any biosample related to each focal cell type (max(ABC) > 0.2 across biosamples). We finally defined *cell-type-specific* sets of SNP-gene links as the set of *cell-type-level* SNP-gene links in each focal cell type that are not as strongly implicated in the 3 non-focal cell types (defining “not as strongly implicated” via max(ABC) < 0.1 across biosamples).

### GWAS variant evaluation data

We applied the *average enrichment across recall values* metric (see *Average enrichment metric*) to assess the ability of pgBoost models trained in focal cell types to prioritize (assign higher linking scores to) causal variants fine-mapped to cell type-relevant traits in GWAS.

We analyzed GWAS fine-mapping results for 7 red blood cell or platelet-related blood cell traits (platelet count, hemoglobin A1c, hemoglobin, red blood cell count, mean corpuscular volume, mean corpuscular hemoglobin, mean corpuscular hemoglobin concentration) and 7 autoimmune diseases and granulocyte-related blood cell traits (white blood cell count, monocyte count, eosinophil count, autoimmune disease, lymphocyte count, neutrophil count, basophil count) from the UK Biobank^44,53^. 518 variants (resp. 493 variants) scored by pgBoost were fine-mapped to any red blood cell or platelet-related blood cell traits (resp. autoimmune disease or granulocyte-related blood cell trait) with PIP > 0.2. (We selected a PIP threshold of > 0.2 instead of > 0.5 to specify a larger variant evaluation set, as approximately 200 variants were fine-mapped to each category of GWAS traits with PIP > 0.5.)

For a given evaluation set of fine-mapped GWAS variants and a given pgBoost model corresponding to a focal cell type, we ranked candidate SNPs by the maximum linking score across all candidate SNP-gene links involving the focal SNP. At each unique score *c* assigned to any SNP, we computed variant-level enrichment and recall as follows:

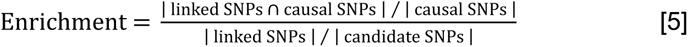

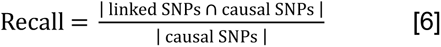

where “linked SNPs” denotes the binarized set of SNPs attaining maximum linking score ≥ *c*, “causal SNPs” denotes the binarized set of SNPs attaining PIP > 0.2 for any trait in the focal category, and “candidate SNPs” denotes SNPs lying in ATAC-seq peaks in the Luecken BMMC^48^ data set (**Table 2** and **Supplementary Table 2**).

For a given evaluation set and set of methods, we used equation 5 and equation 6 to construct variant-level enrichment-recall curves for each method, observed *r* ≤ 1 (maximum recall achieved by all methods), and measured the average enrichment at each unique value of recall [0, *r*] for each method. We obtained standard errors on variant-level average enrichment by bootstrapping genes using 1,000 iterations.

