## Supplementary Figures 1-34 for "Linking regulatory variants to target genes by integrating single-cell multiome methods and genomic distance"

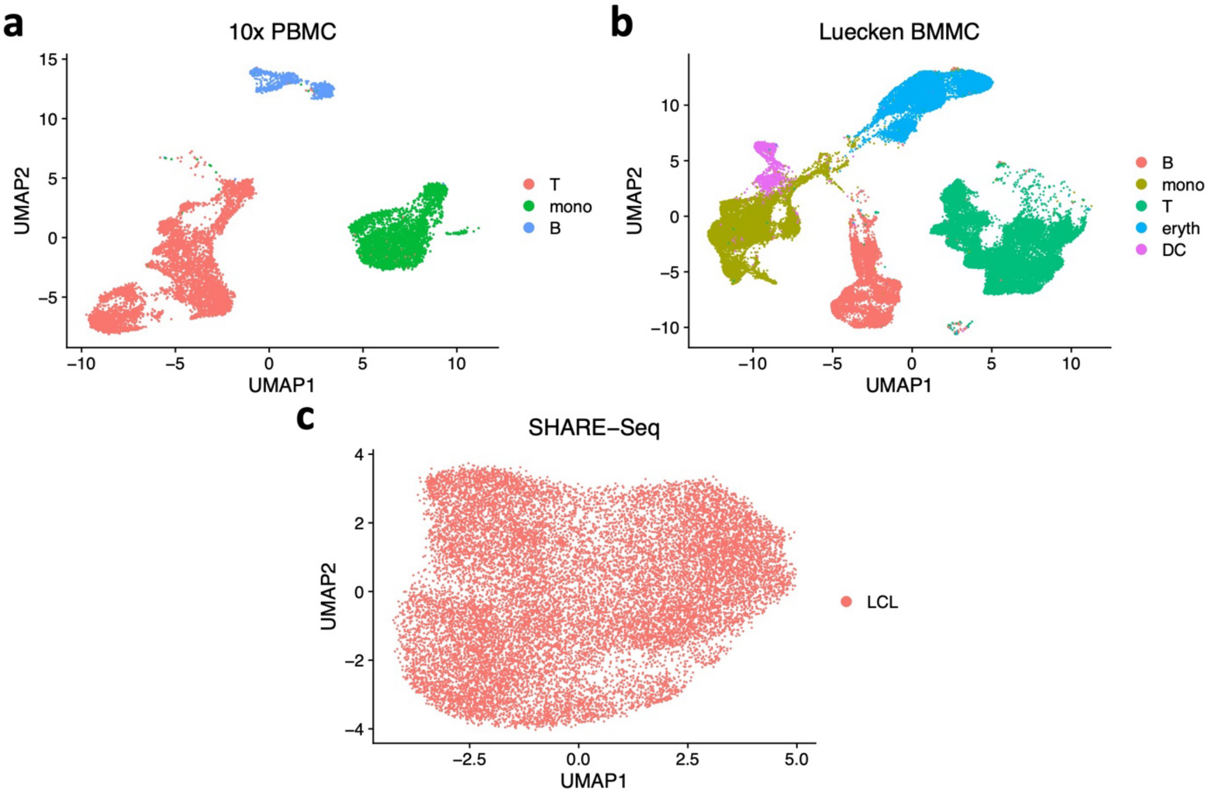


**SF1: UMAP plots of single-cell data sets.** Cells in 3 single-cell RNA/ATAC-seq multiome datasets summarized in a joint UMAP visualization representing both gene expression and chromatin accessibility (built on a joint weighted nearest neighbor graph[102]) and colored by cell type.

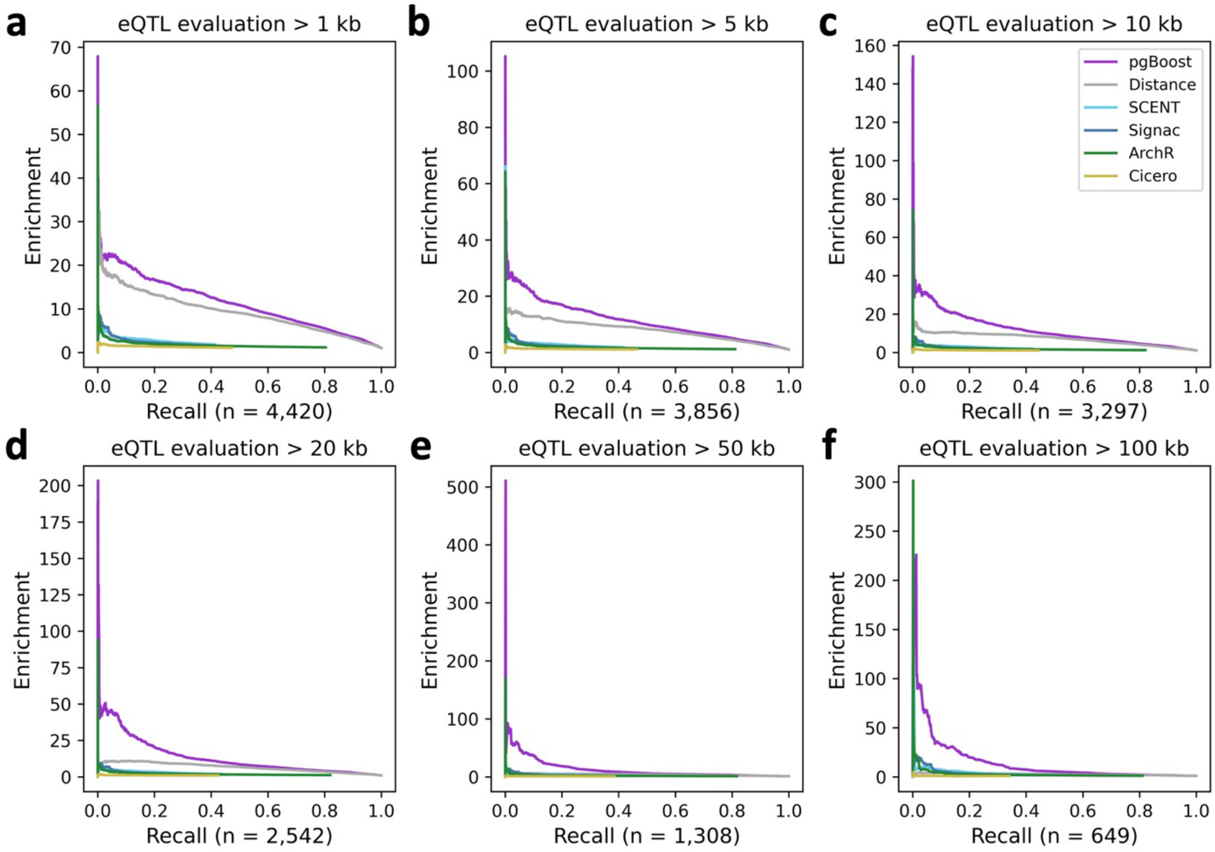


**SF2: Enrichment-recall curves of pgBoost and other methods on eQTL evaluation data set.** Enrichment-recall curves of links predicted by pgBoost, distance, and 4 constituent methods for 4,420 fine-mapped eSNP-eGene pairs attaining maximum PIP > 0.5 across GTEx tissues, at various distance thresholds. The number of positive evaluation links at each distance threshold is specified in parentheses.

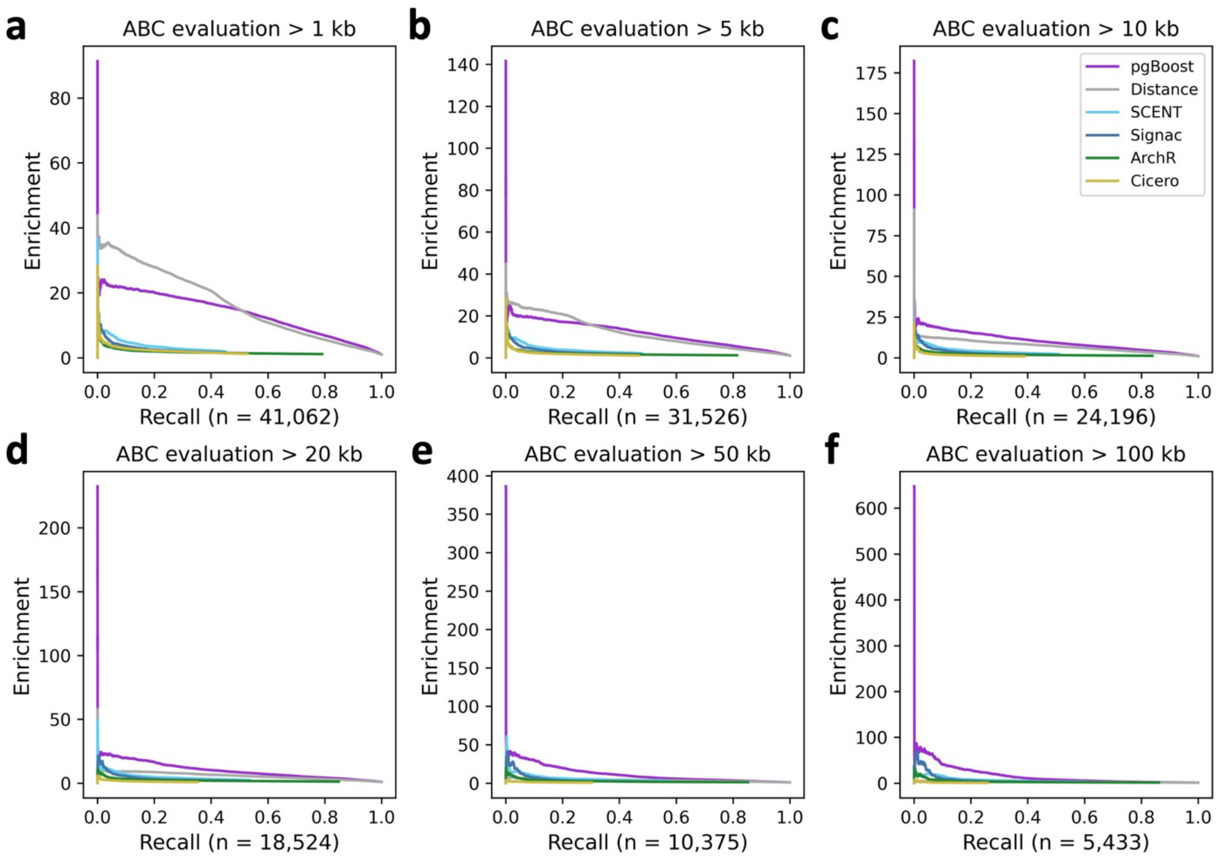


**SF3: Enrichment-recall curves of pgBoost and other methods on ABC evaluation data set.** Enrichment-recall curves of links predicted by pgBoost, distance, and 4 constituent methods for SNP-gene pairs attaining maximum ABC score > 0.2 across 344 biosamples, at various distance thresholds. The number of positive evaluation links at each distance threshold is specified in parentheses.

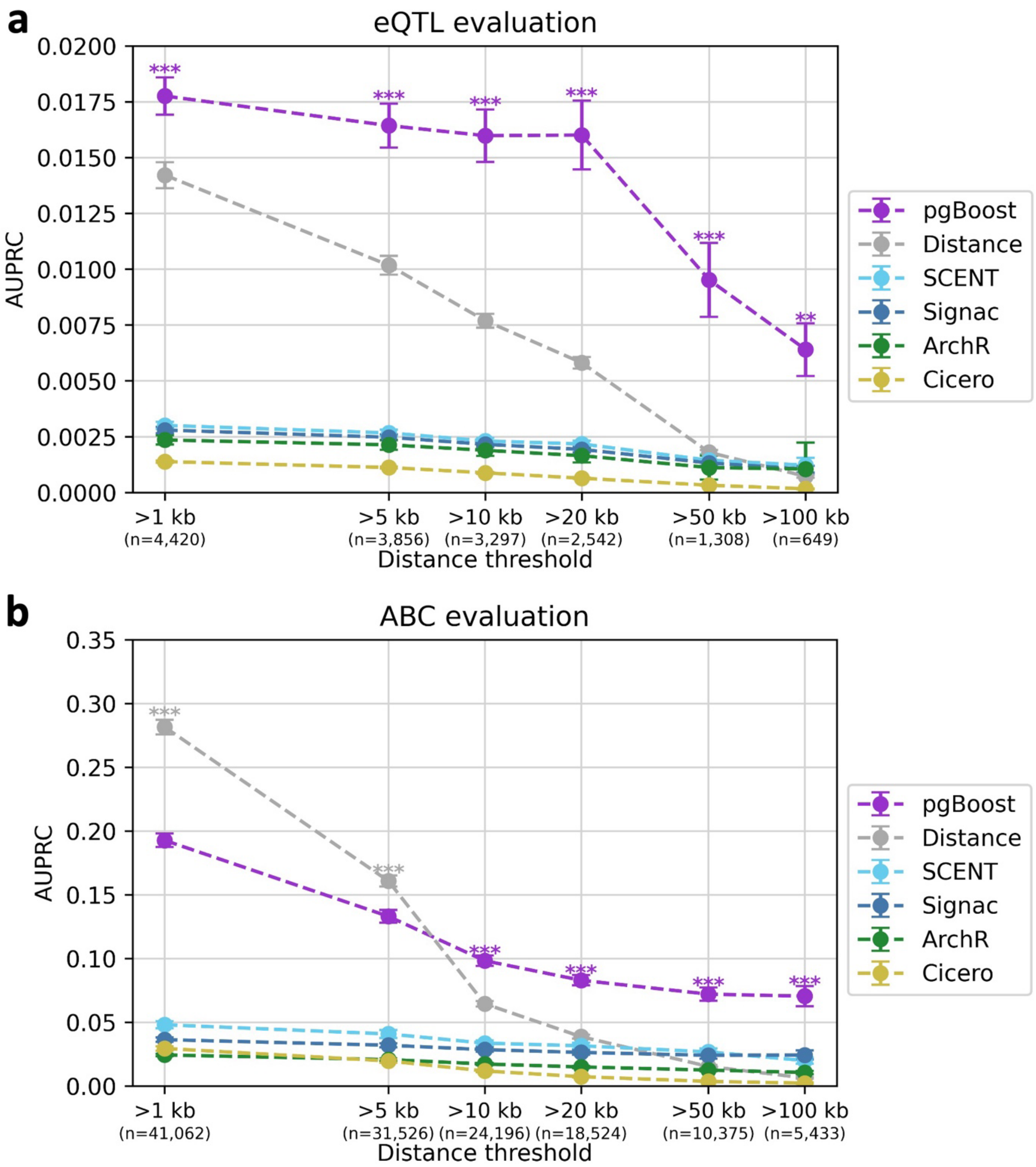


**SF4: AUPRC of pgBoost and other methods on eQTL and ABC evaluation data sets. a)** Area under the precision-recall curve (AUPRC) across recall values (in [0,0.35]) of links predicted by pgBoost, distance, and 4 constituent methods for 4,420 fine-mapped eSNP-eGene pairs attaining maximum PIP > 0.5 across GTEx tissues, at various distance thresholds. **b)** Area under the precision-recall curve (AUPRC) across recall values (in [0,0.25]) of links predicted by pgBoost, distance, and 4 constituent methods for 41,062 SNP-gene pairs attaining maximum ABC score > 0.2 across 344 biosamples, at various distance thresholds. The number of positive evaluation links at each distance threshold is specified in parentheses. Confidence intervals denote standard errors. Stars denote p-values for difference (*: $p < 0.05$, **: $p < 0.01$, ***: $p < 0.001$) of top method vs. each other method.

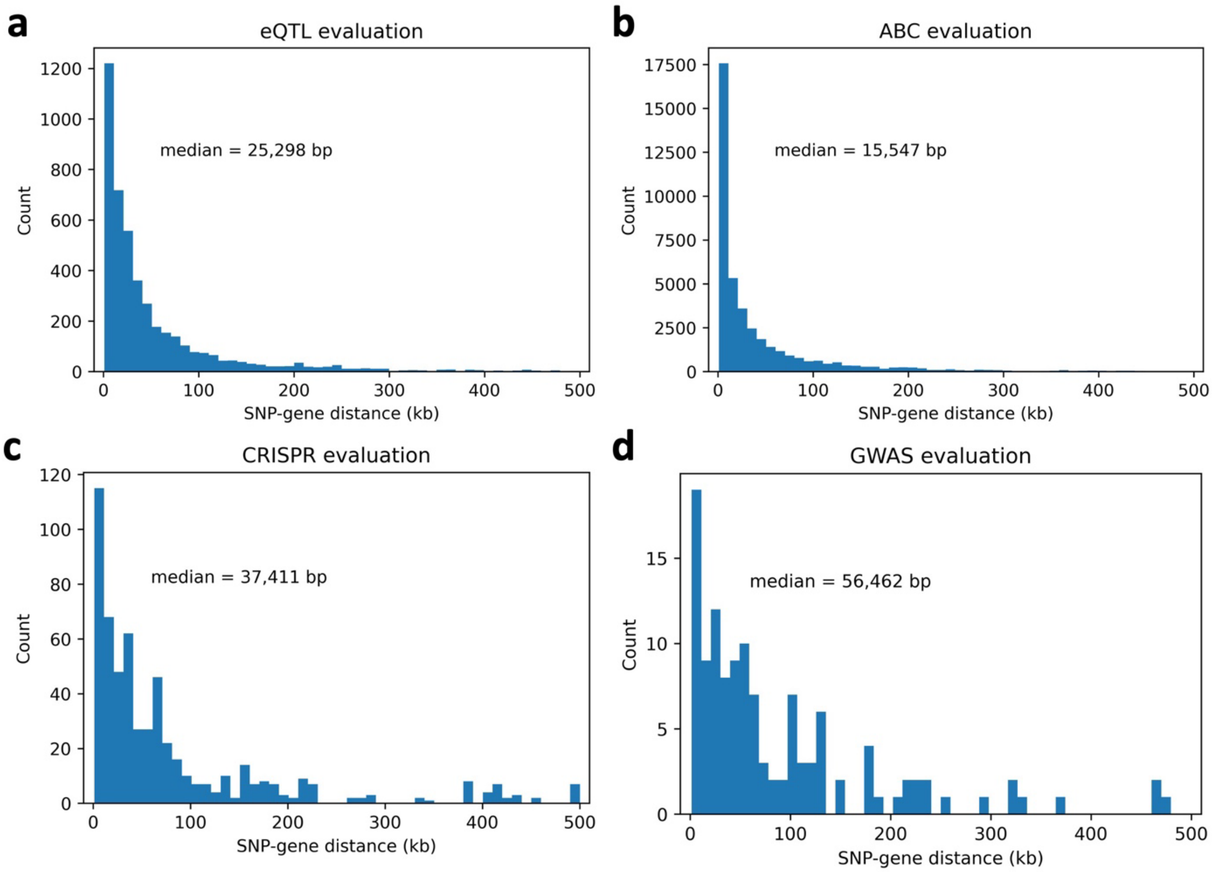


**SF5: Distance distributions of eQTL, ABC, CRISPR, and GWAS evaluation data sets.** SNP-gene distance distributions of evaluation sets comprised of **(a)** 4,420 fine-mapped eSNP-eGene pairs attaining maximum PIP > 0.5 across GTEx tissues, **(b)** 41,062 SNP-gene pairs attaining maximum ABC score > 0.2 across 344 biosamples, **(c)** 571 links validated by CRISPR, and **(d)** 123 non-coding SNP-gene pairs derived from fine-mapped GWAS variants with a unique fine-mapped coding variant within a 2 Mb window. The median SNP-gene distance is reported within each plot.

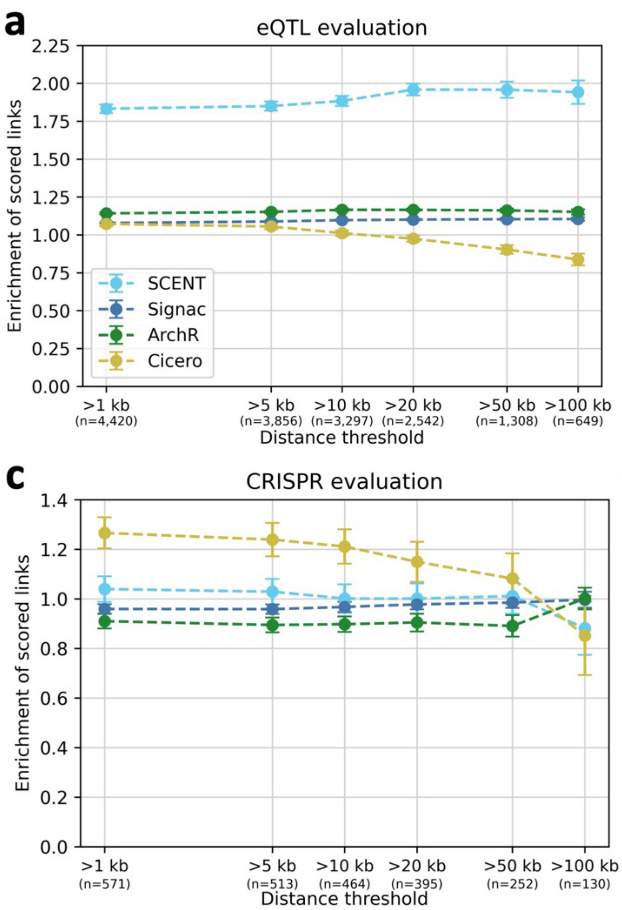


**SF6: Enrichment of candidate links scored for eQTL, ABC, CRISPR, and GWAS evaluation data sets.** Enrichment of the subset of candidate SNP-gene links scored by 4 constituent methods for **(a)** 4,420 fine-mapped eSNP-eGene pairs attaining maximum PIP > 0.5 across GTEx tissues , **(b)** 41,062 SNP-gene pairs attaining maximum ABC score > 0.2 across 344 biosamples, **(c)** 571 links validated by CRISPR, and **(d)** 123 non-coding SNP-gene pairs derived from fine-mapped GWAS variants with a unique fine-mapped coding variant within a 2 Mb window, at various distance thresholds. The number of positive evaluation links at each distance threshold is specified in parentheses. Confidence intervals denote standard errors. SCENT employs stringent criteria for selecting candidate peak-gene links to test (**Table 1** and **Supplementary Table 1**), thereby pre-selecting a smaller set of scored links (**Supplementary Table 11**) with higher enrichments for eQTL and ABC evaluations. This effect is reduced for the CRISPR and GWAS evaluations because enrichments are computed against a restricted background of candidate links tested by CRISPR (**c**) and candidate links including a variant with a gold-standard GWAS link (**d**), respectively (**Methods**).

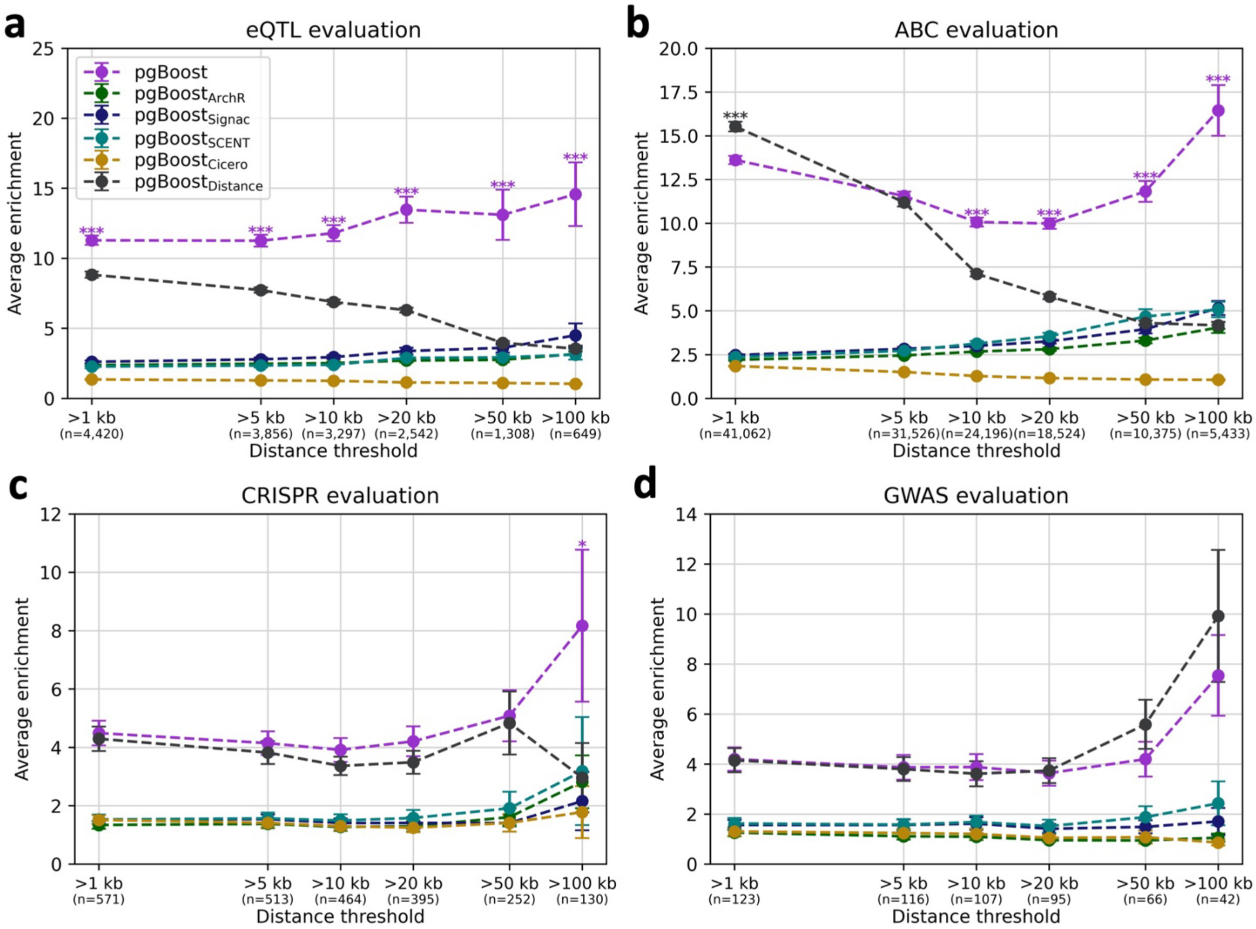


**SF7: Performance of pgBoost models restricted to a single constituent method or distance.** Average enrichment across recall values (in [0,1]) of links predicted by pgBoost restricted to linking scores from single constituent methods or genomic distance for **(a)** 4,420 fine-mapped eSNP-eGene pairs attaining maximum PIP > 0.5 across GTEx tissues, **(b)** 41,062 SNP-gene pairs attaining maximum ABC score > 0.2 across 344 biosamples, **(c)** 571 links validated by CRISPR, and **(d)** 123 non-coding SNP-gene pairs derived from fine-mapped GWAS variants with a unique fine-mapped coding variant within a 2 Mb window, at various distance thresholds. The number of positive evaluation links at each distance threshold is specified in parentheses. Confidence intervals denote standard errors. Stars denote p-values for difference (*: p < 0.05, **: p < 0.01, ***: p < 0.001) of top method vs. each other method.

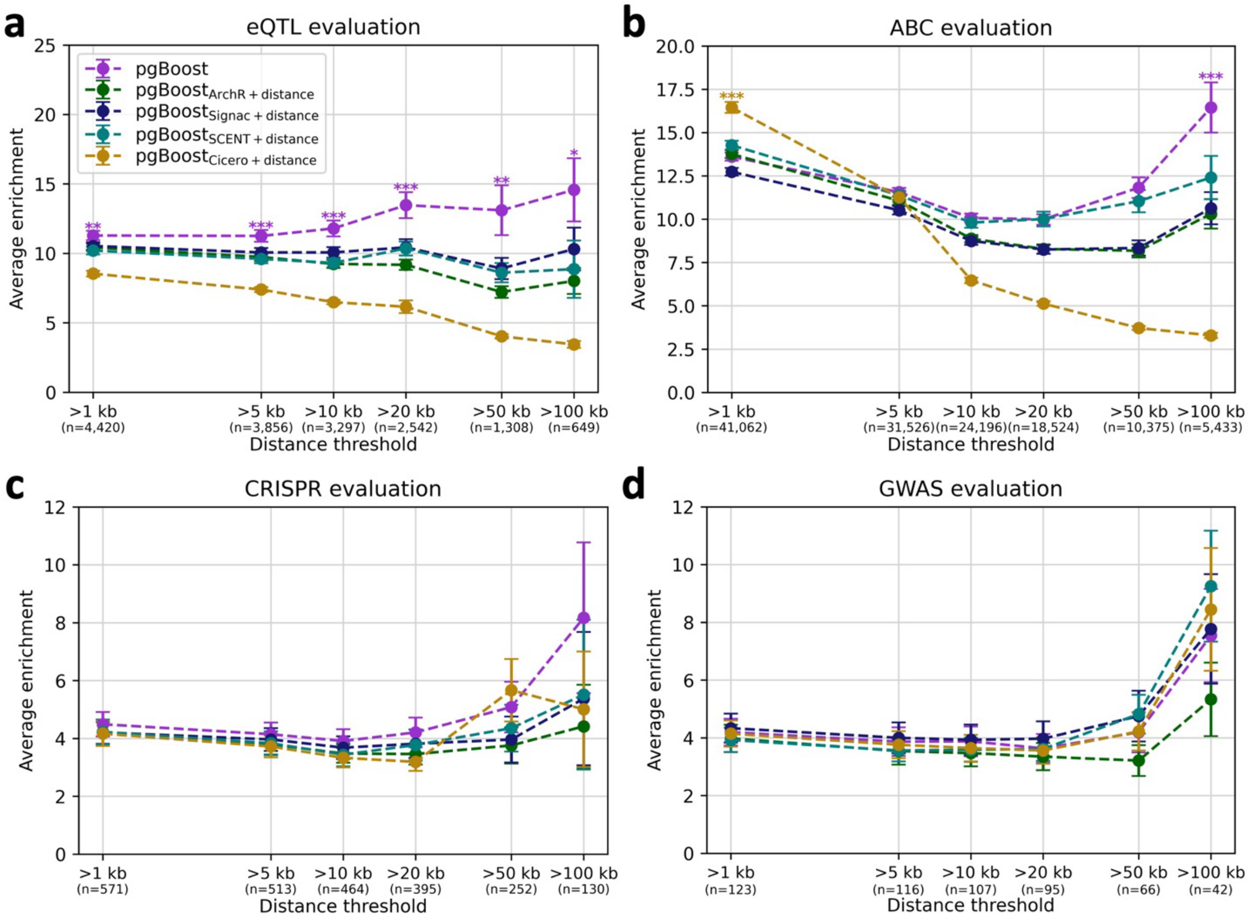


**SF8: Performance of pgBoost models restricted to a single constituent method plus distance.** Average enrichment across recall values (in [0,1]) of links predicted by pgBoost restricted to linking scores from single constituent methods plus distance-based features for **(a)** 4,420 fine-mapped eSNP-eGene pairs attaining maximum PIP > 0.5 across GTEx tissues, **(b)** 41,062 SNP-gene pairs attaining maximum ABC score > 0.2 across 344 biosamples, **(c)** 571 links validated by CRISPR, **(d)** and 123 non-coding SNP-gene pairs derived from fine-mapped GWAS variants with a unique fine-mapped coding variant within a 2 Mb window, at various distance thresholds. The number of positive evaluation links at each distance threshold is specified in parentheses. Confidence intervals denote standard errors. Stars denote p-values for difference (*: $p < 0.05$, **: $p < 0.01$, ***: $p < 0.001$) of top method vs. each other method.

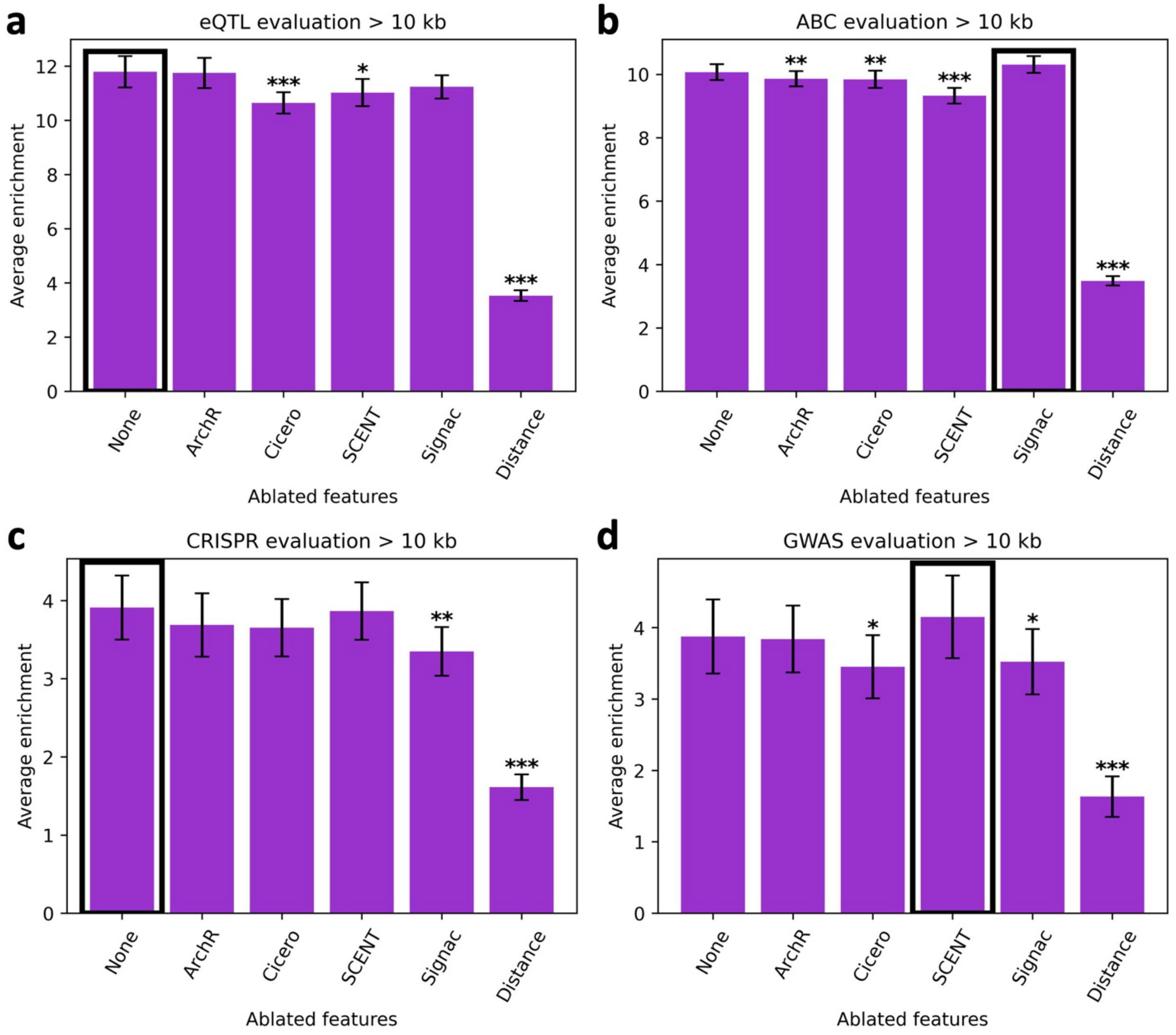


**SF9: Performance of pgBoost models ablating a single constituent method or distance.** Average enrichment across recall values (in [0,1]) of links predicted by pgBoost with one constituent method or distance features ablated for **(a)** 4,420 fine-mapped eSNP-eGene pairs attaining maximum PIP > 0.5 across GTEx tissues, **(b)** 41,062 SNP-gene pairs attaining maximum ABC score > 0.2 across 344 biosamples, **(c)** 571 links validated by CRISPR, and **(d)** 123 non-coding SNP-gene pairs derived from fine-mapped GWAS variants with a unique fine-mapped coding variant within a 2 Mb window. Because enrichment differences are small and highly similar across distance thresholds, results at >10kb are shown as barplots to aid in visual comparison. Confidence intervals denote standard errors. Stars denote p-values for difference (*: $p < 0.05$, **: $p < 0.01$, ***: $p < 0.001$) of focal method vs. top method (denoted in black outline).

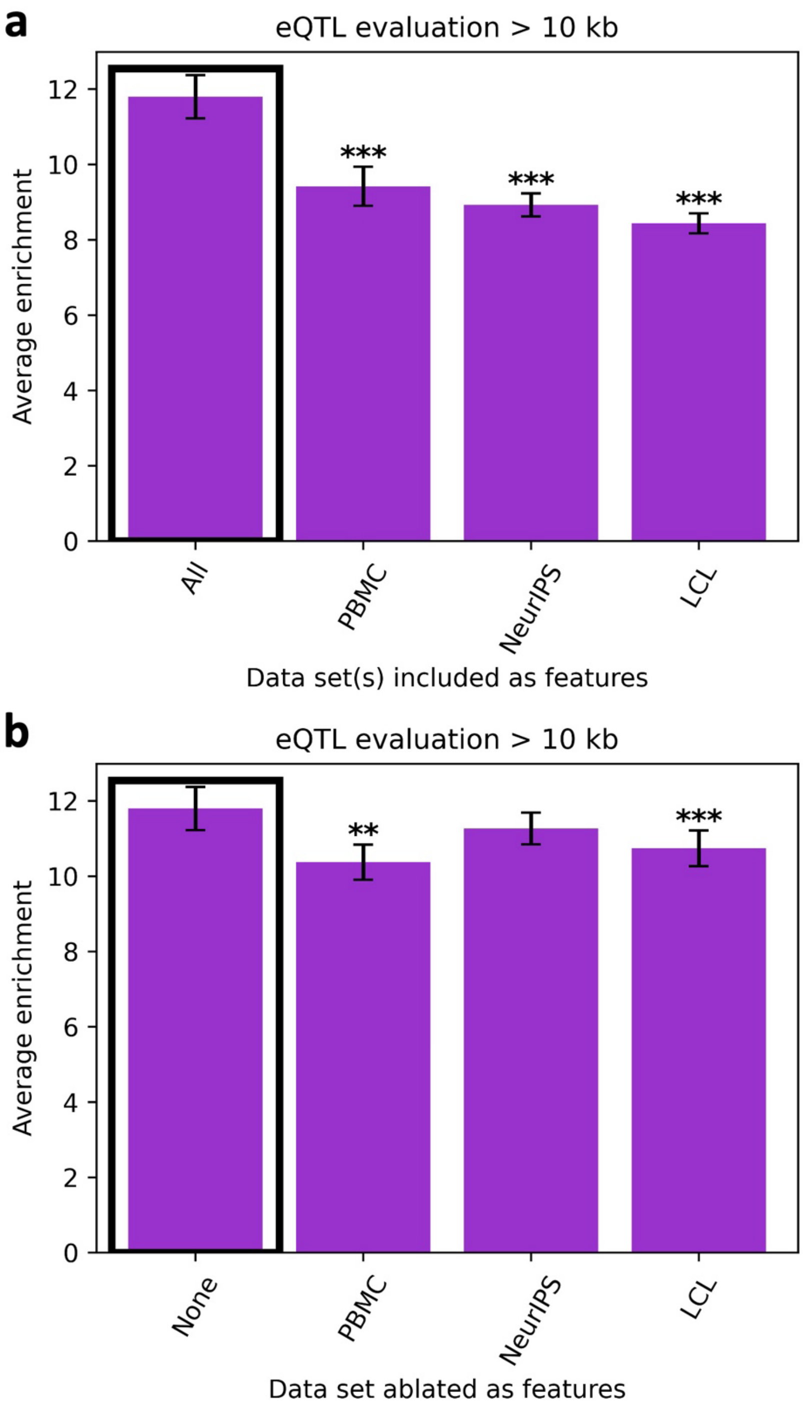


**SF10: Performance of pgBoost models ablating data sets.** Average enrichment across recall values (in [0,1]) of links predicted by **(a)** pgBoost restricted to linking scores from a single data set plus distance features and **(b)** pgBoost with features from a single data set ablated, for 4,420 fine-mapped eSNP-eGene pairs attaining maximum PIP > 0.5 across GTEx tissues. Because enrichment differences are highly similar across distance thresholds, results at >10kb are shown as barplots to aid in visual comparison. Confidence intervals denote standard errors. Stars denote p-values for difference (*: $p < 0.05$, **: $p < 0.01$, ***: $p < 0.001$) of focal method vs. top method (denoted in black outline).

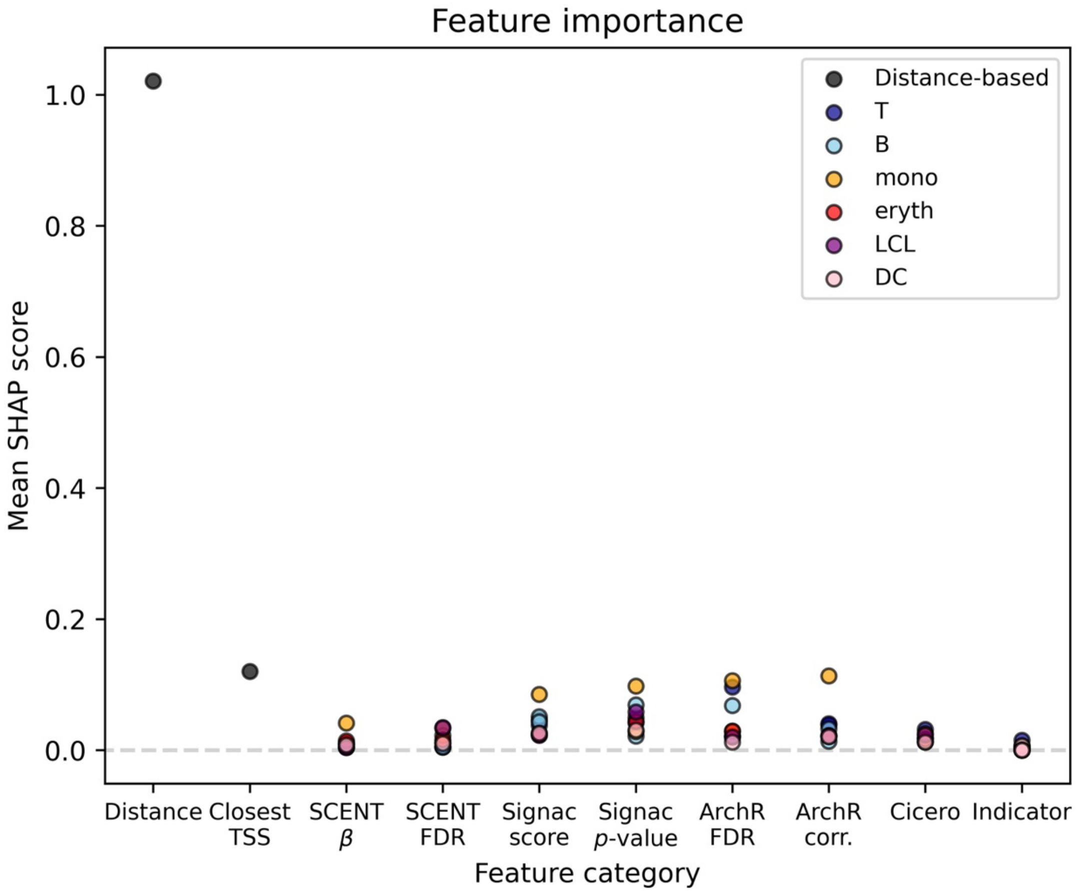


**SF11**: **SHAP analysis of pgBoost feature importance.** Mean SHAP scores of pgBoost features, categorized by feature type (denoted by x-axis position) and color-coded by cell type (if applicable). Distance attains the highest mean SHAP score followed by the binary indicator for "closest TSS," indicating that these distance-based features contribute most to linking predictions. Signac and ArchR-derived features attain slightly higher mean SHAP scores than SCENT and Cicero (possibly because Signac and ArchR score more candidate links than SCENT and Cicero).

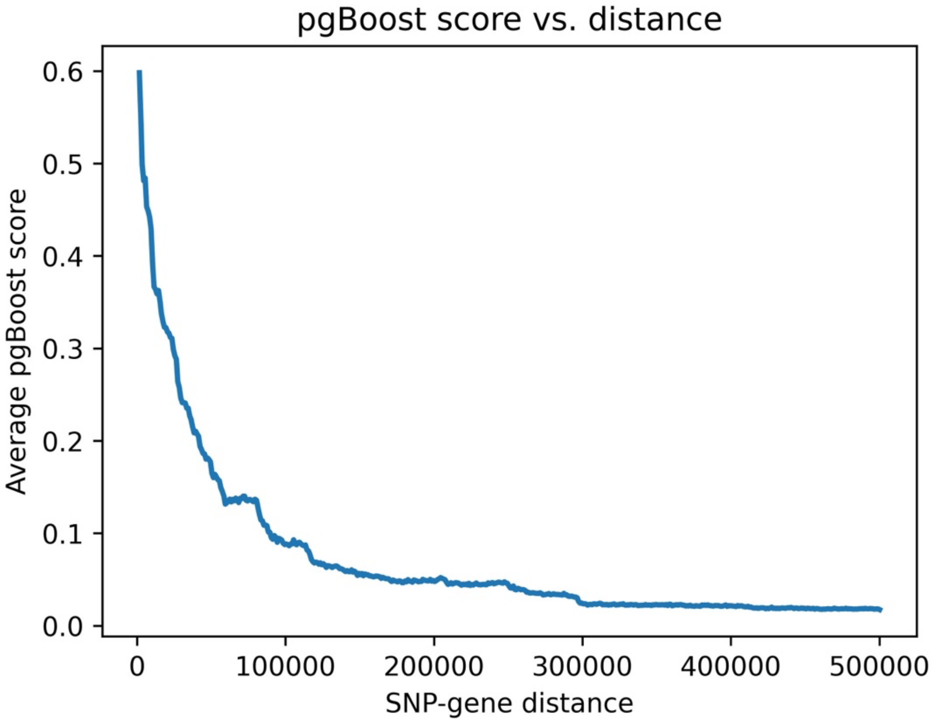


**SF12: Average pgBoost scores at different linking distances.** Relationship between average SNP-gene distance and average pgBoost score across. Candidate links are partitioned equally into 500 bins by SNP-gene distance (each represented by 1 point). In general, more proximal SNP-gene links are assigned higher pgBoost scores.

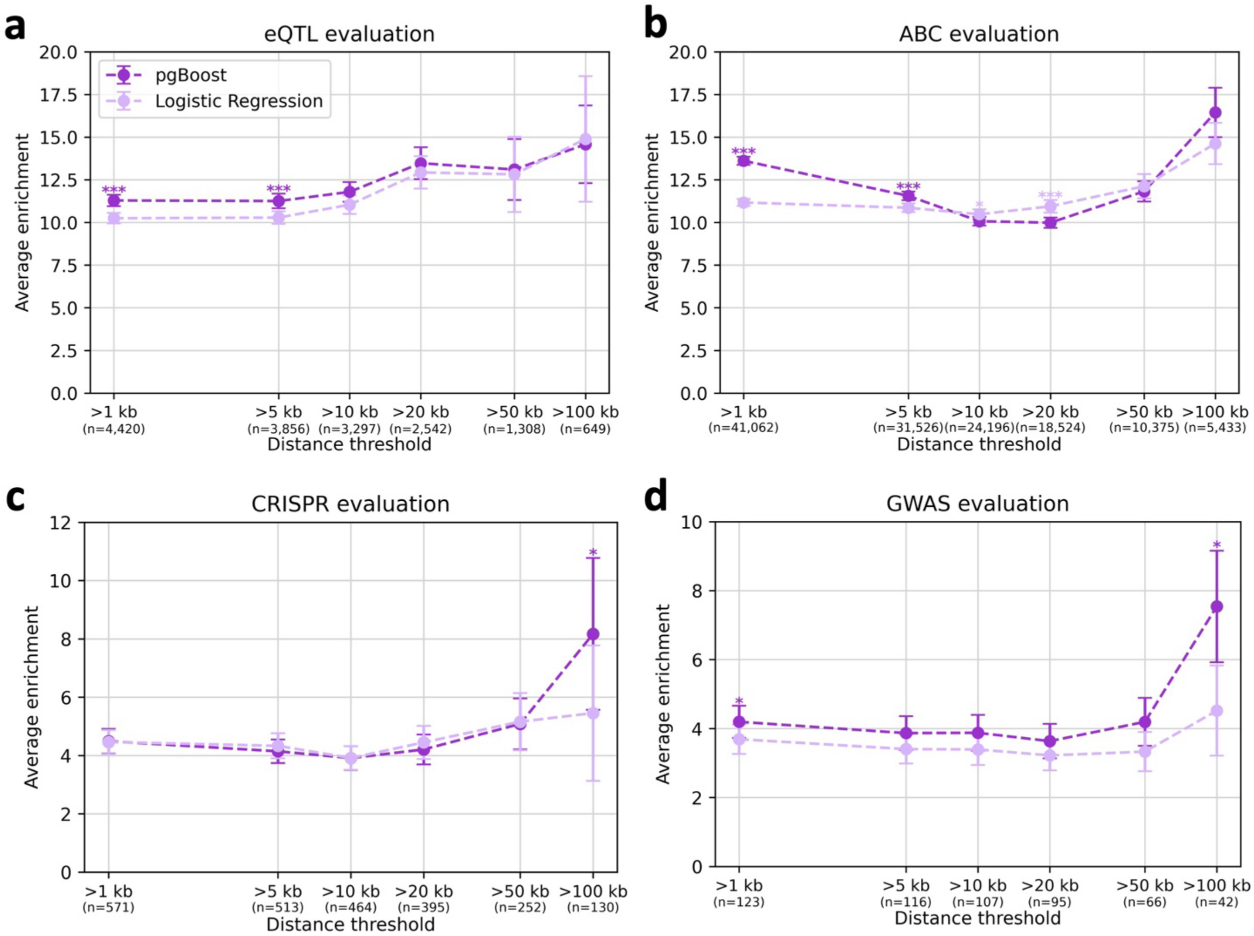


**SF13: Performance of pgBoost vs. a logistic regression model.** Average enrichment across recall values (in [0,1]) of links predicted by pgBoost vs. a logistic regression model with identical features on **(a)** 4,420 fine-mapped eSNP-eGene pairs attaining maximum PIP > 0.5 across GTEx tissues, **(b)** 41,062 SNP-gene pairs attaining maximum ABC score > 0.2 across 344 biosamples, **(c)** 571 links validated by CRISPR, and **(d)** 123 non-coding SNP-gene pairs derived from fine-mapped GWAS variants with a unique fine-mapped coding variant within a 2 Mb window, at various distance thresholds. The number of positive evaluation links at each distance threshold is specified in parentheses. Confidence intervals denote standard errors. Stars denote p-values for difference (*: p < 0.05, **: p < 0.01, ***: p < 0.001) of top method vs. each other method.

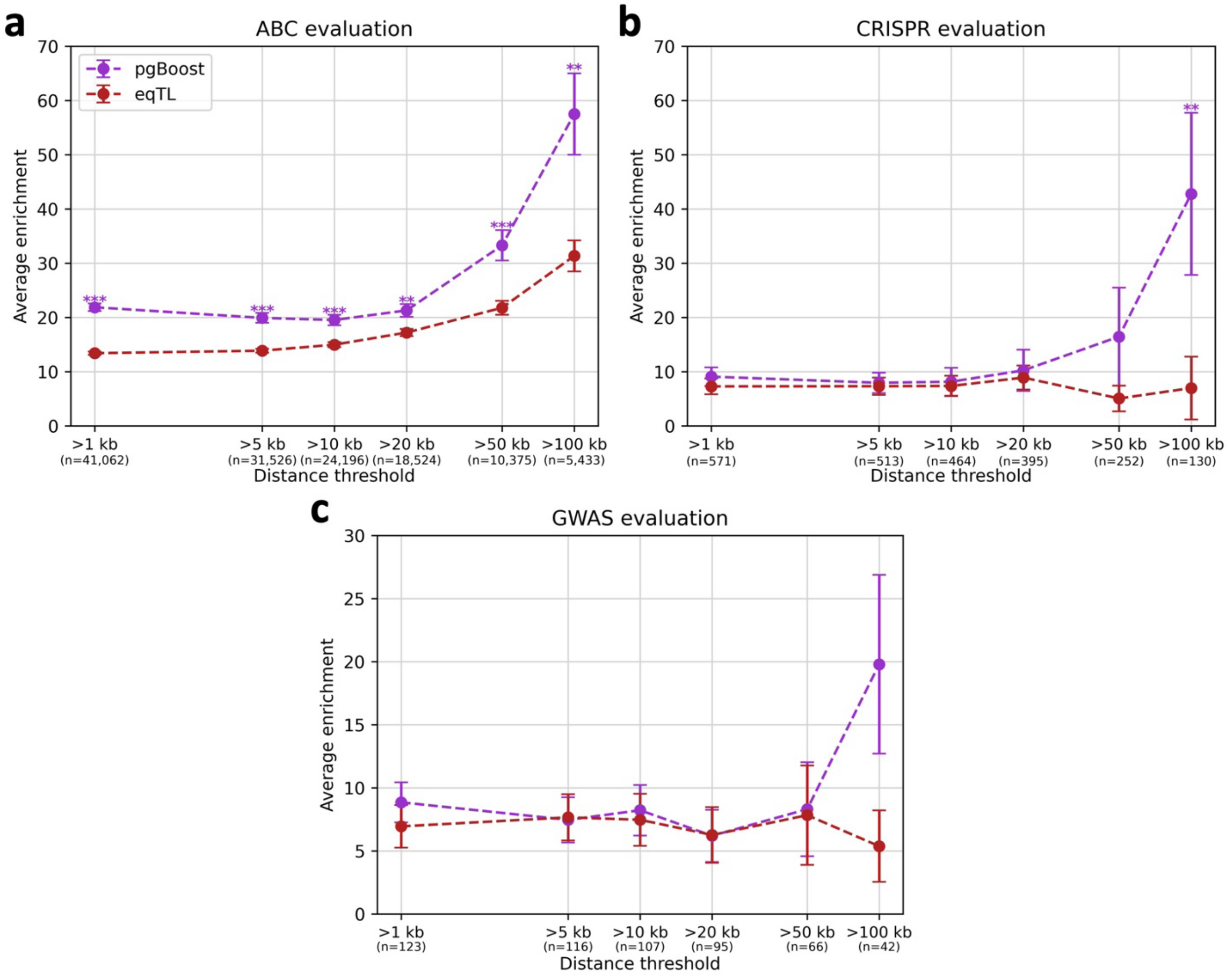


**SF14: Performance of pgBoost vs. fine-mapped eQTL. a)** Average enrichment across recall values (in [0,0.15]) of links predicted by pgBoost vs. fine-mapped eQTL for 41,062 SNP-gene pairs attaining maximum ABC score > 0.2 across 344 biosamples, at various distance thresholds. **b)** Average enrichment across recall values (in [0,0.05]) of links predicted by pgBoost vs. fine-mapped eQTL for 571 links validated by CRISPR, at various distance thresholds. **c)** Average enrichment across recall values (in [0,0.1]) of links predicted by pgBoost vs. fine-mapped eQTL for 123 non-coding SNP-gene pairs derived from fine-mapped GWAS variants with a unique fine-mapped coding variant within a 2 Mb window, at various distance thresholds. The number of positive evaluation links at each distance threshold is specified in parentheses. Confidence intervals denote standard errors. Stars denote p-values for difference (*: $p < 0.05$, **: $p < 0.01$, ***: $p < 0.001$) of top method vs. each other method.

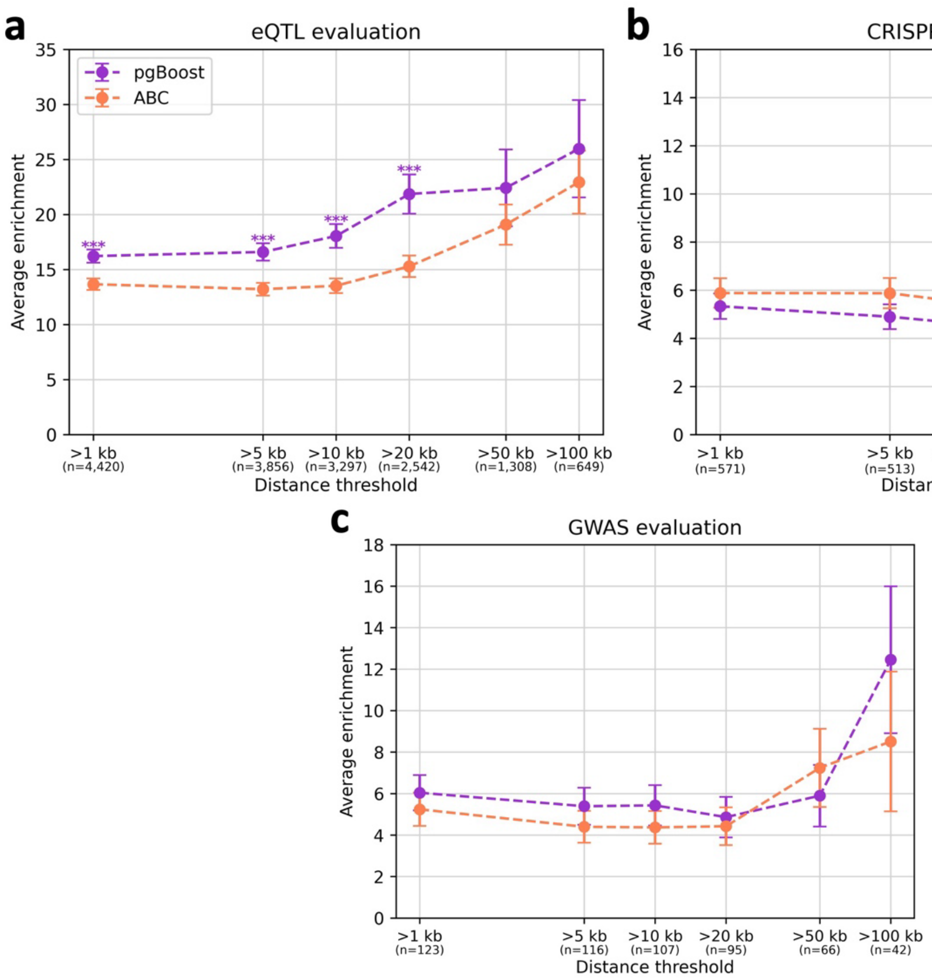


**SF15: Performance of pgBoost vs. ABC. a)** Average enrichment across recall values (in [0,0.51]) of links predicted by pgBoost vs. ABC for 4,420 fine-mapped eSNP-eGene pairs attaining maximum PIP > 0.5 across GTEx tissues, at various distance thresholds. **b)** Average enrichment across recall values (in [0,0.71]) of links predicted by pgBoost vs. ABC for 571 links validated by CRISPR, at various distance thresholds. **c)** Average enrichment across recall values (in [0,0.71]) of links predicted by pgBoost vs. ABC for 123 non-coding SNP-gene pairs derived from fine-mapped GWAS variants with a unique fine-mapped coding variant within a 2 Mb window, at various distance thresholds. The number of positive evaluation links at each distance threshold is specified in parentheses. Confidence intervals denote standard errors. Stars denote p-values for difference (*: $p < 0.05$, **: $p < 0.01$, ***: $p < 0.001$) of top method vs. each other method.

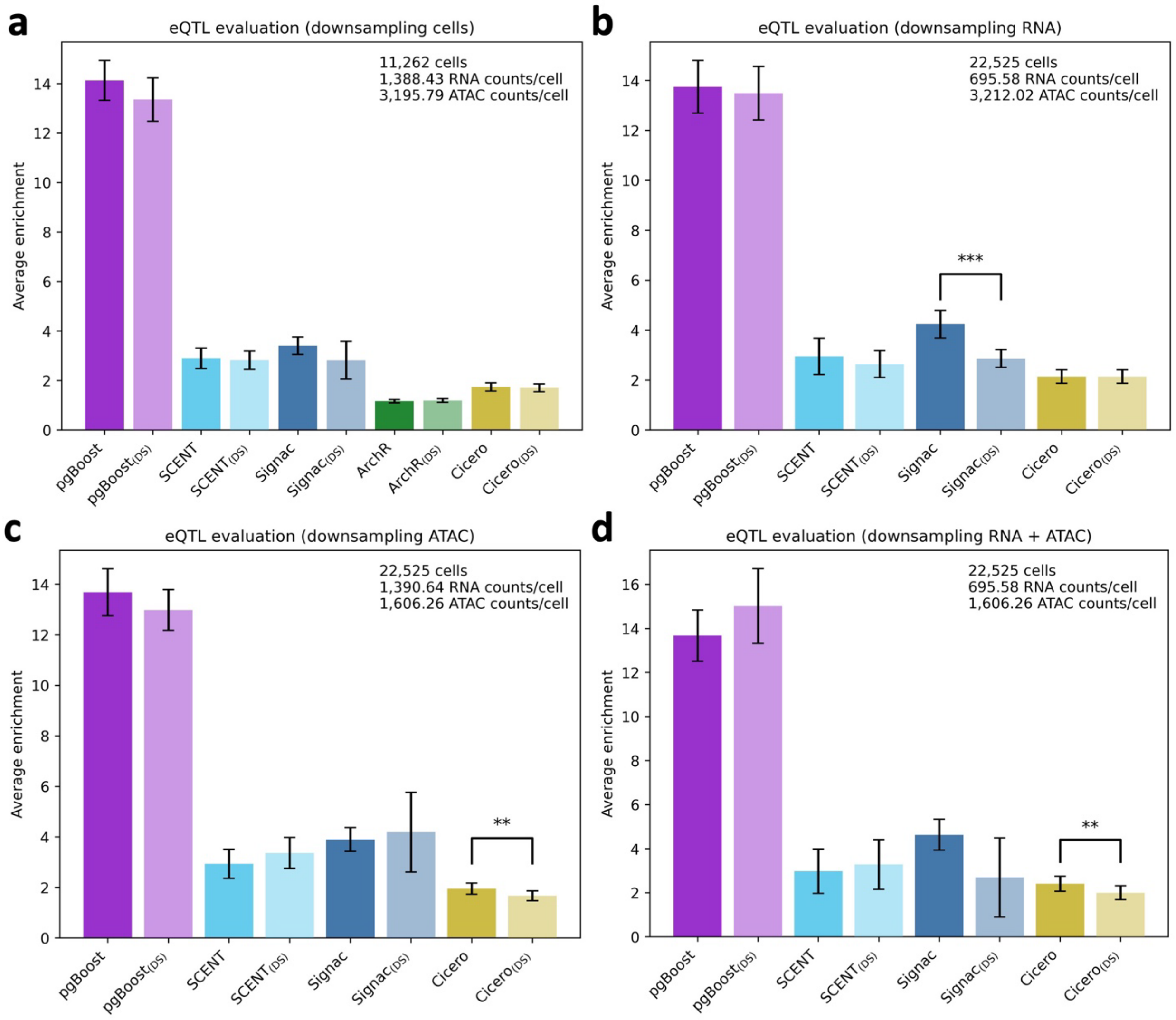


**SF16: Performance of pgBoost and constituent methods in downsampled conditions.** Average enrichment across recall values of links predicted by pgBoost and 4 constituent methods using all data from NeurIPS T cells (**Table 2** and **Supplementary Table 2**) vs. downsampling to **(a)** 50% of cells, **(b)** 50% of RNA counts, **(c)** 50% of ATAC counts, **(d)** 50% of RNA counts and 50% of ATAC counts, for 4,420 fine-mapped eSNP-eGene pairs attaining maximum PIP > 0.5 across GTEx tissues. Average enrichments are measured across recall values in [0, 0.13] **(a)**, [0, 0.07] **(b)**, [0, 0.09] **(c)**, and [0, 0.05] **(d)**. Dark and light bars report average enrichments for each method in non-downsampled and downsampled ("DS") conditions, respectively. Stars denote p-values for difference (*: $p < 0.05$, **: $p < 0.01$, ***: $p < 0.001$) of focal method in downsampled vs. non-downsampled conditions. Downsampling conditions (number of cells, average per-cell RNA and ATAC counts) are reported within each plot. Because ArchR software does not support modification of the ATAC peak count matrix, ArchR is excluded from analyses **b-d**; in addition, both pgBoost and $pgBoost_{(DS)}$ are trained excluding ArchR features in analyses **b-d.** We note that Cicero performance is unaffected by RNA count downsampling **(b)** because Cicero only uses ATAC-seq data as input.

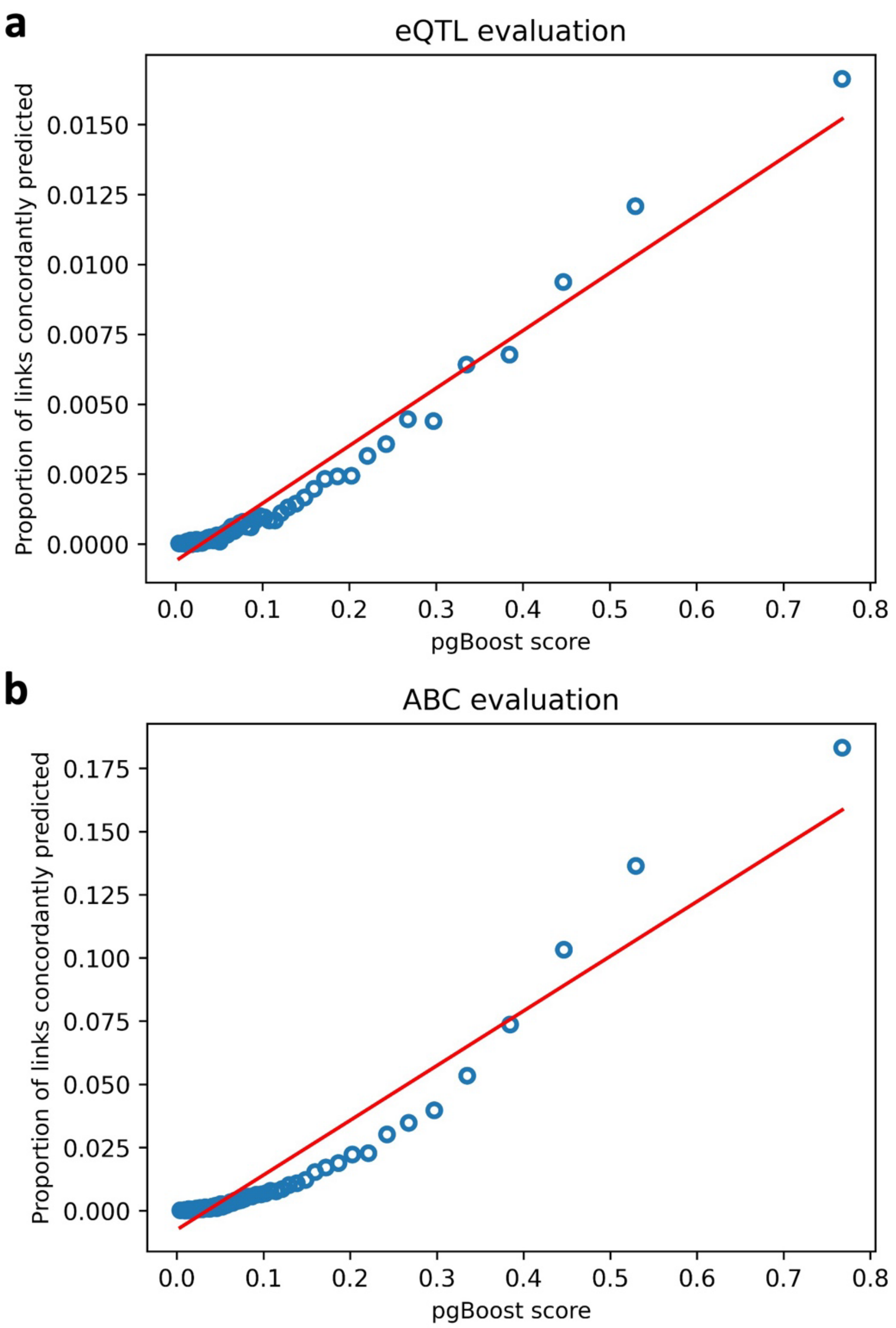


**SF17: Calibration of the pgBoost probabilistic score.** Relationship between average pgBoost score and the proportion of links concordantly predicted by **(a)** 4,420 fine-mapped eSNP-eGene pairs attaining maximum PIP > 0.5 across GTEx tissues and **(b)** 41,062 SNP-gene pairs attaining maximum ABC score > 0.2 across 344 biosamples. All 4,503,599 candidate links are partitioned equally into 100 bins by pgBoost score (each represented by 1 point). The discrepancy between predicted probability and actual proportion of concordantly predicted links is expected given the different definitions of "negative" SNP-gene links in model training (link includes a gene in the set of positive links) vs. enrichment computations (any candidate SNP-gene link that is not a positive link) (**Methods**). The relationship between proportion of links concordantly predicted and pgBoost score is not perfectly linear, supporting the use of binarized sets of top-scoring links predicted by pgBoost rather than the probabilistic score in downstream analyses.

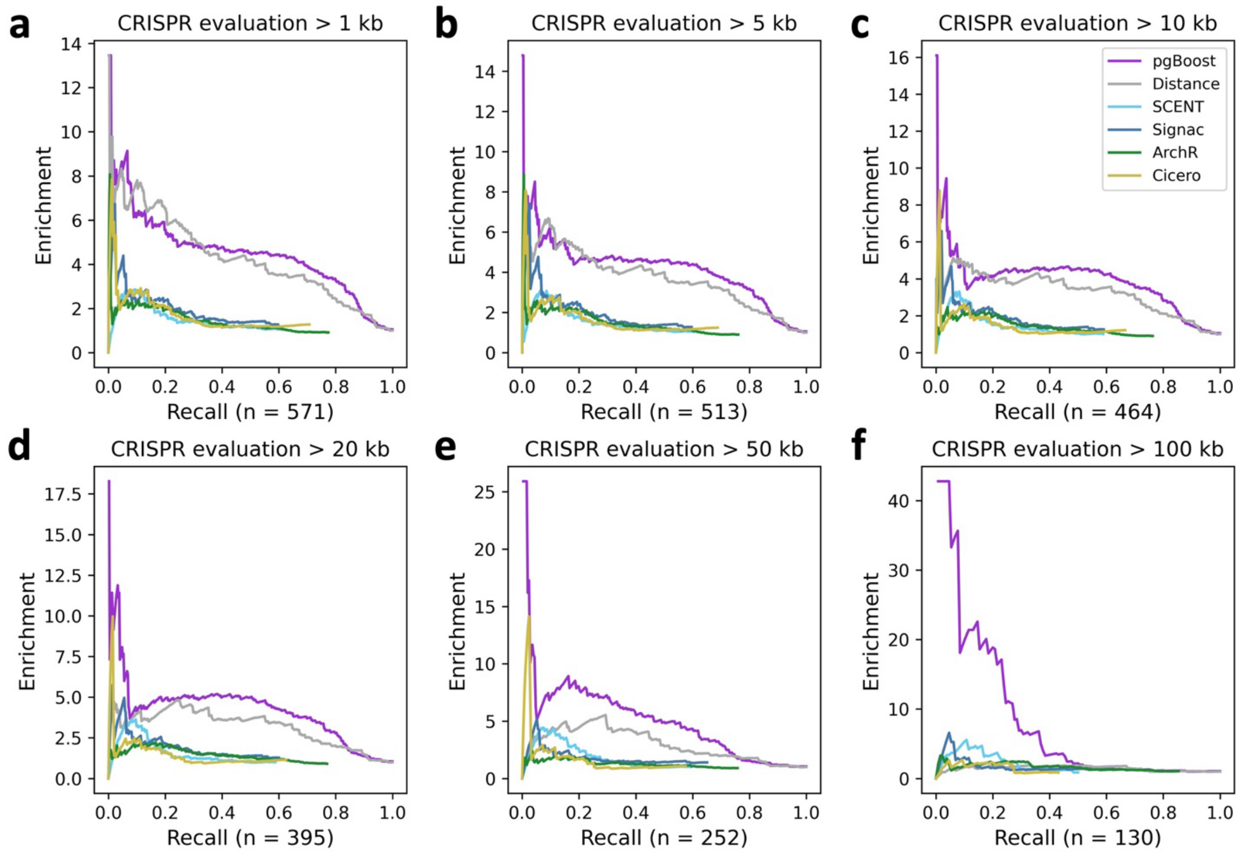


**SF18: Enrichment-recall curves of pgBoost and other methods on CRISPR evaluation data set.** Enrichment-recall curves of links predicted by pgBoost, distance, and 4 constituent methods for 571 links validated by CRISPR, at various distance thresholds. The number of positive evaluation links at each distance threshold is specified in parentheses.

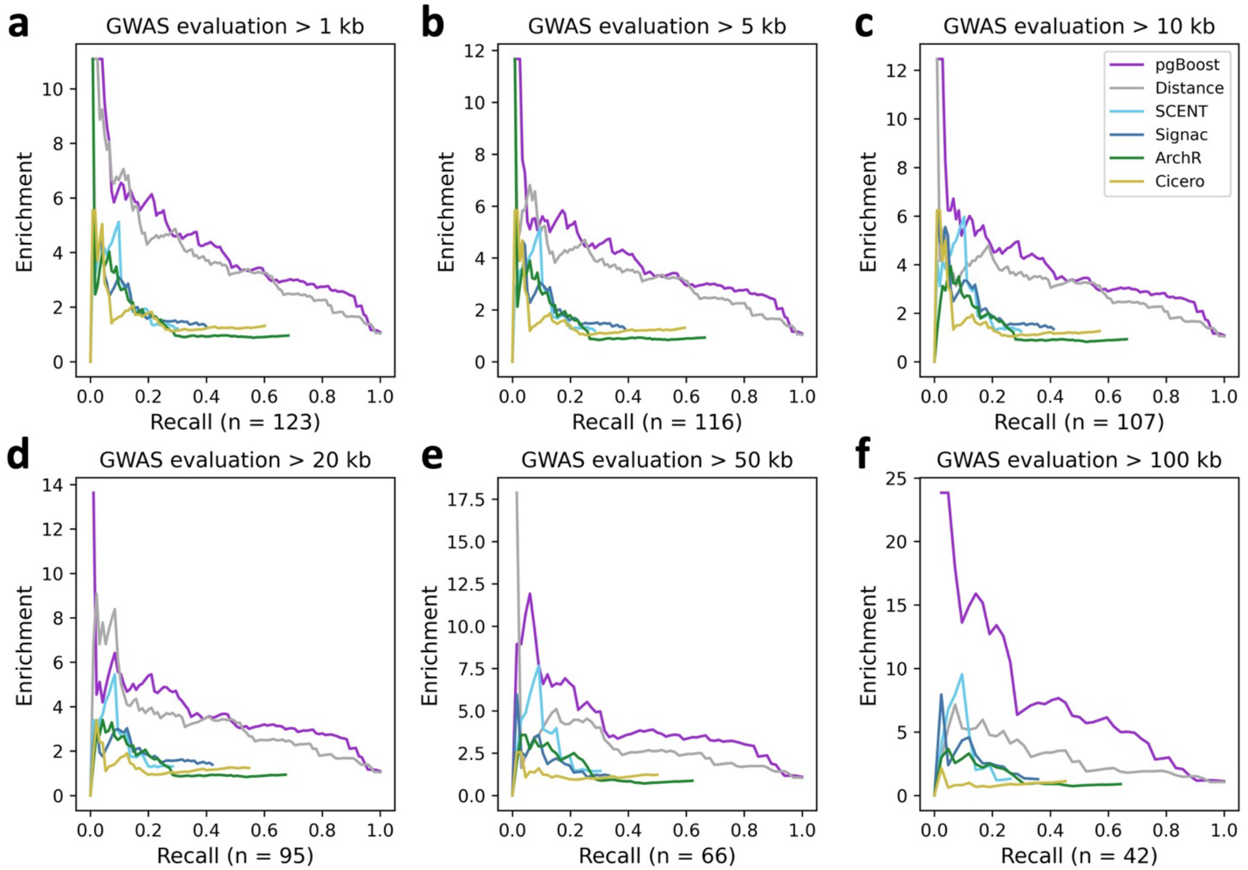


**SF19: Enrichment-recall curves of pgBoost and other methods on GWAS-derived evaluation data set.** Enrichment-recall curves of links predicted by pgBoost, distance, and 4 constituent methods for 123 non-coding SNP-gene pairs derived from fine-mapped GWAS variants with a unique fine-mapped coding variant within a 2 Mb window, at various distance thresholds. The number of positive evaluation links at each distance threshold is specified in parentheses.

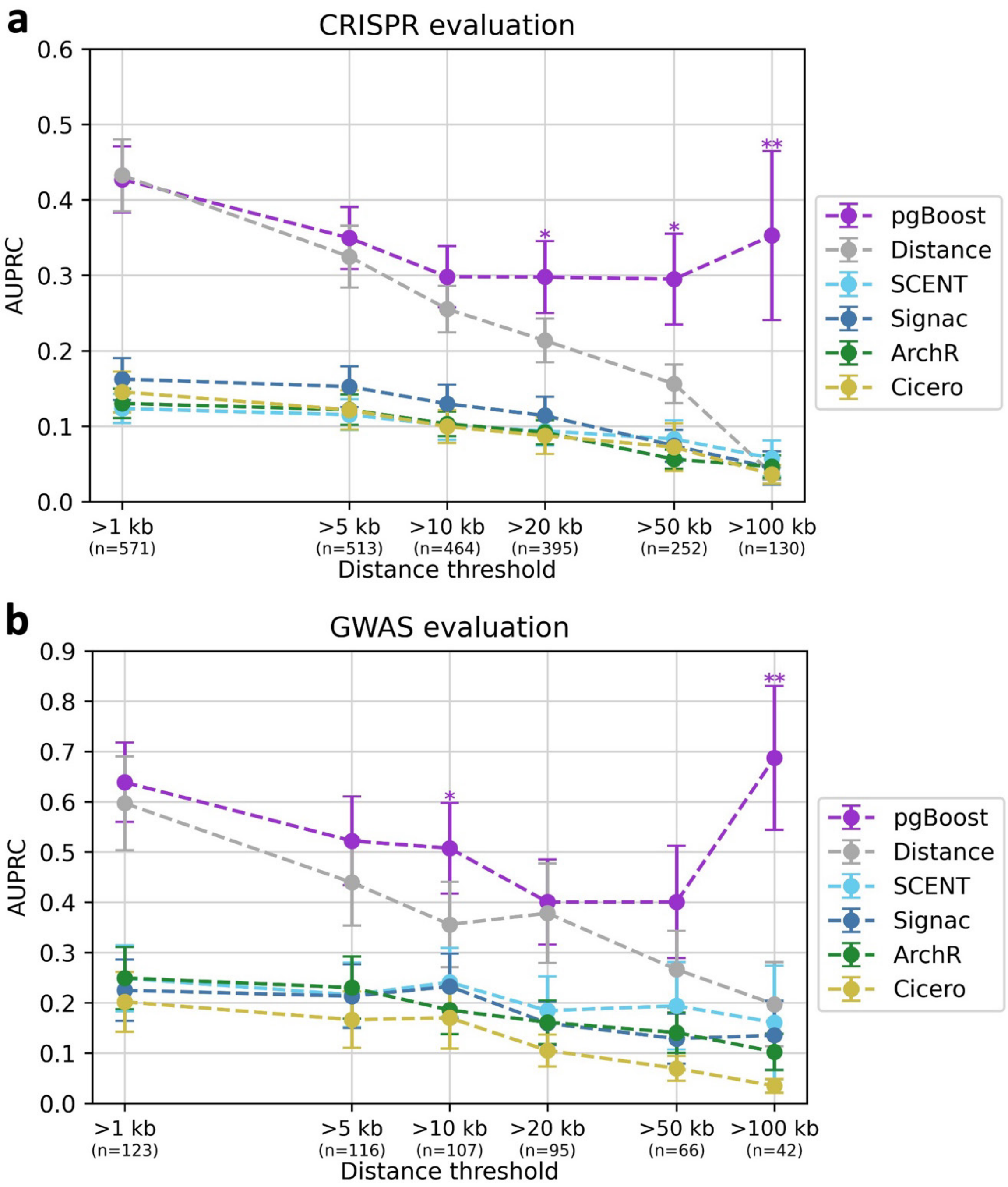


**SF20: AUPRC of pgBoost and other methods on gold-standard evaluation data sets. a)** Area under the precision-recall curve (AUPRC) across recall values (in [0,0.5]) of links predicted by pgBoost, distance, and 4 constituent methods for 571 links validated by CRISPR, at various distance thresholds. **b)** Area under the precision-recall curve (AUPRC) across recall values (in [0,0.25]) of links predicted by pgBoost, distance, and 4 constituent methods for 123 non-coding SNP-gene pairs derived from fine-mapped GWAS variants with a unique fine-mapped coding variant within a 2 Mb window, at various distance thresholds. The number of positive evaluation links at each distance threshold is specified in parentheses. Confidence intervals denote standard errors. Stars denote p-values for difference (*: $p < 0.05$, **: $p < 0.01$, ***: $p < 0.001$) of top method vs. each other method.

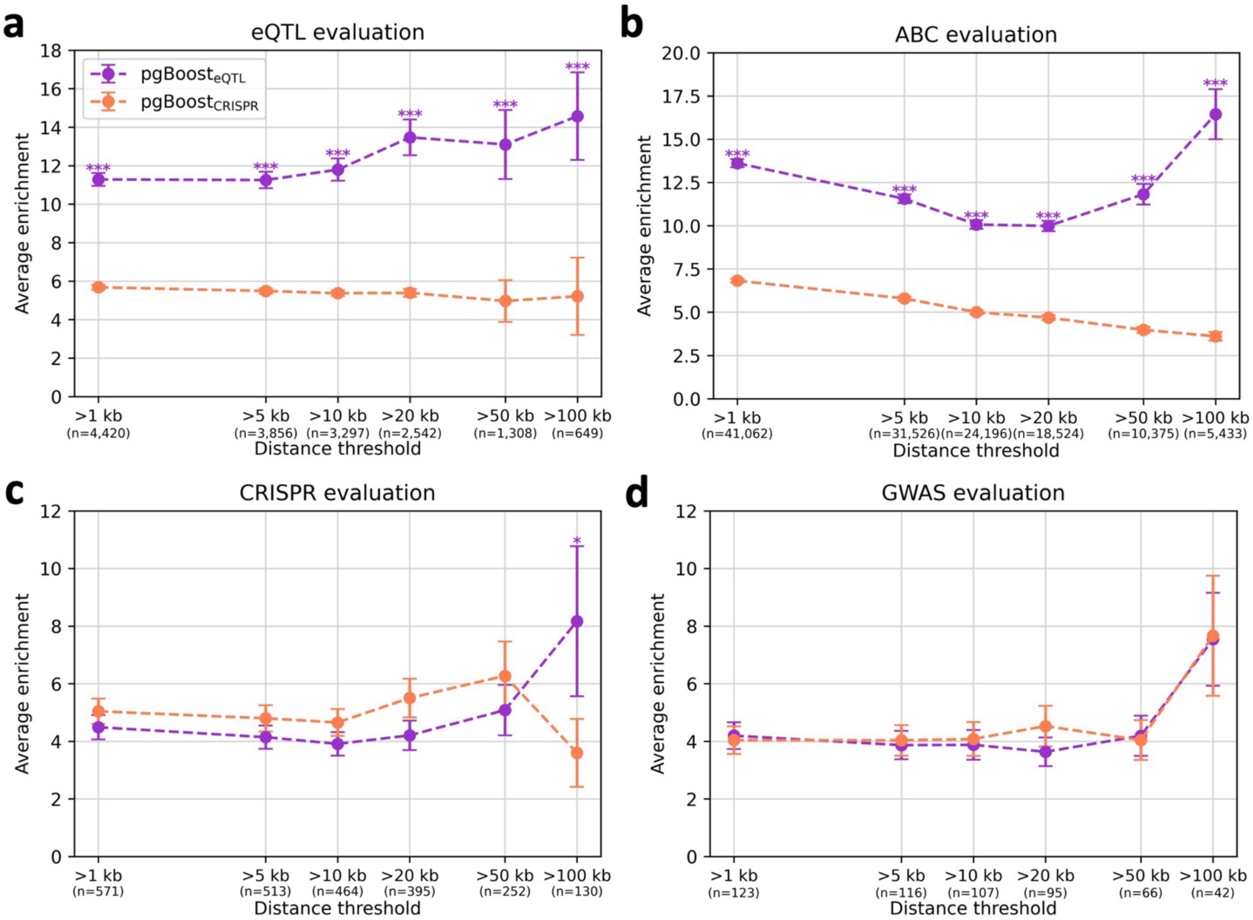


**SF21: Performance of pgBoost trained on eQTL vs. CRISPR data.** Average enrichment across recall values (in [0,1]) of links predicted by pgBoost trained on fine-mapped eSNP-eGene pairs vs. CRISPR-validated links for **(a)** 4,420 fine-mapped eSNP-eGene pairs attaining maximum PIP > 0.5 across GTEx tissues, **(b)** 41,062 SNP-gene pairs attaining maximum ABC score > 0.2 across 344 biosamples, **(c)** 571 links validated by CRISPR, and **(d)** 123 non-coding SNP-gene pairs derived from fine-mapped GWAS variants with a unique fine-mapped coding variant within a 2 Mb window, at various distance thresholds. The number of positive evaluation links at each distance threshold is specified in parentheses. Confidence intervals denote standard errors. Stars denote p-values for difference (*: $p < 0.05$, **: $p < 0.01$, ***: $p < 0.001$) of top method vs. each other method. For pgBoost$_{CRISPR}$, we defined a training set comprised of 571 SNP-gene links validated by CRISPR as positives and 1,071 links tested but not validated by CRISPR as negatives (**Supplementary Table 12**).

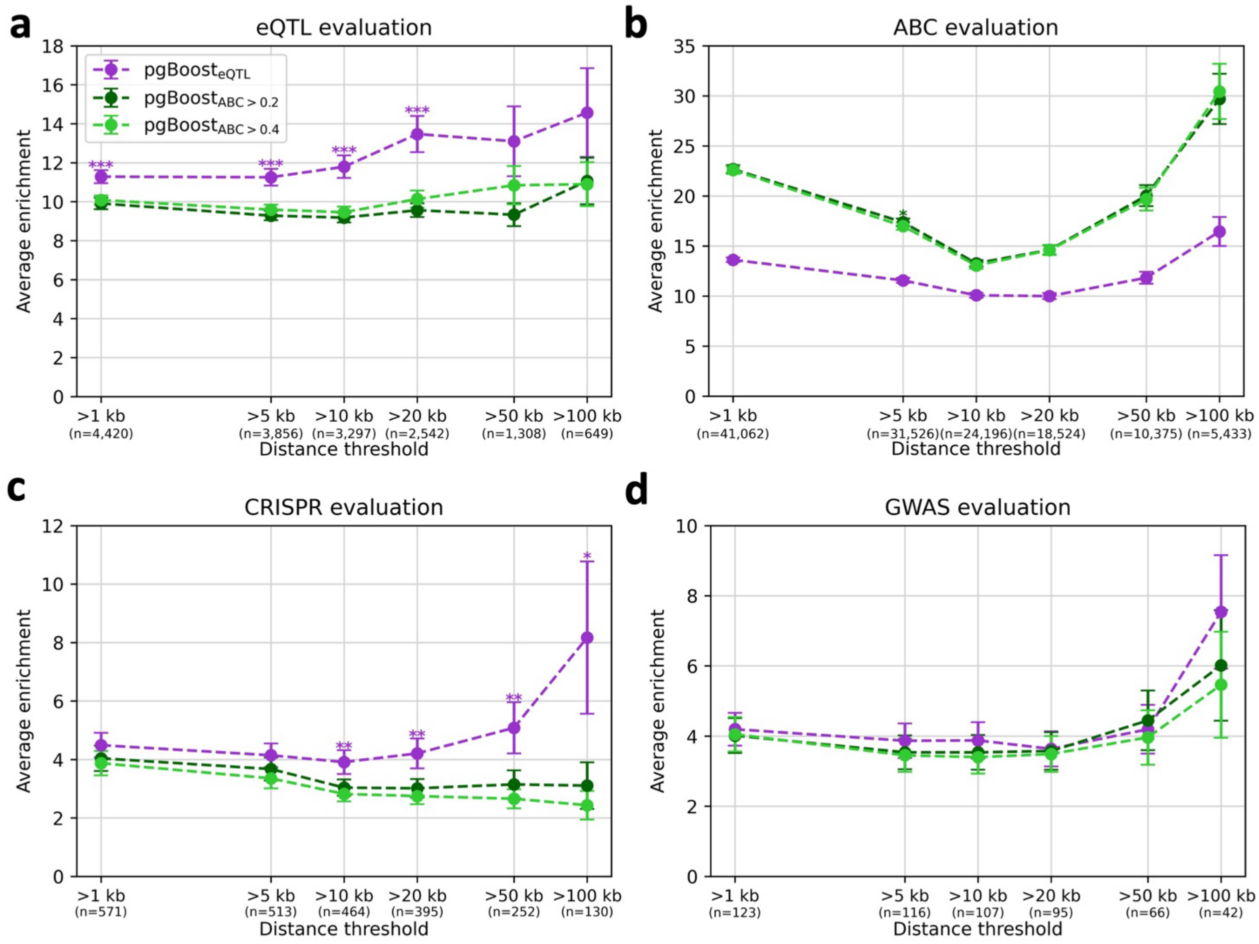


**SF22: Performance of pgBoost trained on eQTL vs. ABC data.** Average enrichment across recall values (in [0,1]) of links predicted by pgBoost trained on fine-mapped eSNP-eGene pairs vs. ABC links for **(a)** 4,420 fine-mapped eSNP-eGene pairs attaining maximum PIP > 0.5 across GTEx tissues, **(b)** 41,062 SNP-gene pairs attaining maximum ABC score > 0.2 across 344 biosamples, **(c)** 571 links validated by CRISPR, and **(d)** 123 non-coding SNP-gene pairs derived from fine-mapped GWAS variants with a unique fine-mapped coding variant within a 2 Mb window, at various distance thresholds. The number of positive evaluation links at each distance threshold is specified in parentheses. Confidence intervals denote standard errors. Stars denote p-values for difference (*: $p < 0.05$, **: $p < 0.01$, ***: $p < 0.001$) of top method vs. each other method. For pgBoost$_{ABC>0.2}$, we defined a training set comprised of 41,062 SNP-gene pairs attaining maximum ABC score > 0.2 across 344 biosamples as positives and 750,213 SNP-gene pairs including a gene in the positive set and attaining maximum ABC score < 0.015 across 344 biosamples as negatives. For pgBoost$_{ABC>0.4}$, we defined a training set comprised of 11,491 SNP-gene pairs attaining maximum ABC score > 0.2 across 344 biosamples as positives and 577,790 SNP-gene pairs including a gene in the positive set and attaining maximum ABC score < 0.015 across 344 biosamples as negatives.

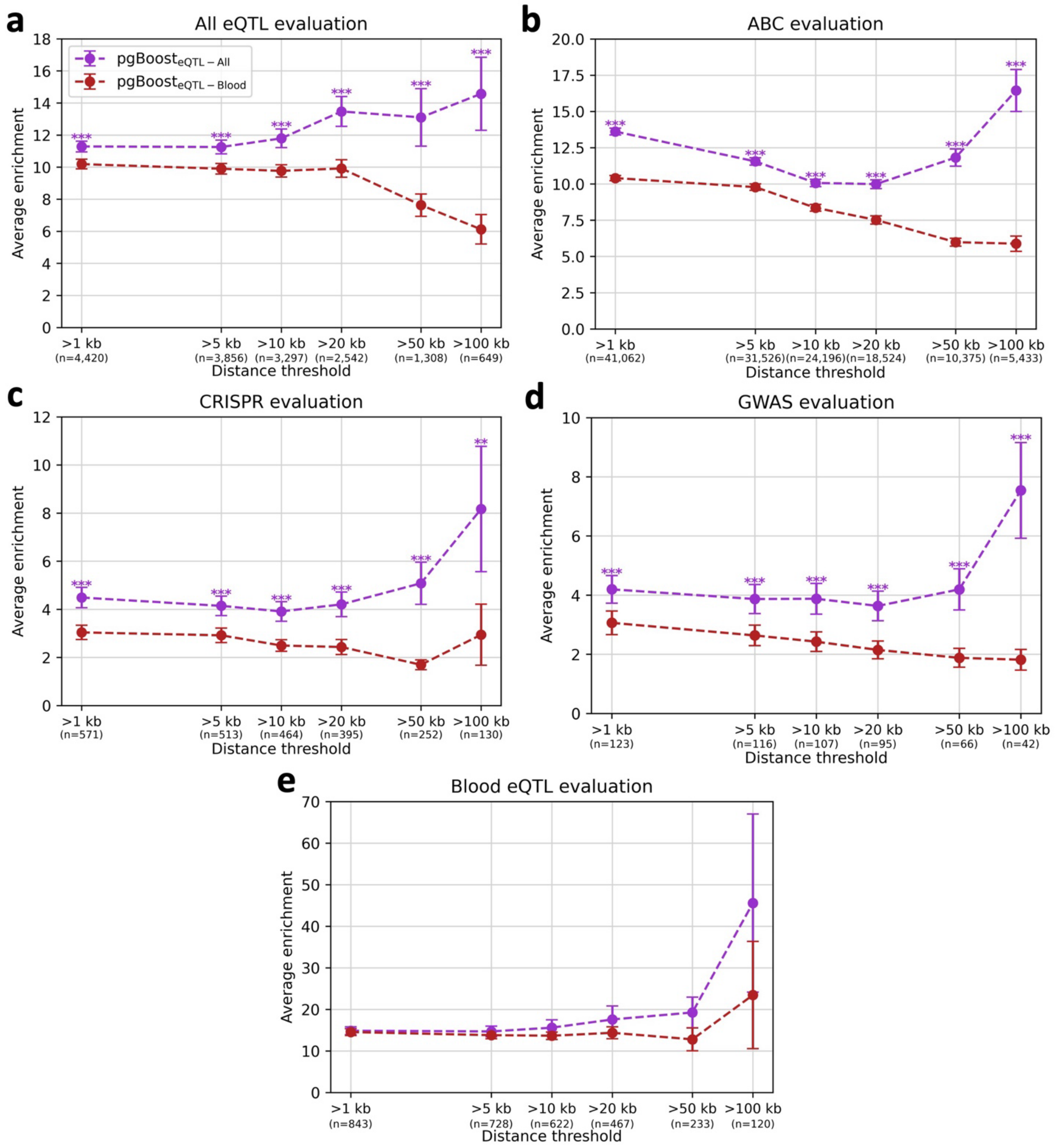


**SF23: Performance of pgBoost trained on eQTL data from all tissues vs. whole blood.** Average enrichment across recall values (in [0,1]) of links predicted by pgBoost trained on fine-mapped eSNP-eGene pairs from all GTEx tissues vs. fine-mapped eSNP-eGene pairs from whole blood for **(a)** 4,420 fine-mapped eSNP-eGene pairs attaining maximum PIP > 0.5 across GTEx tissues, **(b)** 41,062 SNP-gene pairs attaining maximum ABC score > 0.2 across 344 biosamples, **(c)** 571 links validated by CRISPR, **(d)** 123 non-coding SNP-gene pairs derived from fine-mapped GWAS variants with a unique fine-mapped coding variant within a 2 Mb window, and **(e)** 843 fine-mapped eSNP-eGene pairs attaining PIP > 0.5 in GTEx whole blood, at various distance thresholds. The number of positive evaluation links at each distance threshold is specified in parentheses. Confidence intervals denote standard errors. Stars denote p-values for difference

(*: $p < 0.05$, **: $p < 0.01$, ***: $p < 0.001$) of top method vs. each other method. For $pgBoost_{eQTL-Blood}$, we defined a training set of 843 eSNP-eGene pairs attaining PIP > 0.5 in whole blood as positives and 1,170 eSNP-eGene pairs attaining PIP < 0.01 in whole blood as negatives.

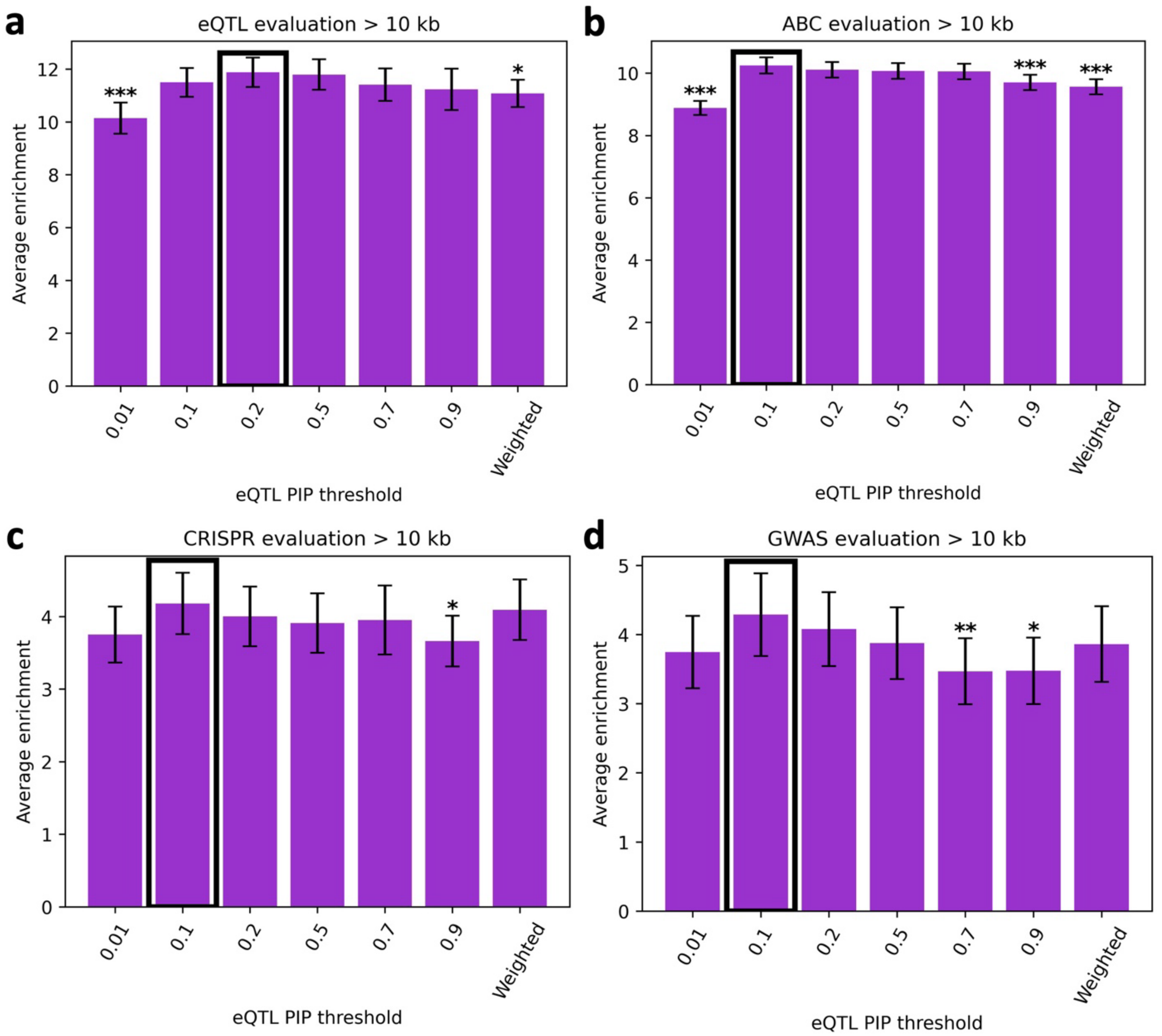


**SF24: Performance of pgBoost trained on various eQTL data sets.** Average enrichment across recall values (in [0,1]) of links predicted by pgBoost trained on eSNP-eGene pairs passing varying PIP thresholds for **(a)** 4,420 fine-mapped eSNP-eGene pairs attaining maximum PIP > 0.5 across GTEx tissues, **(b)** 41,062 SNP-gene pairs attaining maximum ABC score > 0.2 across 344 biosamples, **(c)** 571 links validated by CRISPR, and **(d)** 123 non-coding SNP-gene pairs derived from fine-mapped GWAS variants with a unique fine-mapped coding variant within a 2 Mb window. Because enrichment differences are highly similar across distance thresholds, results at >10kb are shown as barplots to aid in visual comparison. Confidence intervals denote standard errors. Stars denote p-values for difference (*: $p < 0.05$, **: $p < 0.01$, ***: $p < 0.001$) of focal method vs. top method (denoted in black outline). We defined training sets comprised of SNP-gene links attaining maximum PIP across GTEx tissues above each threshold as positives and SNP-gene links including a gene in the positive set and attaining maximum PIP < 0.1 across GTEx tissues as negatives. Using this strategy, we defined training sets of fine-mapped eSNP-eGene pairs passing PIP thresholds of 0.01 (94,969 positives), 0.1 (20,229 positives), 0.2 (11,204 positives), 0.5 (4,420 positives), 0.7 (3,008 positives), 0.9 (1,951 positives). We also assessed a "weighted" training set where SNP-gene pairs attaining PIP > 0.8 are assigned a weight of 4, SNP-gene pairs attaining PIP > 0.4 and < 0.8 are assigned a weight of 2, and SNP-gene pairs attaining PIP > 0.1 and < 0.4 are assigned a weight of 1 (and SNP-gene links including a gene in the positive set and attaining maximum PIP < 0.1 are defined as negatives).

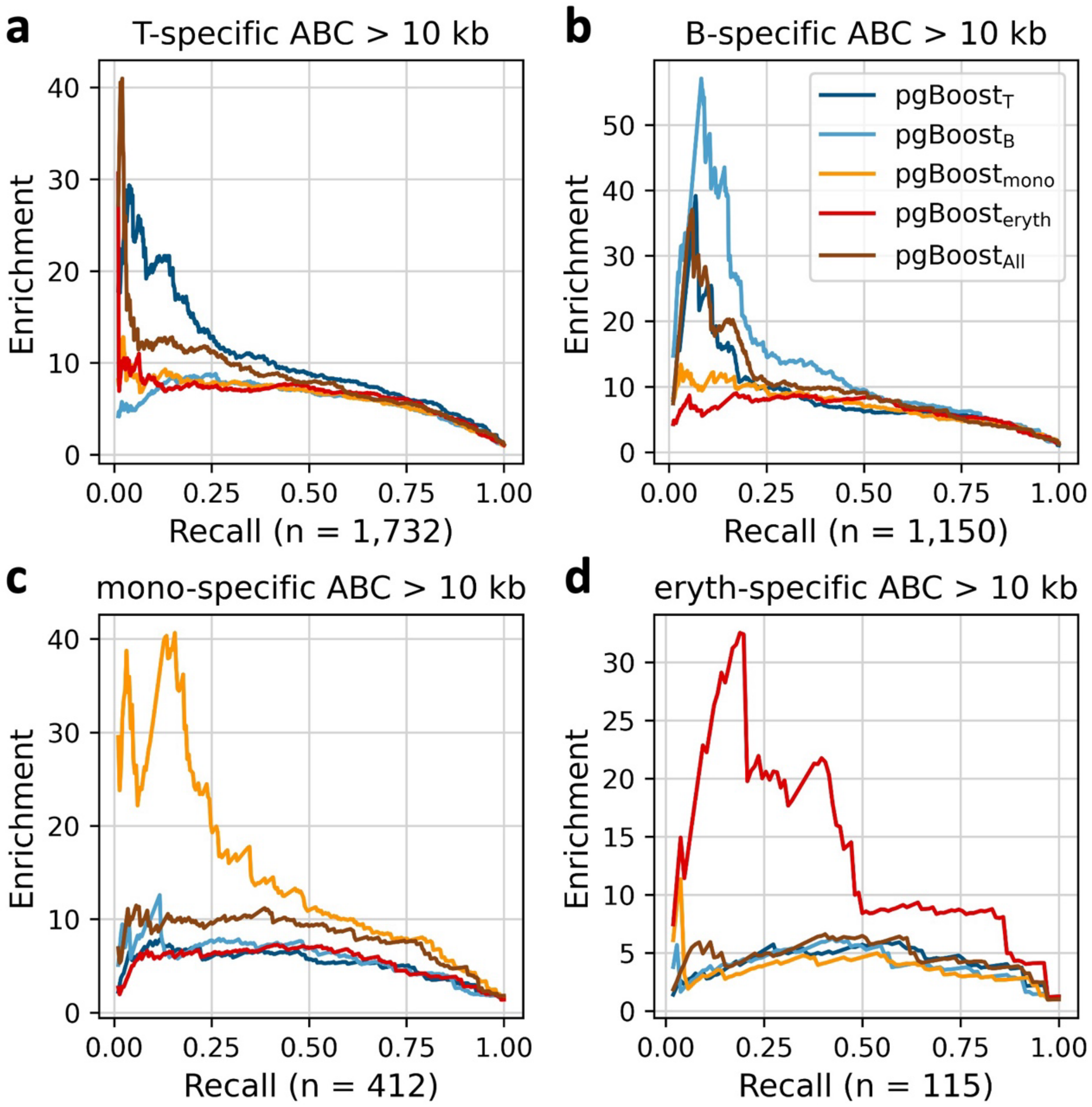


**SF25: Enrichment-recall curves of pgBoost restricted to features from a focal cell type in cell-type-specific ABC evaluation data sets.** Enrichment-recall curves of links predicted by pgBoost restricted to features from a focal cell type (All denotes all features) for cell-type-specific ABC evaluation links detected in biosamples related to **(a)** T cells, **(b)** B cells, **(c)** myeloid cells, and **(d)** erythroid cells, at distance >10kb. The number of positive links in each evaluation set is specified in parentheses.

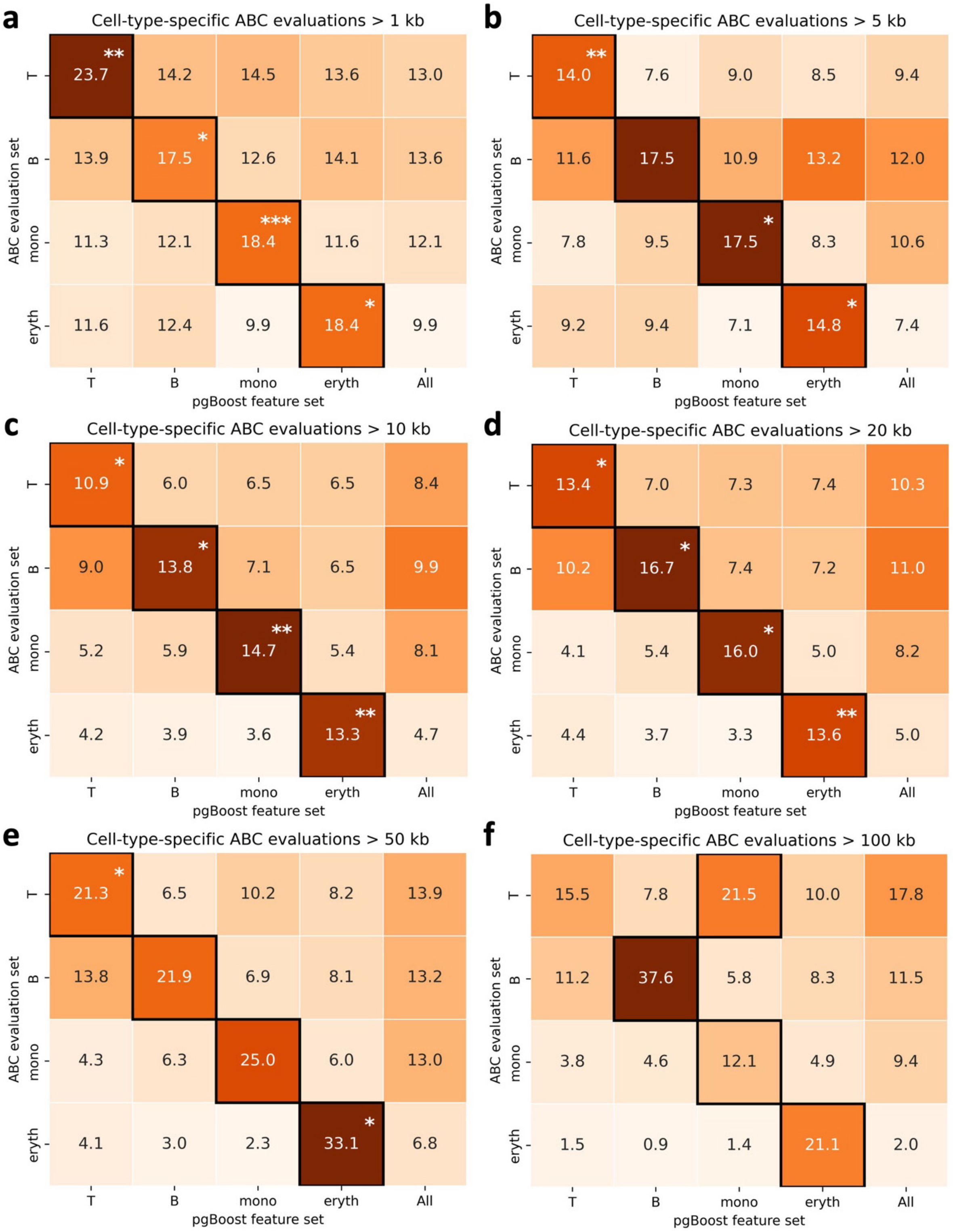


**SF26: Performance of pgBoost restricted to features from a focal cell type in cell-type-specific ABC evaluation data sets.** Average enrichment across recall values (in [0,1]) of links predicted by pgBoost restricted to features from a focal cell type (columns; All denotes all features) for cell-type-specific ABC evaluation links (rows), at various distance thresholds. Stars denote p-values for difference (*: $p < 0.05$, **: $p < 0.01$, ***: $p < 0.001$) of top method (defined separately for each row and denoted in black outline) vs. each other method.

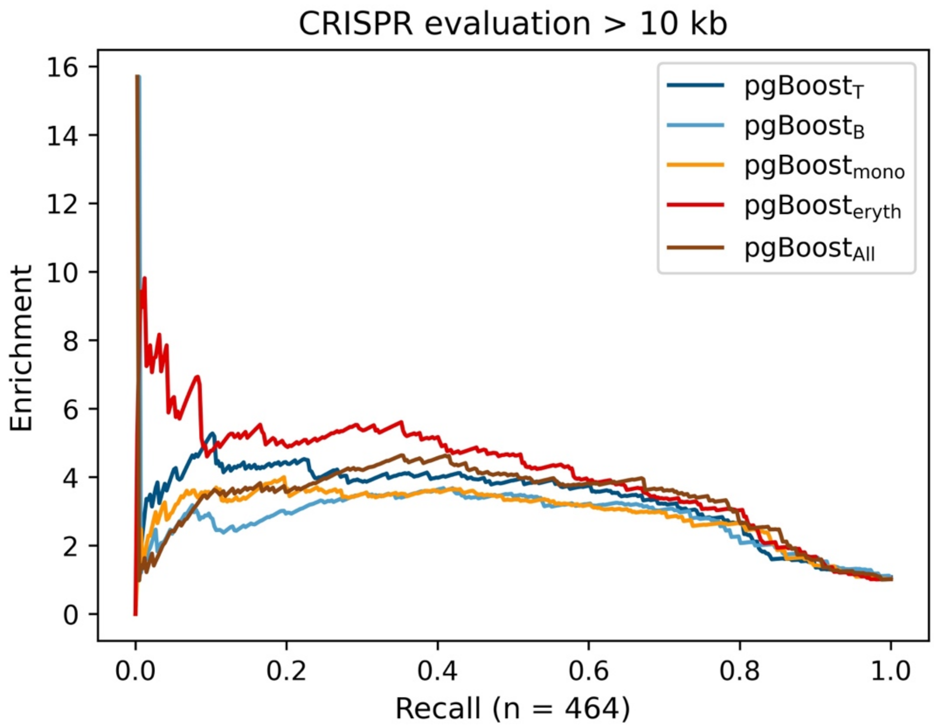


**SF27: Enrichment-recall curves of pgBoost restricted to features from a focal cell type on CRISPR evaluation data set.** Enrichment-recall curves of links predicted by pgBoost restricted to features from a focal cell type (All denotes all features) for 571 links validated by CRISPR. The number of positive evaluation links is specified in parentheses.

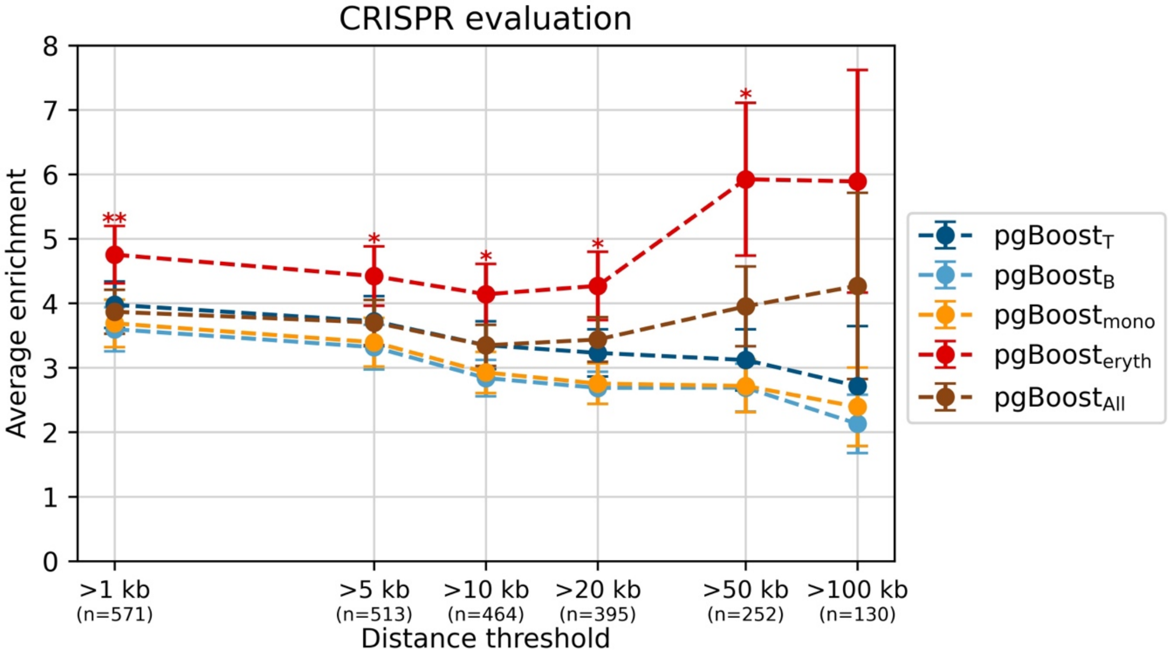


**SF28: Performance of pgBoost restricted to features from a focal cell type on CRISPR evaluation data set.** Average enrichment across recall values (in [0,1]) of links predicted by pgBoost restricted to features from a focal cell type (All denotes all features) for 571 links validated by CRISPR, at various distance thresholds. The number of positive evaluation links at each distance threshold is specified in parentheses. Confidence intervals denote standard errors. Stars denote p-values for difference (*: $p < 0.05$, **: $p < 0.01$, ***: $p < 0.001$) of top method vs. each other method.

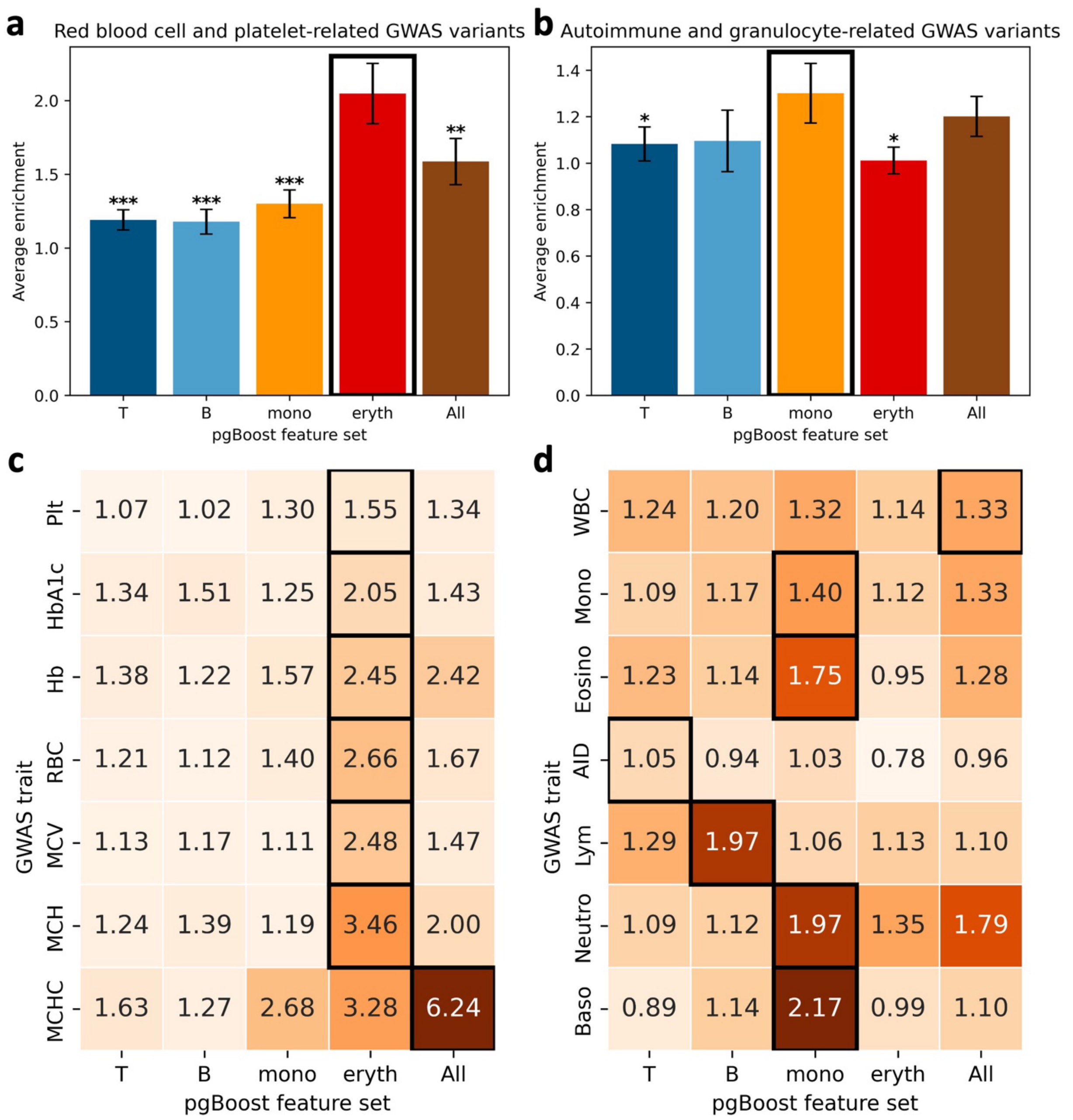


**SF29: Performance of pgBoost restricted to features from a focal cell type on fine-mapped GWAS variants.** Average enrichment across recall values (in [0,1]) of variants linked to genes by pgBoost restricted to features from a focal cell type (All denotes all features) for **(a)** n = 493 fine-mapped variants (PIP > 0.2) for 7 red blood cell or platelet-related blood cell traits, **(b)** n = 518 fine-mapped variants (PIP > 0.2) for 7 autoimmune diseases or granulocyte-related blood cell traits, **(c)** fine-mapped variants for individual traits from **(a)**, **(d)** fine-mapped variants for individual traits from **(b)**. Confidence intervals denote standard errors, and stars denote p-values for difference (*: p < 0.05, **: p < 0.01, ***: p < 0.001) of focal method vs. top method (denoted in black outline). Plt = platelet count, HbA1c = hemoglobin A1c, Hb = hemoglobin, RBC = red blood cell count, MCV = mean corpuscular volume, MCH = mean corpuscular hemoglobin, MCHC = mean corpuscular hemoglobin concentration, WBC = white blood cell count, Mono = monocyte count, Eosino = eosinophil count, AID = autoimmune disease, Lym = lymphocyte count, Neutro = neutrophil count, Baso = basophil count.

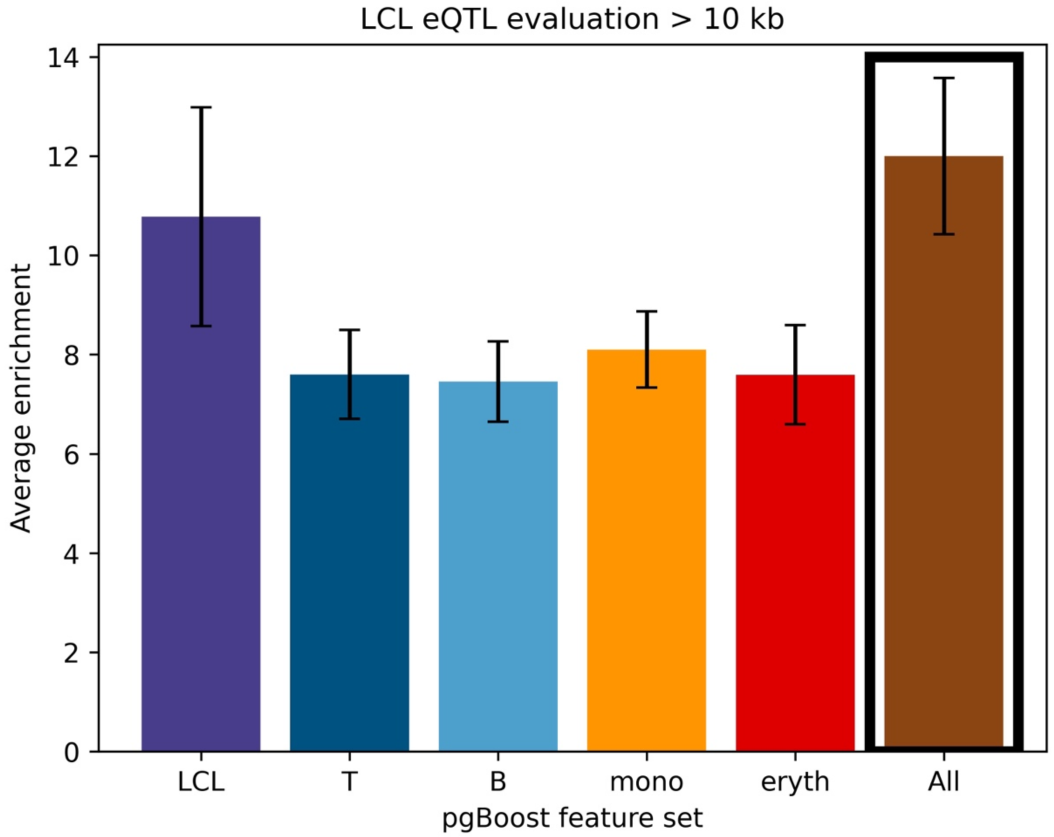


**SF30**: **Performance of pgBoost restricted to features from a focal cell type on eQTL data from LCL.** Average enrichment across recall values (in [0,1]) of links predicted by pgBoost restricted to features from a focal cell type (columns; All denotes all features) for fine-mapped eSNP-eGene pairs attaining maximum PIP > 0.2 in GTEx EBV-transformed lymphocytes or LCL, at distance >10kb (number of evaluation links: n = 160). Confidence intervals denote standard errors. Top method denoted in black outline.

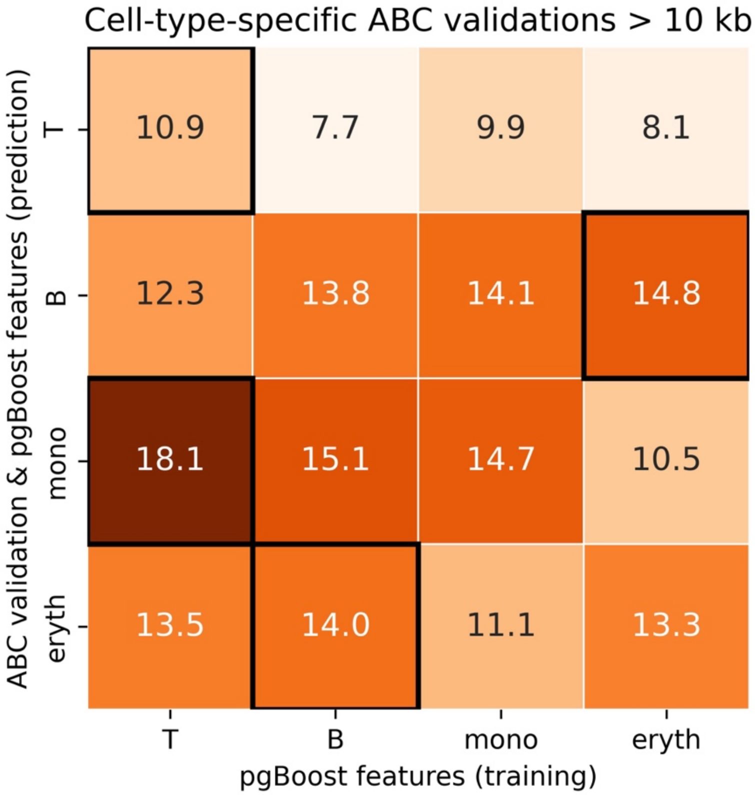


**SF31: Performance of pgBoost restricted to features from a focal or non-focal cell type for training only in evaluation data sets relevant to the focal cell type.** Average enrichment across recall values (in [0,1]) of links predicted by a pgBoost model restricted to features from an individual cell type during training (columns) for cell-type-specific ABC evaluation links (rows), where predictions are always generated by applying models to linking scores from the evaluation cell type. Enrichments are measured at distance >10kb (number of evaluation links in each cell type: $n_T$ = 1,732, $n_B$ = 1,150, $n_{mono}$ = 412, $n_{eryth}$ = 115). The top-performing model within each row never attains p-value < 0.05 vs. each other method. This result suggests that pgBoost models trained using linking scores from individual cell types are generally portable across cell types.

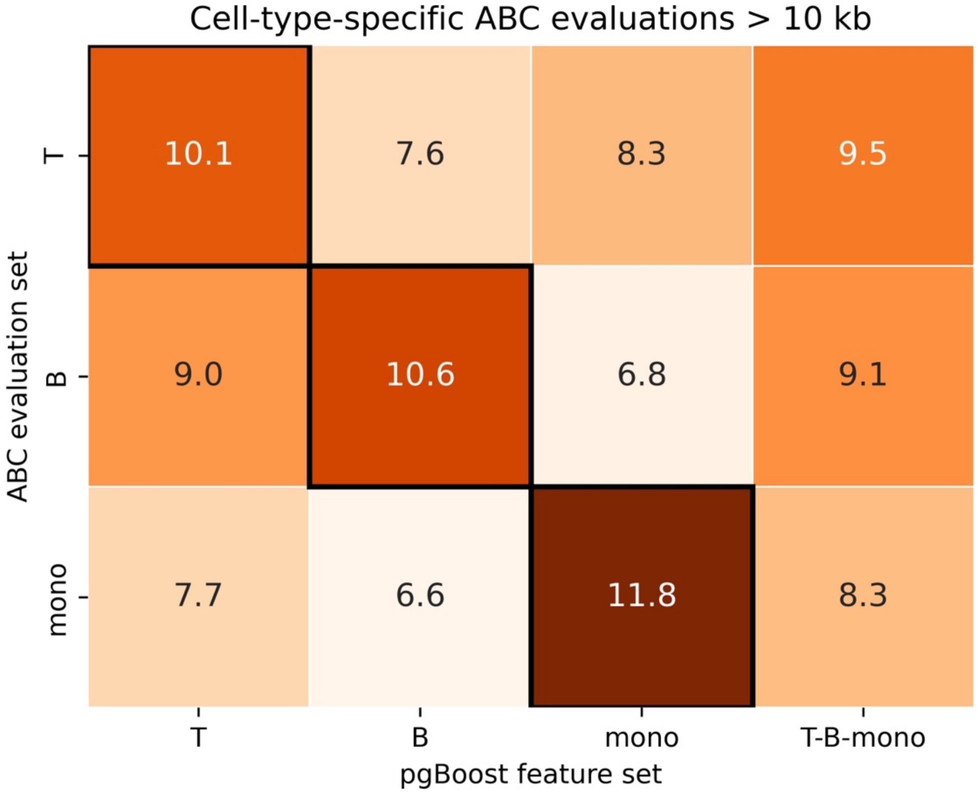


**SF32: Performance of pgBoost restricted to features from a focal cell type across two single-cell data sets in cell-type-specific ABC evaluation data sets.** Average enrichment across recall values (in [0,1]) of links predicted by pgBoost restricted to features from a focal cell type (columns; T-B-mono denotes all features) included in the Luecken BMMC and 10X PBMC data sets (**Table 2** and **Supplementary Table 2**) for cell-type-specific ABC evaluation links (rows), at distance >10kb. Top method is defined separately for each row and denoted in black outline.

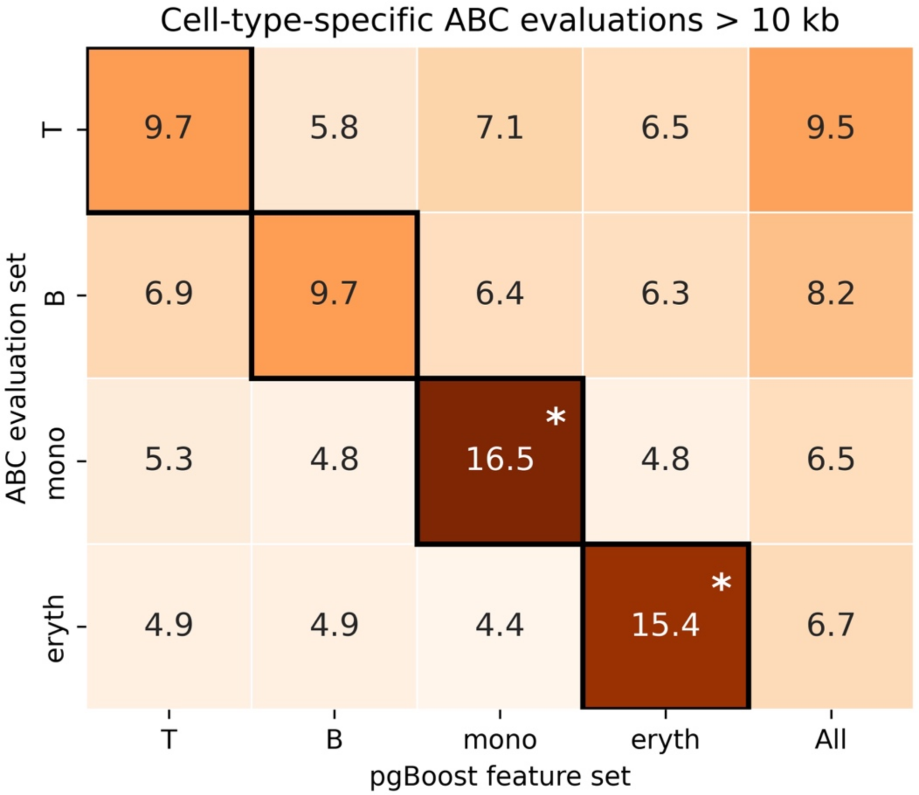


**SF33**: **Performance of pgBoost restricted to features from a focal cell type in ABC evaluation links passing a stringent threshold for cell-type-specificity.** Average enrichment across recall values (in [0,1]) of links predicted by pgBoost restricted to features from a focal cell type (columns; All denotes all features) for cell-type-specific ABC evaluation links (rows) defined by SNP-gene links attaining ABC score > 0.2 in biosample(s) related to the focal cell type and below a more stringent threshold in non-focal cell types (ABC score < 0.05), at distance >10kb (number of evaluation links: $n_T$ = 1,087, $n_B$ = 743, $n_{mono}$ = 213, $n_{eryth}$ = 72). Top method is defined separately for each row and denoted in black outline.

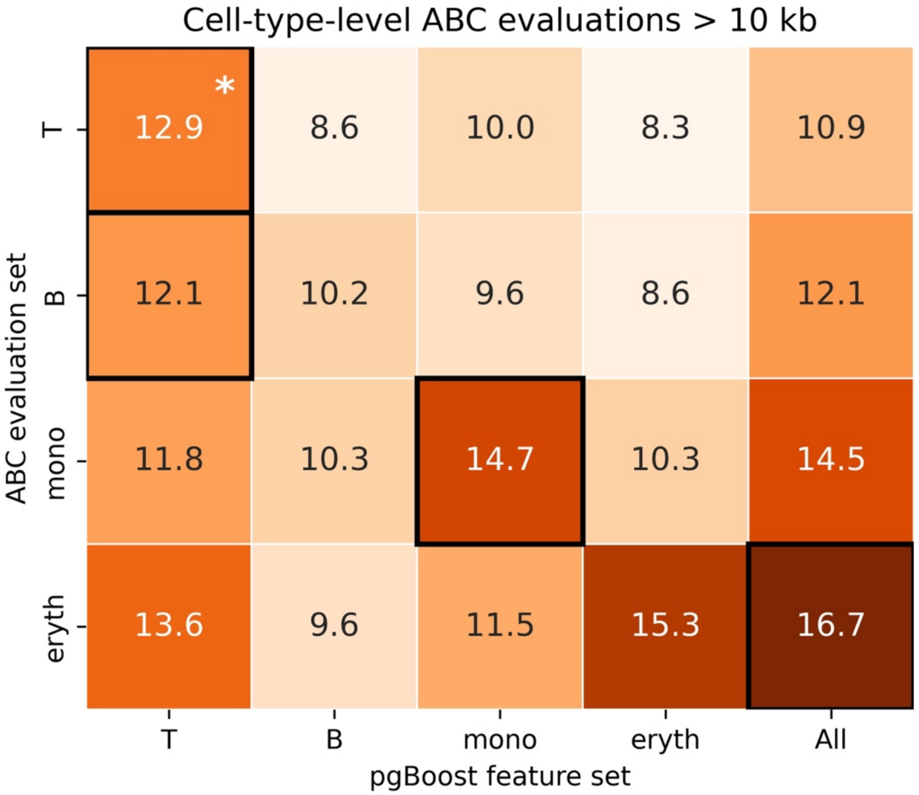


**SF34: Performance of pgBoost restricted to features from a focal cell type in cell-type-level ABC evaluation data sets.** Average enrichment across recall values (in [0,1]) of links predicted by pgBoost restricted to features from a focal cell type (columns; All denotes all features) for cell-type-level ABC evaluation links (rows) defined by SNP-gene links attaining ABC score > 0.2 in biosample(s) related to the focal cell type (irrespective of ABC scores attained in other biosamples) at distance >10kb (number of evaluation links: $n_T$ = 16,932, $n_B$ = 14,597, $n_{mono}$ = 7,238, $n_{eryth}$ = 2,435). Top method is defined separately for each row and denoted in black outline.
